# Physical Activity, Sedentary Behavior and Microbiome: A Systematic Review and Meta-analysis

**DOI:** 10.1101/2024.01.29.24301919

**Authors:** Inmaculada Pérez-Prieto, Abel Plaza-Florido, Esther Ubago-Guisado, Francisco B. Ortega, Signe Altmäe

## Abstract

**Background:** The effects of physical activity and sedentary behavior on human health are well known, however, the molecular mechanisms are poorly understood. Growing evidence points to physical activity as an important modulator of the microbial composition, while evidence of sedentary behavior is scarce. We aimed to synthesize and meta-analyze the current evidence about the effects of physical activity and sedentary behavior on microbiome across different body sites and in different populations.

**Methods:** A systematic search in PubMed, Web of Science, Scopus and Cochrane databases was conducted until September 2022. Random-effects meta-analyses including cross-sectional studies (active *vs*. inactive / athletes *vs*. non-athletes) or trials reporting the chronic effect of physical activity interventions on gut microbiome alpha-diversity in healthy individuals were performed.

**Results:** Ninety-one studies were included in this systematic review. Our meta-analyses of 2632 participants indicated no consistent effect of physical activity on microbial alpha-diversity, although there seems to be a trend toward a higher microbial richness in athletes compared to non-athletes. We observed an increase in short-chain fatty acids-producing bacteria such as *Akkermansia*, *Faecalibacterium*, *Veillonella* or *Roseburia* in active individuals and after physical activity interventions.

**Conclusions:** Physical activity levels were positively associated with the relative abundance of short-chain fatty acids-producing bacteria. Athletes seem to have a richer microbiome compared to non-athletes. However, high heterogeneity between studies avoids to obtain conclusive information on the role of physical activity in microbial composition. Future multi-omics studies would enhance our understanding of the molecular effects of physical activity and sedentary behavior on the microbiome.

## 1. Introduction

It is well-known that physical activity (PA) (i.e., any movement produced by skeletal muscles which demands a higher energy expenditure than in resting conditions) can improve different health-related outcomes such as insulin resistance, adiposity, and fitness, among others ^1,2^. A related yet different construct is sedentary behavior (SB) (i.e., a behavior characterized by an energy expenditure of 1.5 or fewer metabolic equivalents [METs]), and is associated with a higher risk of different diseases ^3,4^. Thus, increasing PA and reducing SB have been considered to prevent and treat multiple chronic diseases^5^. However, the molecular mechanisms underlying the health benefits of PA (acute or chronic effects) and the adverse effects of SB on health are poorly understood ^6^.

In the last decades, a new sight of the human being as a set of microbial and human cells has emerged ^7^. The collection of microorganisms encompasses bacteria, viruses, fungi and archaea that inhabits our body is defined as the microbiota and is at least as abundant as the number of human cells ^8^. The genomes of the abovementioned microorganisms (i.e., microbiota) are called the microbiome, which is considered “our second genome” and “our last organ” due to its important role in human physiology ^9,10^. Metagenomics studies (e.g., marker gene sequencing and whole metagenome sequencing) led characterization of microbiome composition using three common analyses: (1) alpha-diversity, that characterizes the microbial diversity within a sample considering richness and evenness (i.e., the number and the relative abundance of microbes); (2) beta-diversity, which measures the diversity between samples assigning a numerical value for every pair of samples to determine microbial community-level dissimilarities; (3) differential abundance analysis, that identifies those microorganisms that differ in abundance when compared different samples.

There is evidence indicating that environmental and lifestyle factors such as pollutants, antibiotics, diet, lack of PA and increased SB, among others, may have a negative impact on microbiome composition and function leading to the disruption of the microbial homeostasis (i.e., dysbiosis) ^11–13^. In fact, microbial dysbiosis has been associated with the development of multiple diseases such as obesity ^14,15^, type 2 diabetes ^16^, and cancer ^17,18^, among others. Thus, there is a growing interest to determine the composition of the “healthy core” microbiome and the factors that could shape the microbial communities, such as PA and SB, in order to design new therapeutic interventions ^19,20^.

Particularly, PA has been described as one of the most important modulators of the microbiome, while little is known about the effect of SB on microbial communities due to the limited number of studies ^21,22^. Recent advances in meta-omics-based studies (i.e., marker gene sequencing, metagenomics, meta-transcriptomics, meta-proteomics, and meta-metabolomics) allow the identification of the molecular pathways regulated by PA ^23^. Thus, the effect of PA on the microbiome, especially on the gut microbial communities, is a research topic of increasing interest ^24,25^. In the last years, several systematic reviews reported the effects of PA on the gut microbiome of healthy adults ^26–29^, older adults ^30^ and adults with obesity and type 2 diabetes ^31,32^. In addition, a systematic review on the effect of aerobic athletic performance has been recently published ^33^. However, the aforementioned systematic reviews showed inconsistent findings from observational and intervention studies ^26–32^. Therefore, there is a need to synthesize the whole body of knowledge about the effect of PA and SB on the microbiome including healthy (e.g. non-athletes and professional athletes), unhealthy populations (e.g. obesity, diabetes, cancer), different stages of life (i.e., children, young and older adults), and different body niches (e.g. gut, saliva, vaginal, etc.) through metagenomics approaches.

The current study aimed: 1) to summarize all the studies available about the relationship of PA and SB (observational and intervention studies) with microbiome performing metagenomics in humans and 2) meta-analyse the available data.

## 2. Material and methods

This systematic review and meta-analysis was conducted following the Preferred Reporting Items for Systematic Reviews and Meta-Analyses guidelines (PRISMA) ^34^. The review protocol was registered in the International Prospective Register of Systematic Reviews (PROSPERO; http://www.crd.york.ac.uk/PROSPERO) with the reference number: CRD42022298526.

### 2.1 Search strategy

A systematic search was conducted in PubMed, Web of Science, SCOPUS, and Cochrane electronic databases from inception to September 29, 2022. Search terms were included based on the sports science and microbiome terms of interest. **Table 1** includes a list with the main terms and their definitions related to microbiome field used in this systematic search. **Electronic Supplementary Table S1** illustrates the search terms and strategy for each database.

**Table 1.**
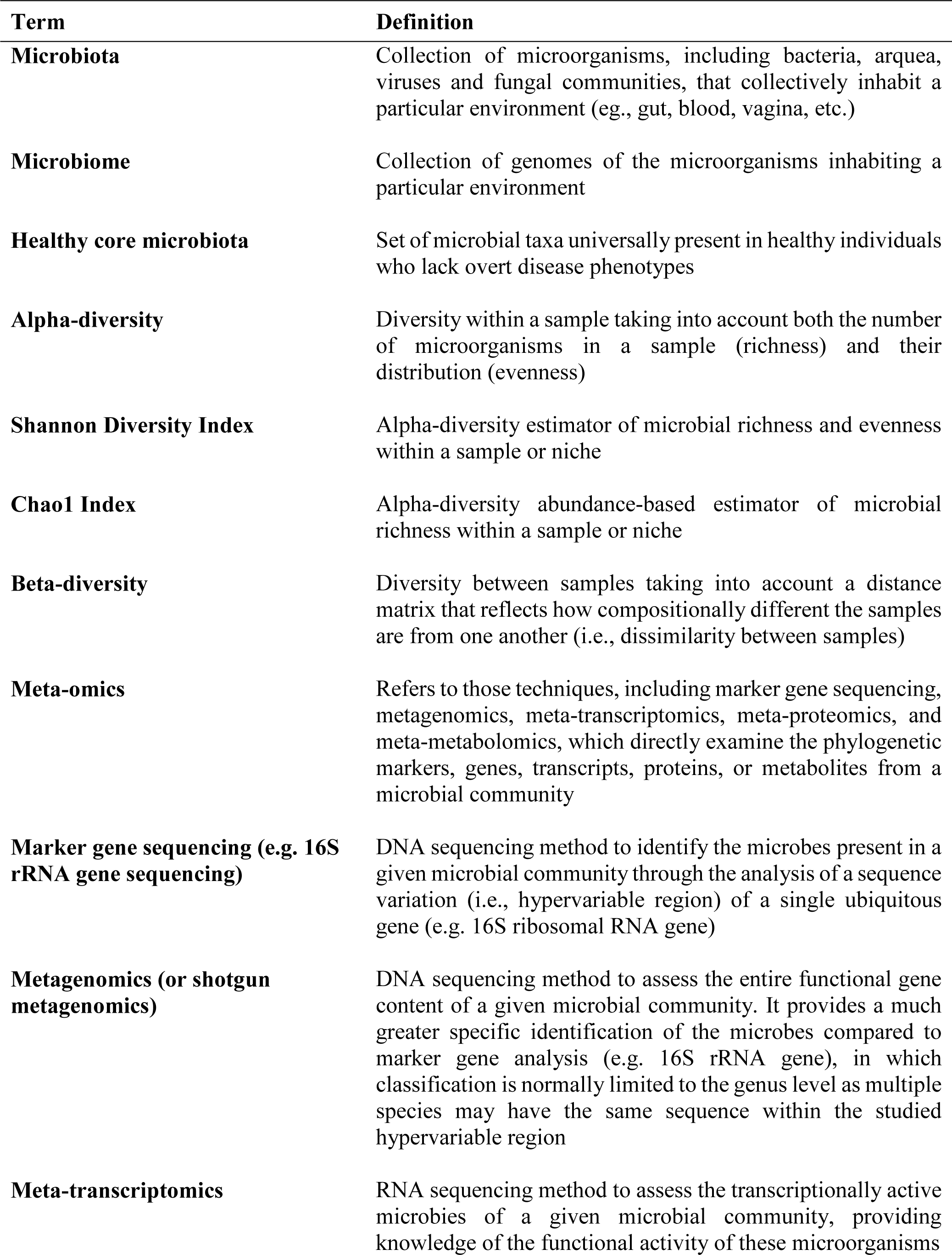
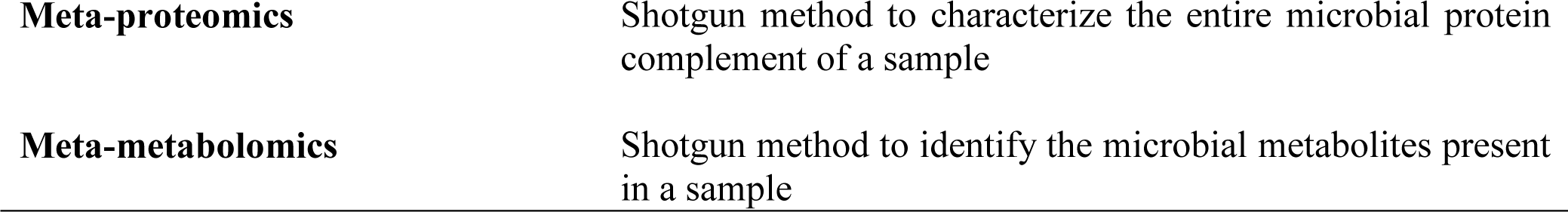
Definition of main microbiome-related terms used in this systematic review.

| **Term** | **Definition** |
| --- | --- |
| **Microbiota** | Collection of microorganisms, including bacteria, archaea, viruses and fungal communities, that collectively inhabit a particular environment (eg., gut, blood, vagina, etc.) |
| **Microbiome** | Collection of genomes of the microorganisms inhabiting a particular environment |
| **Healthy core microbiota** | Set of microbial taxa universally present in healthy individuals who lack overt disease phenotypes |
| **Alpha-diversity** | Diversity within a sample taking into account both the number of microorganisms in a sample (richness) and their distribution (evenness) |
| **Shannon Diversity Index** | Alpha-diversity estimator of microbial richness and evenness within a sample or niche |
| **Chao1 Index** | Alpha-diversity abundance-based estimator of microbial richness within a sample or niche |
| **Beta-diversity** | Diversity between samples taking into account a distance matrix that reflects how compositionally different the samples are from one another (i.e., dissimilarity between samples) |
| **Meta-omics** | Refers to those techniques, including marker gene sequencing, metagenomics, meta-transcriptomics, meta-proteomics, and meta-metabolomics, which directly examine the phylogenetic markers, genes, transcripts, proteins, or metabolites from a microbial community |
| **Marker gene sequencing (e.g. 16S rRNA gene sequencing)** | DNA sequencing method to identify the microbes present in a given microbial community through the analysis of a sequence variation (i.e., hypervariable region) of a single ubiquitous gene (e.g. 16S ribosomal RNA gene) |
| **Metagenomics (or shotgun metagenomics)** | DNA sequencing method to assess the entire functional gene content of a given microbial community. It provides a much greater specific identification of the microbes compared to marker gene analysis (e.g. 16S rRNA gene), in which classification is normally limited to the genus level as multiple species may have the same sequence within the studied hypervariable region |
| **Meta-transcriptomics** | RNA sequencing method to assess the transcriptionally active microbes of a given microbial community, providing knowledge of the functional activity of these microorganisms |

### 2.2 Study selection criteria

The inclusion criteria were as follow: (1) all observational studies (longitudinal or cross-sectional) that report the association of PA and/or SB with microbiome; (2) all original studies that included the effect of PA (acute and/or chronic effects) on microbiome. The exclusion criteria were: (1) studies addressing the effect of PA (acute or chronic effects) on microbiome containing diet modifications, probiotics and prebiotics supplements or caloric restriction, in which it was not possible to isolate the independent effect of PA, (2) non-eligible publication types, such as editorials, study protocols, letters to the editors, meeting abstracts, or review articles, (3) studies written in any language other than English or Spanish.

The selection process of the studies resulting from the literature search was performed using the software “Covidence” (https://www.covidence.org/), which detected duplicates. After removing the duplicates, the articles were first independently filtered by title/abstract screening by two researchers (I.P.P and A.P.F). Those articles that met the inclusion criteria were selected for the full-text review. Conflictive articles were solved through common consensus by the same researchers (I.P.P and A.P.F). Any article that did not meet the eligibility criteria was excluded. The quality assessment of the included studies was independently conducted by I.P.P and A.P.F (see **Electronic Supplementary Material Appendix S1**).

### 2.3 Data extraction

For each study, one researcher (I.P.P) conducted the data extraction including the following information: (1) author’s name and date of publication, (2) study design, (3) characteristics of the population (number of participants, sex, age and ethnicity), (4) characteristics of the exposure (i.e., PA or SB), (5) sample origin, (6) dependent outcome (i.e., DNA extraction method, detection method of the microbiome and sequencing platform), (7) dietary record, (8) main findings and (9) raw data availability. A second researcher (A.P.F) performed a double-check for data correction.

### 2.4 Data synthesis and meta-analysis

We conducted three meta-analyses including cross-sectional studies (active *vs*. inactive / athletes *vs*. non-athletes) or trials reporting the chronic effect of PA interventions on gut microbiome diversity (specifically alpha-diversity, expressed by the Shannon diversity and Chao1 indexes) in healthy individuals (see **Electronic Supplementary Material Appendix S1** for detailed explanation).

Statistical analyses were performed using the Comprehensive Meta-Analysis software (version 3; Biostat Inc.,1385, NJ, USA). The effect size was calculated using Cohen’s d and 95% confidence intervals (CIs) for standardized mean difference (SMD). Pooled SMD was estimated using a random-effects model. Heterogeneity between studies was assessed using the I^2^ statistics, which represents the percentage of total variation across studies, considering I^2^ values of 25%, 50%, and 75% as low, moderate and high heterogeneity, respectively ^35^. A p value of less than 0.05 was considered statistically significant.

## 3. Results

### 3.1 General overview

PRISMA checklist 2020 reflects the appropriateness of the methods performed in this systematic review and meta-analysis (**Electronic Supplementary Tables S2, S3**). **Figure 1** illustrates the PRISMA flow diagram of the search process. A total of 12503 articles were detected across the four databases, and after removing the duplicates and non-eligible articles, 91 studies were included in this systematic review: 50 observational studies (all cross-sectional) ^36–85^, 9 studies reported the acute effects of PA (e.g., following a marathon, rowing, etc.) on microbiome ^23,86–93^, and 32 studies reported the chronic effects of PA on microbiome (17 non-RCT, 13 RCT and 2 randomized controlled cross-over trials) ^94–125^. Of the 50 cross-sectional studies, 8 were eligible (based on availability of microbiome diversity data and healthy participants) for the first meta-analysis comparing groups of high and low PA levels in non-athletes ^44,66,69,70,78,81,83,85^, and 11 were included in the second meta-analysis comparing athletes *vs*. non-athletes ^47,49,104,51,56,60,74,76,78,79,81^. Of the 32 intervention studies, 7 were selected for the third meta-analysis, to evaluate the chronic effects of PA on microbiome alpha-diversity^96,97,99,109,112,119,124^.

**Fig. 1.**
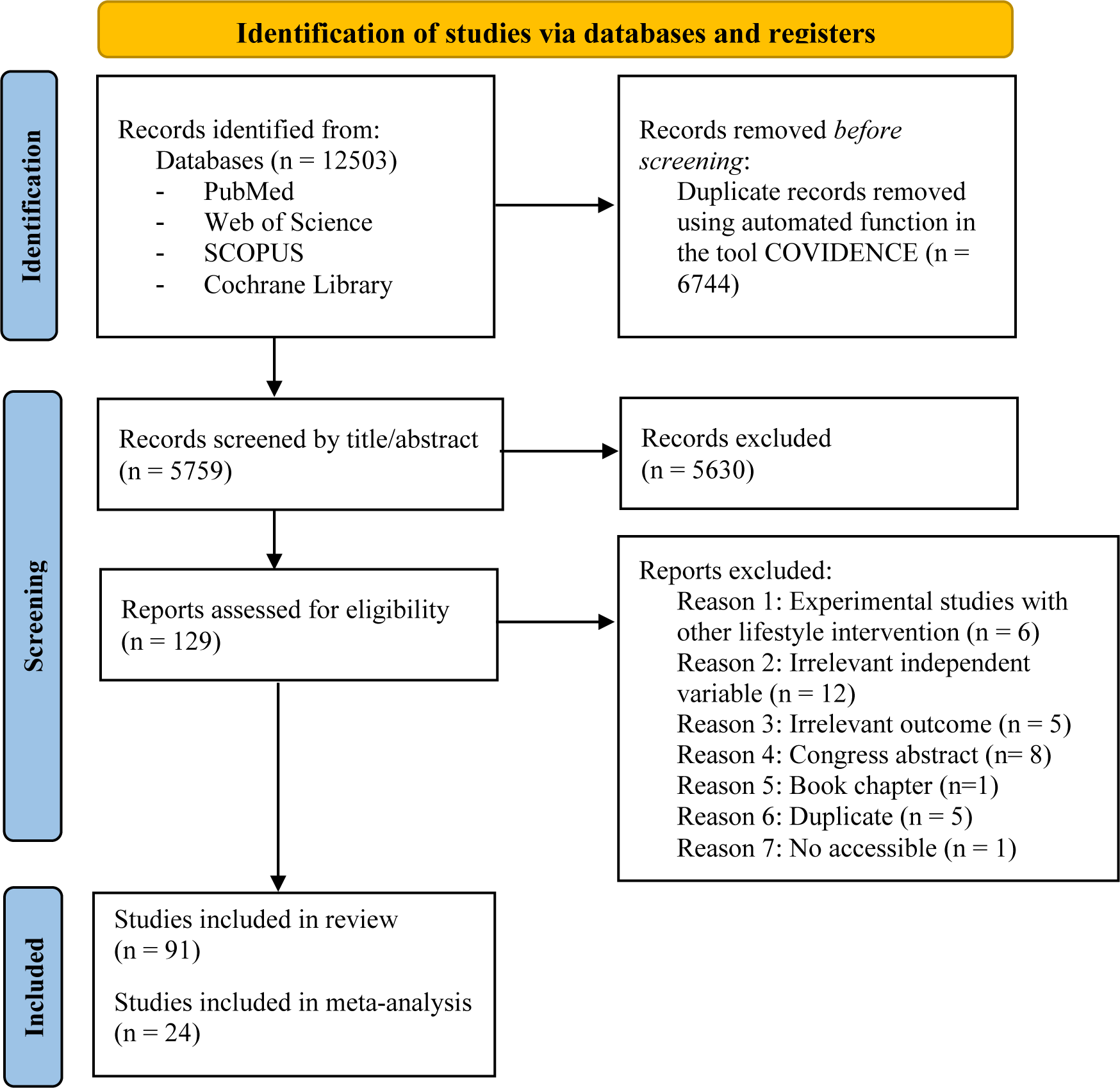
Search process according to the Preferred Reporting Items for Systematic Reviews and Meta-Analyses (PRISMA) 2020 flow diagram

Seventeen studies reported significant associations between PA ^38,41,72,83–85,43,44,59,63,65–68^ or SB ^48,64,70^ and microbial diversity (i.e., alpha- and/or beta-diversity), and 19 studies found significant differences in the relative abundance of specific bacteria in active *vs*. inactive participants ^36,38,63,66,68–71,78,84,85,39,41,42,44–46,48,59^ (**Table 2**). Sixteen studies found significant differences in microbial diversity ^47,49,62,74–76,79,80,51,52,54–58,61^ and 13 in the abundance of specific microbial taxa ^47,52,78,79,104,53,55–58,74–76^ between athletes *vs*. non-athletes, professional *vs*. amateur or athletes from different sports. Three studies detected significant differences in alpha-diversity ^89,90,92^, while 8 studies described significant changes in the relative abundance of certain bacteria after acute PA interventions ^23,86,87,89–93^. Seventeen studies detected significant differences in alpha- and/or beta-diversity ^94,95,115,117–120,123,125,98,101,102,104,108,111,112,114^, and 24 studies described significant changes in the relative abundance of certain bacteria after chronic PA interventions ^94,95,109,111–117,119,121,97,122–125,98,99,101,102,105,107,108^.

**Table 2.**
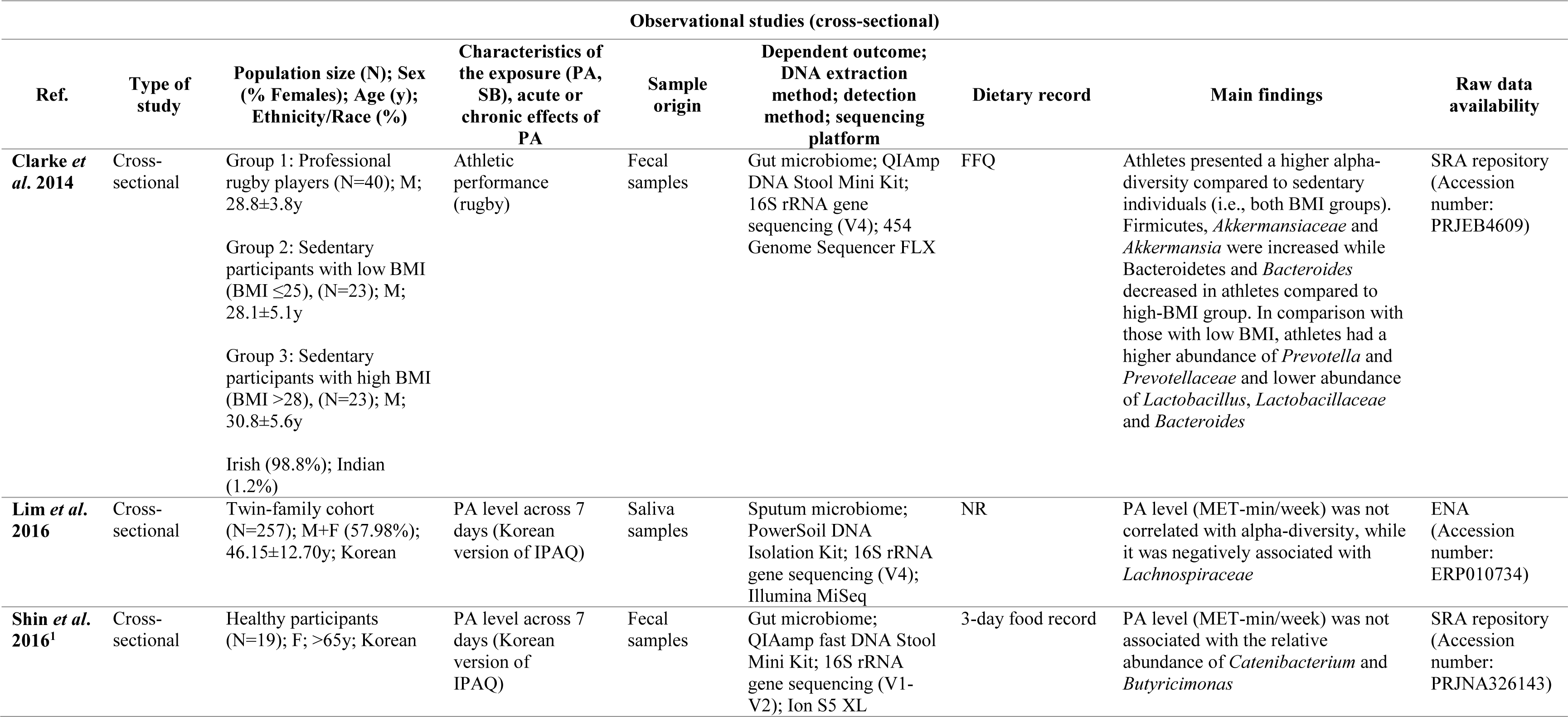
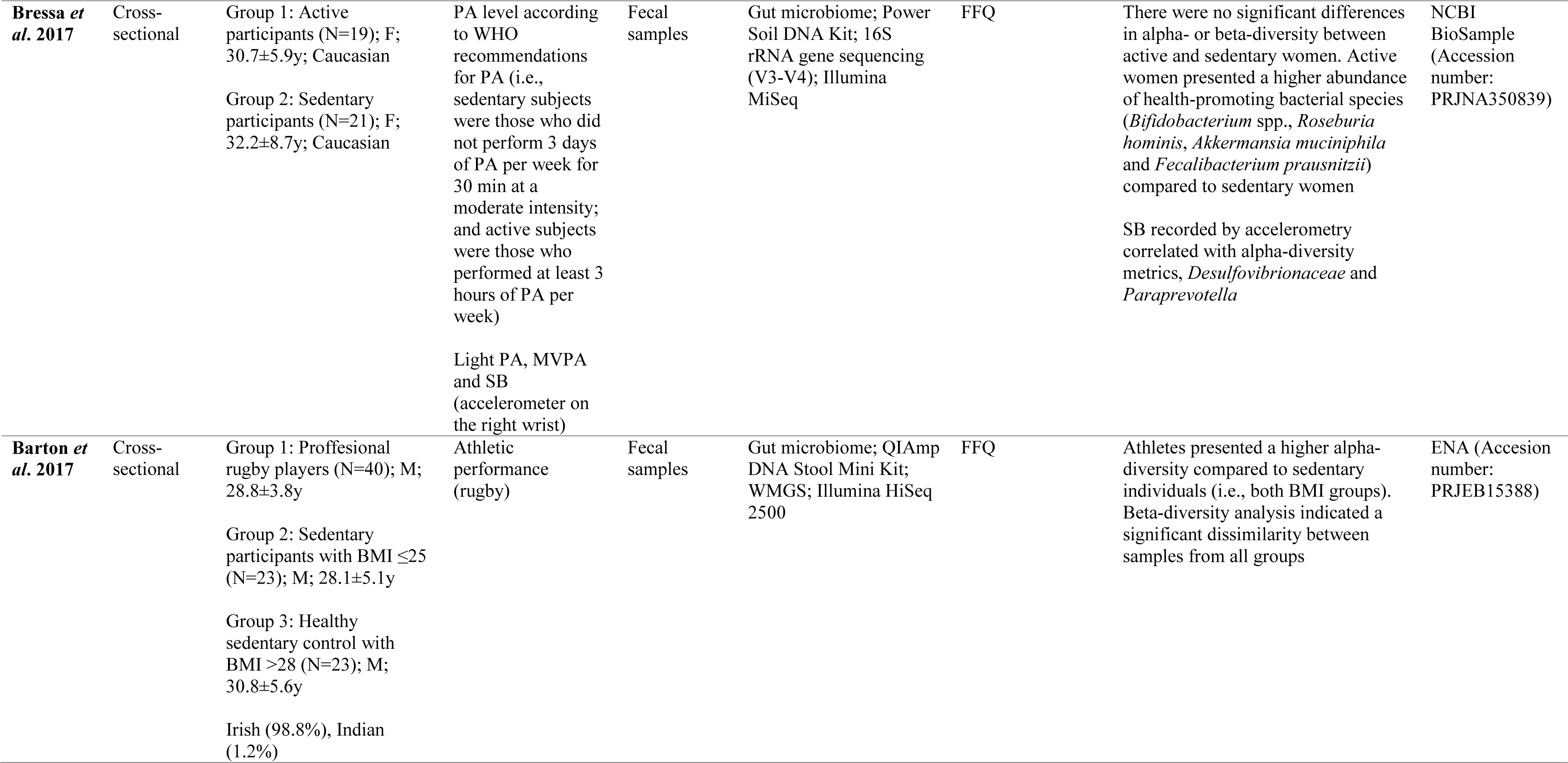
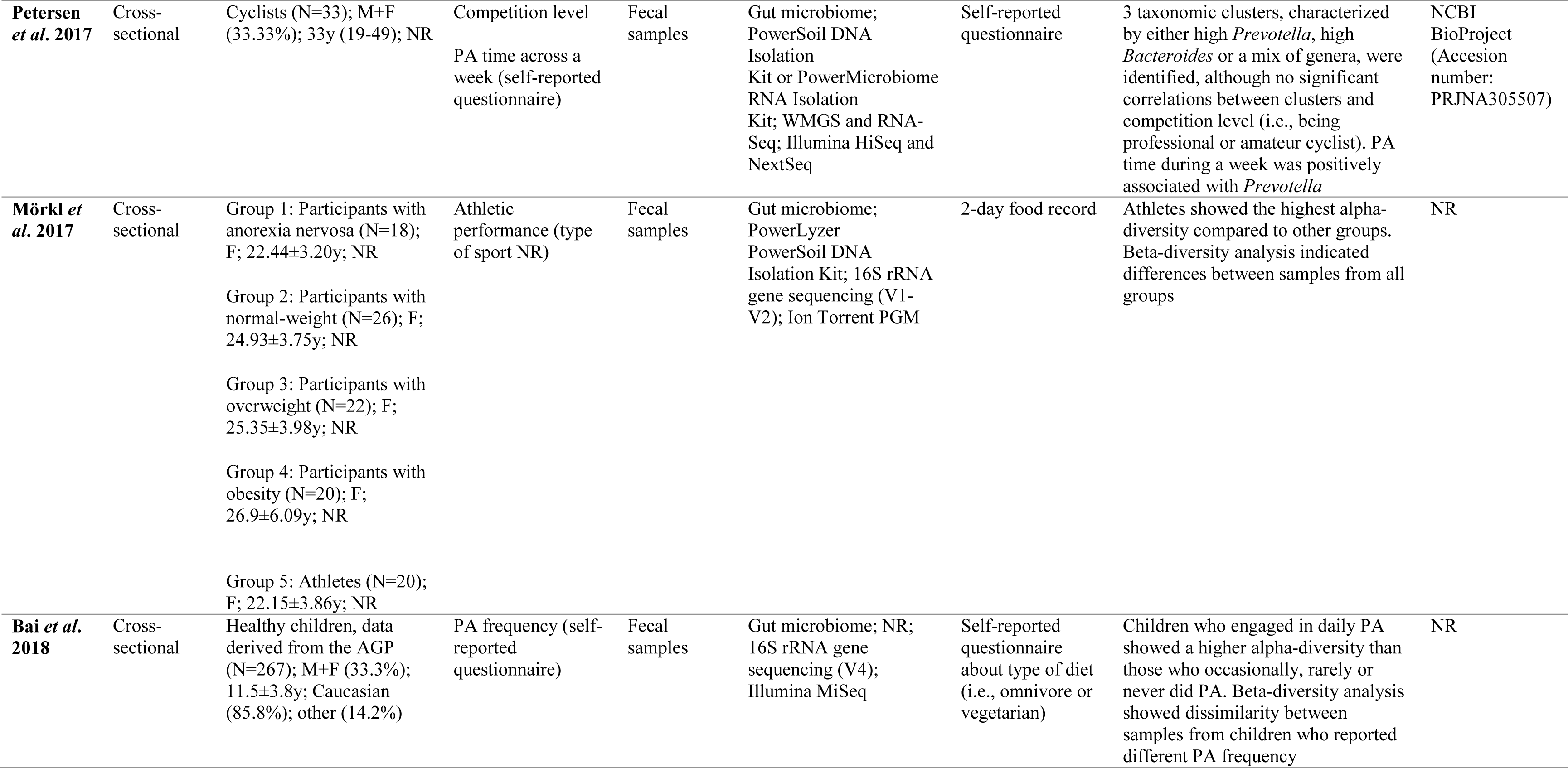
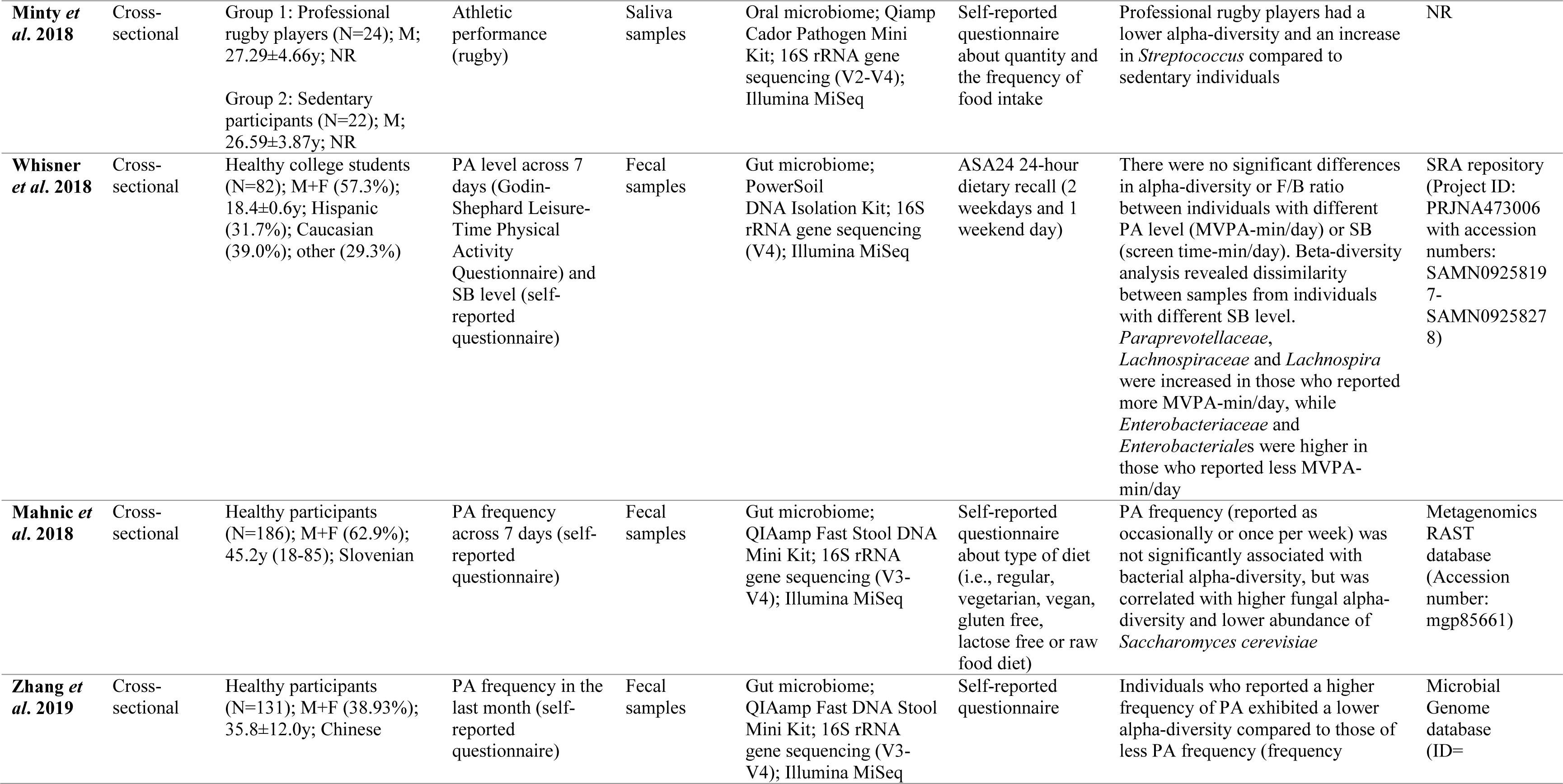
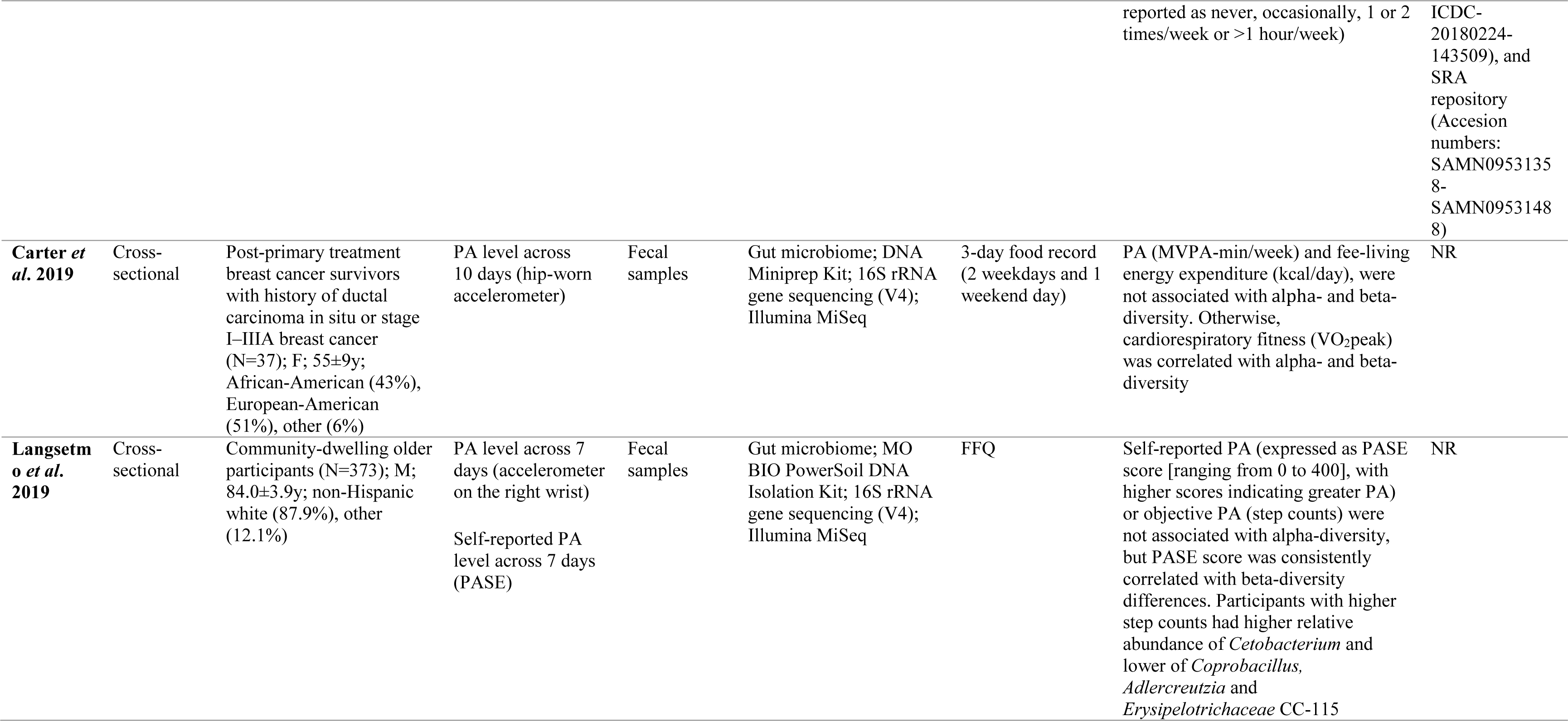
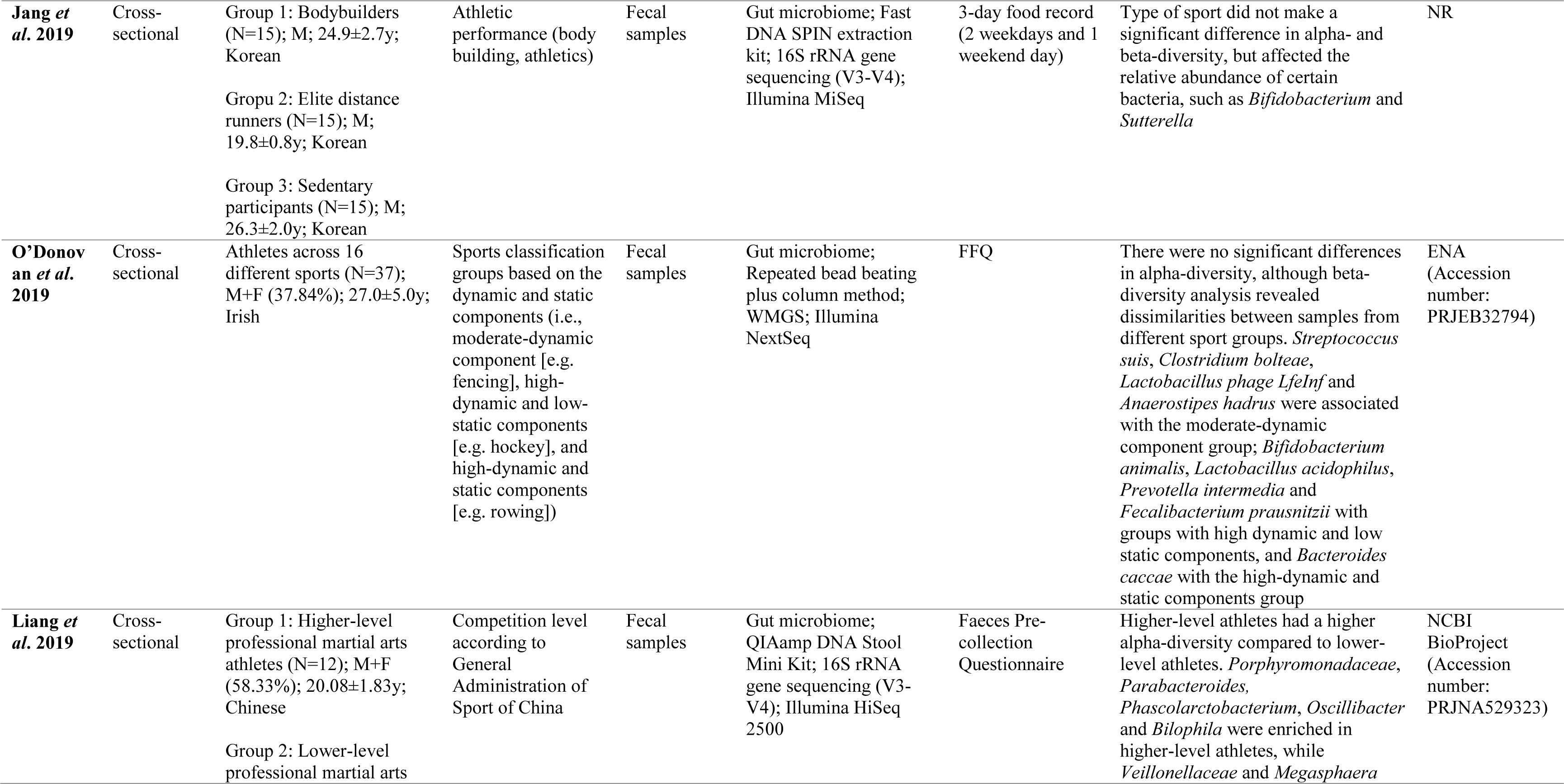
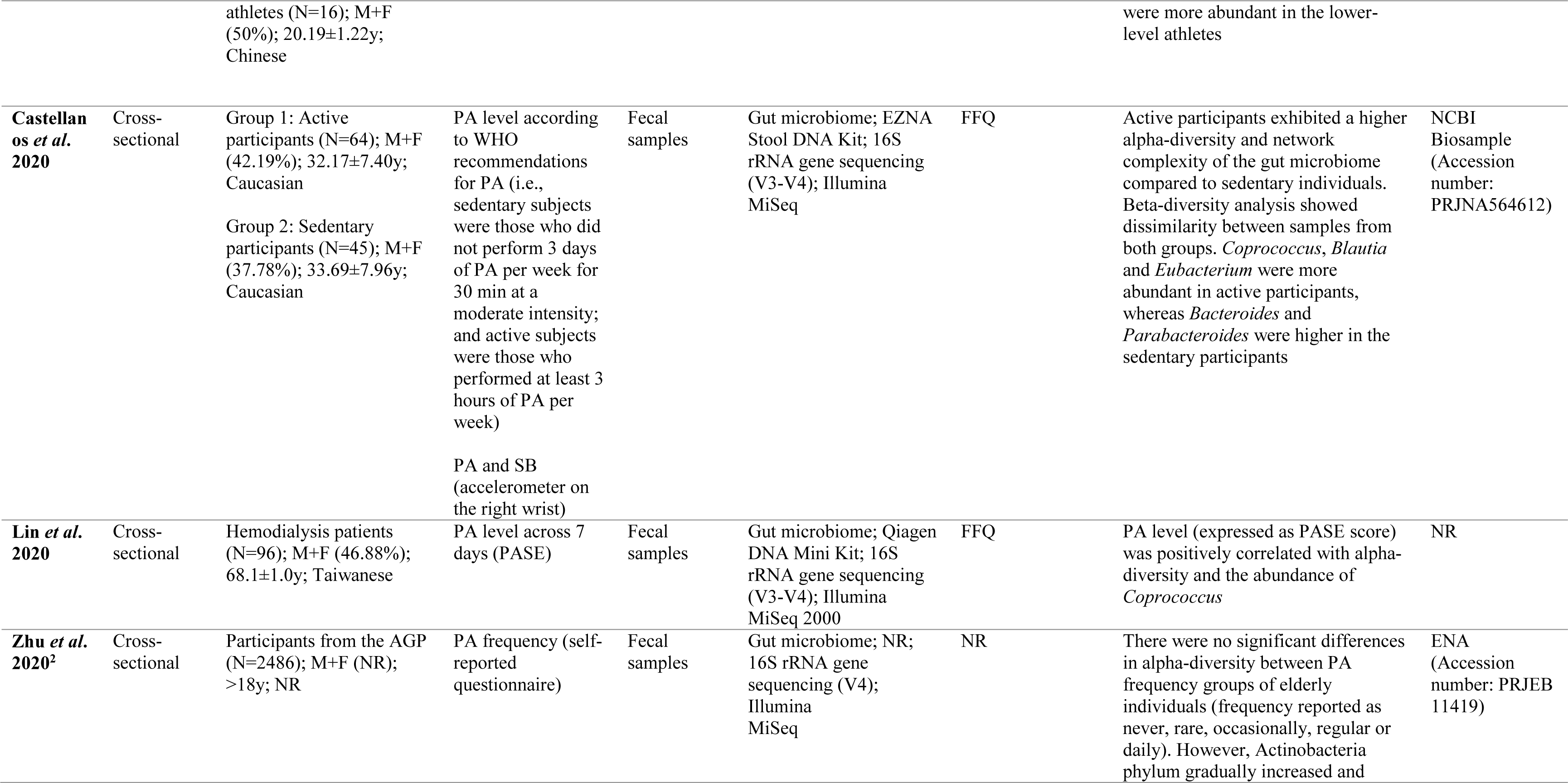
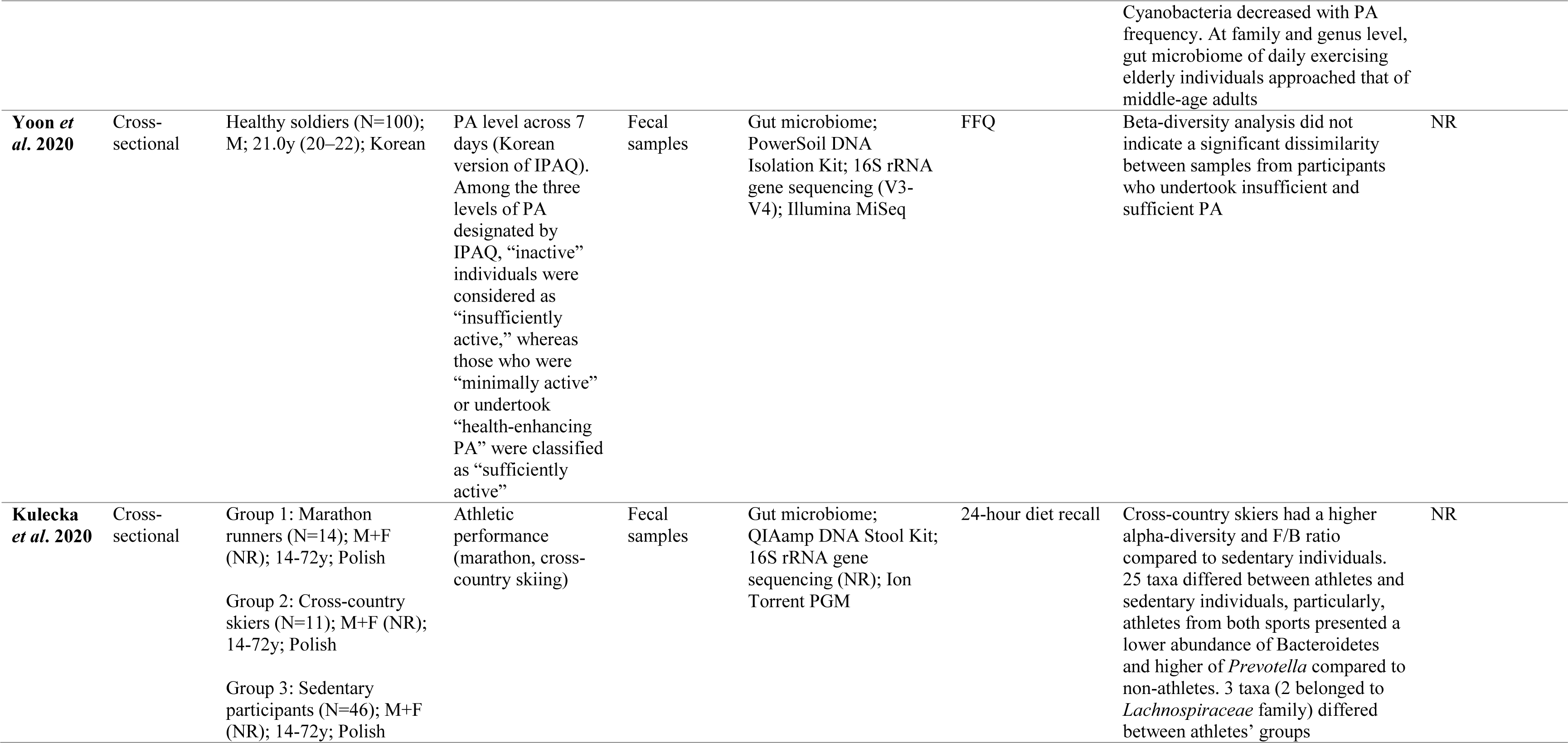
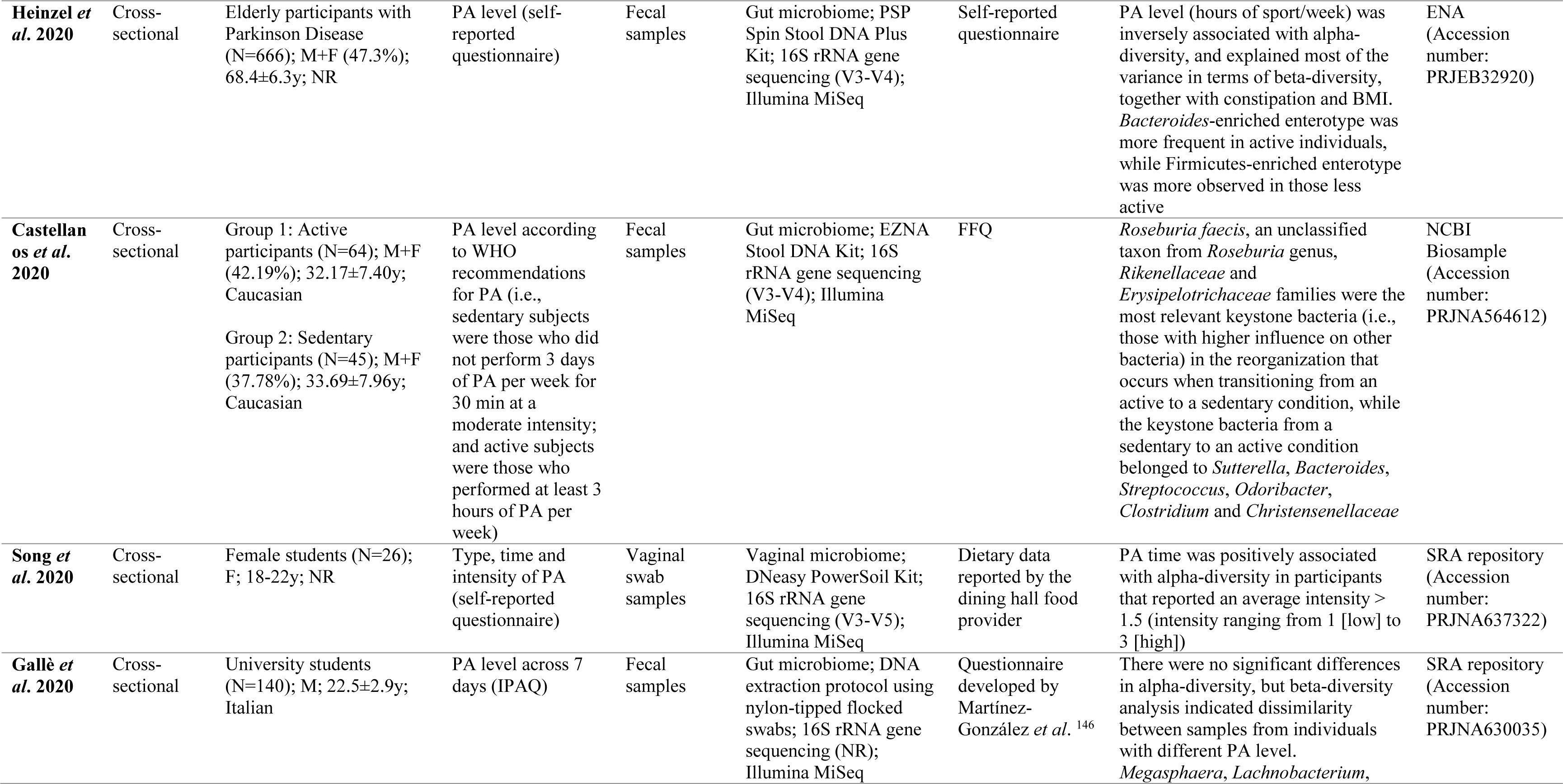
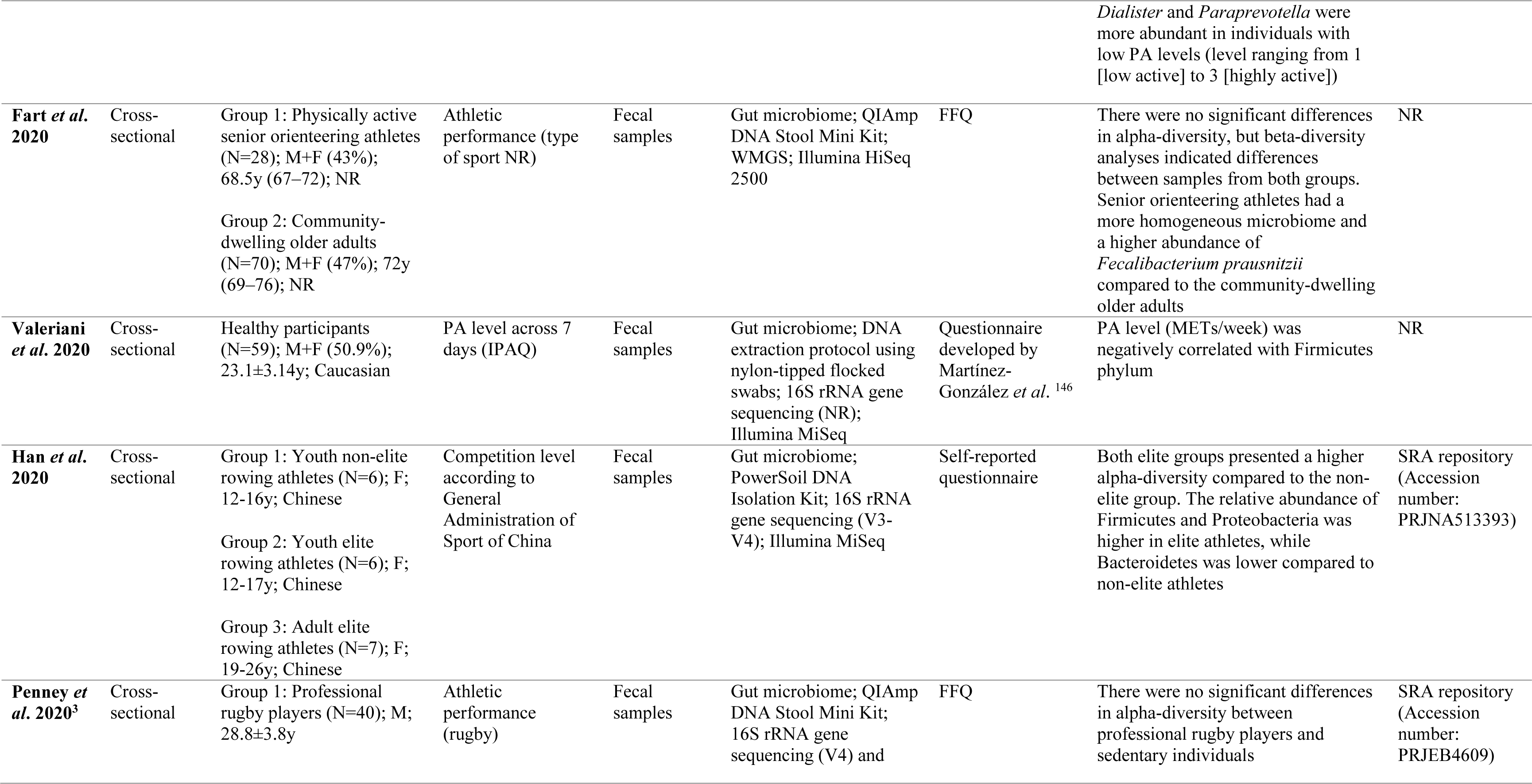
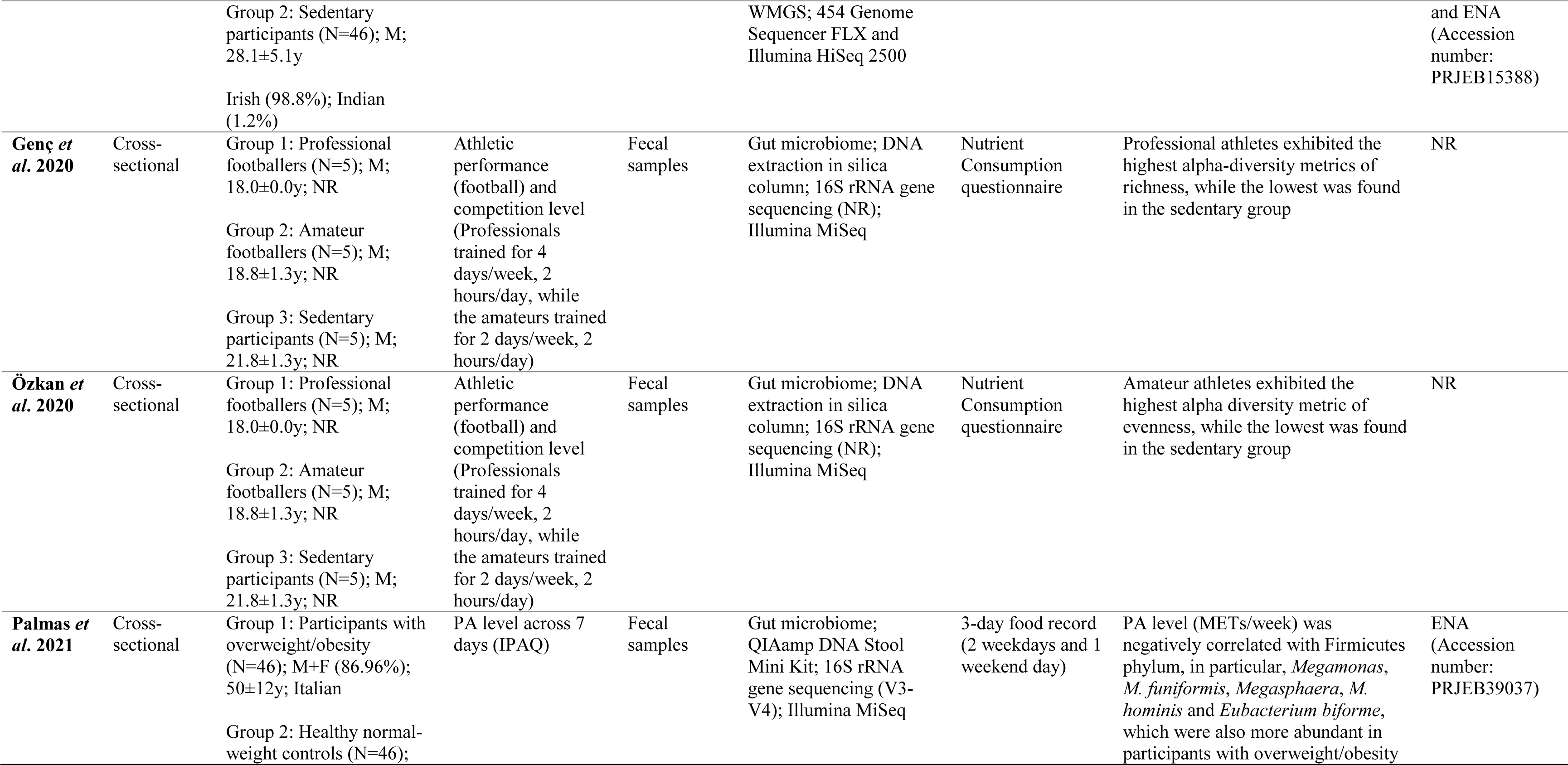
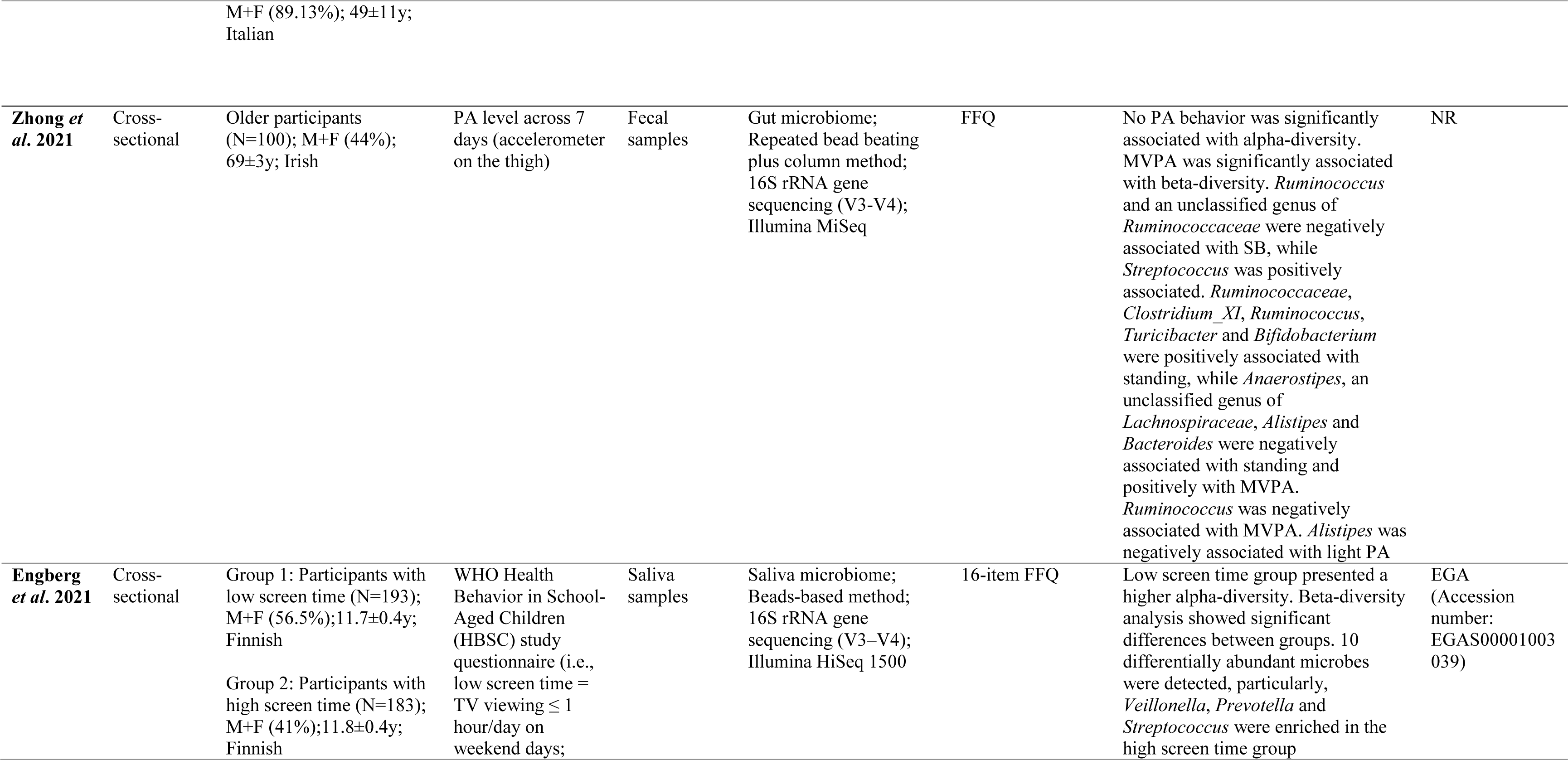
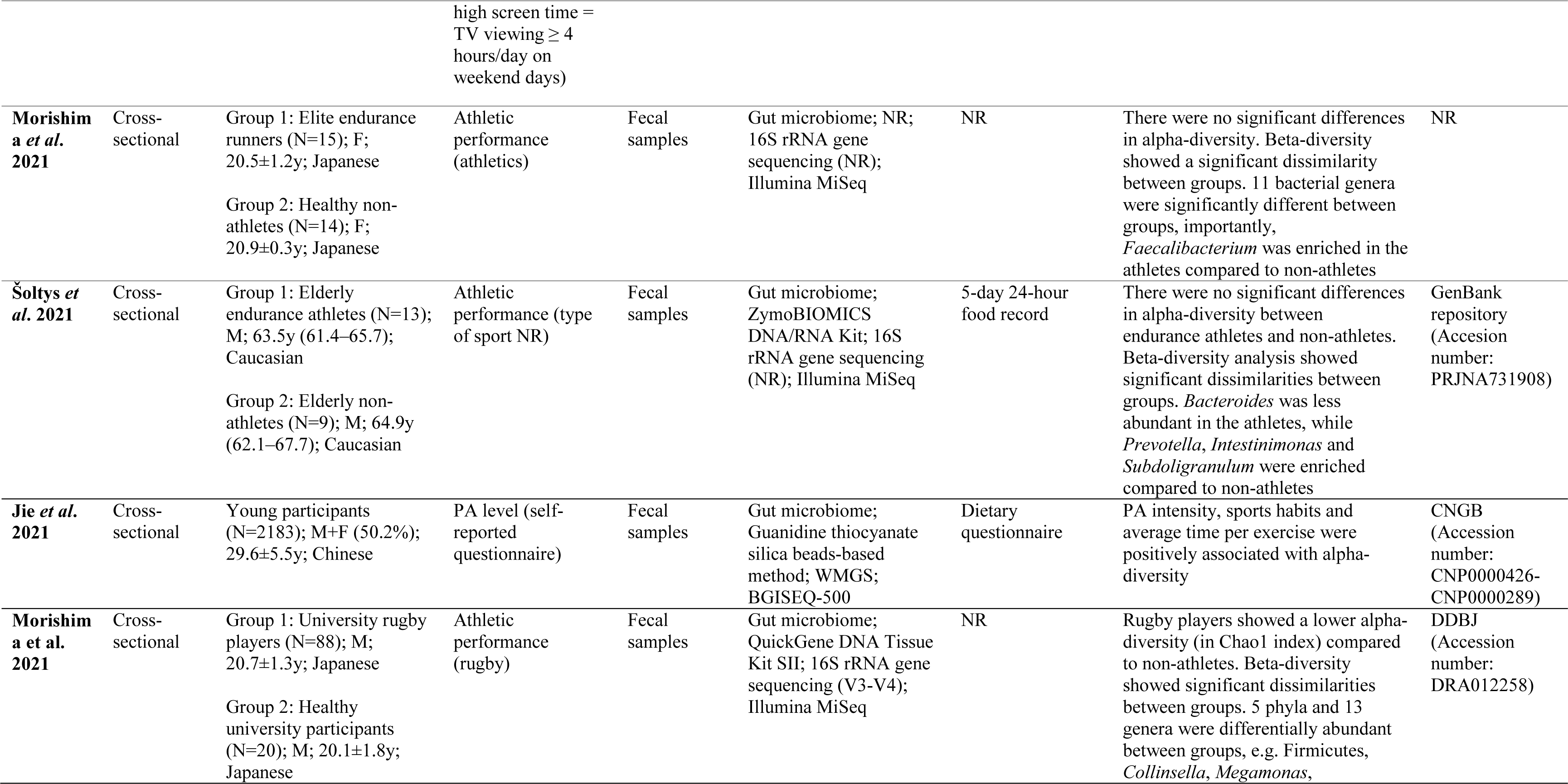
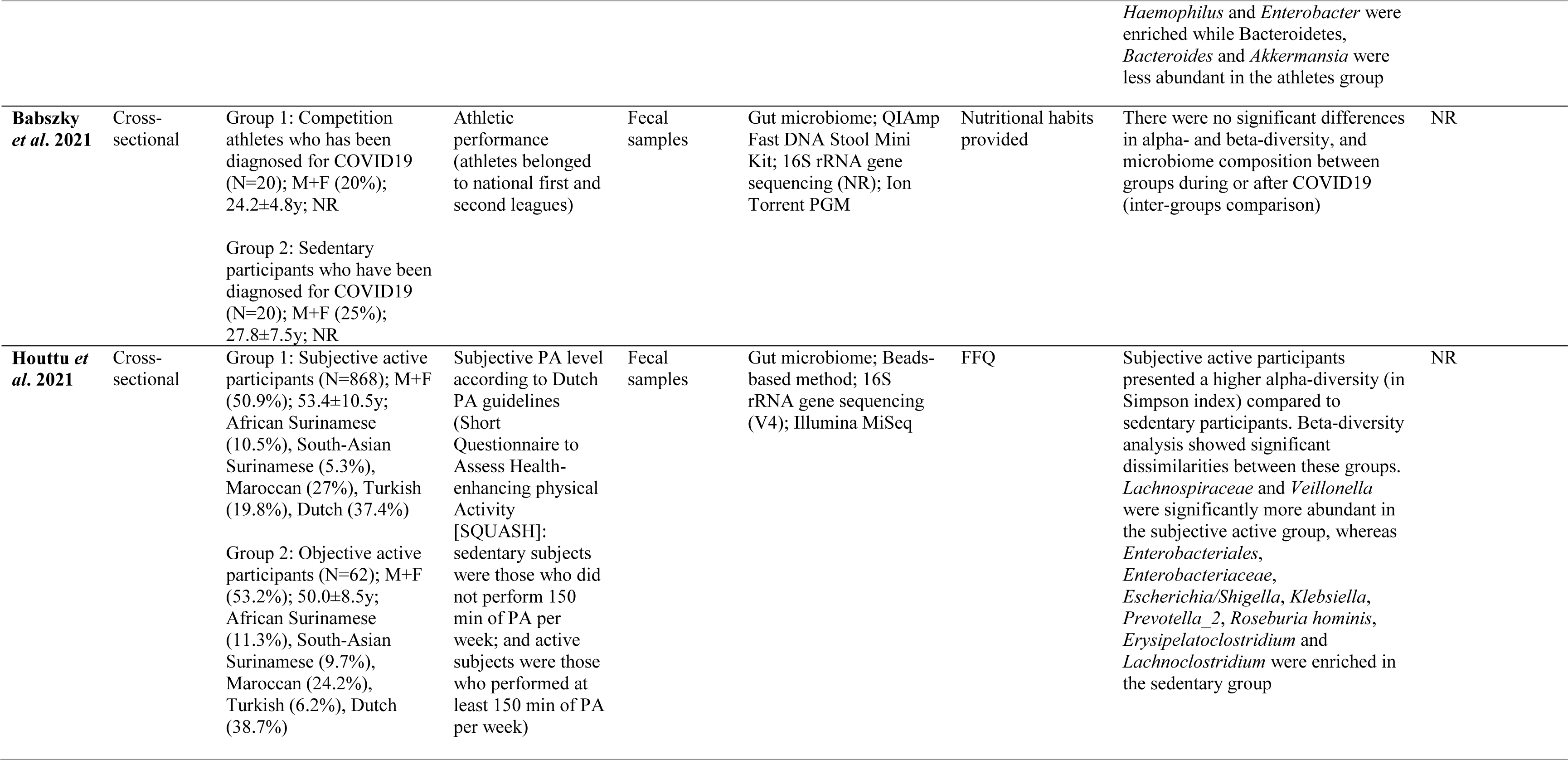
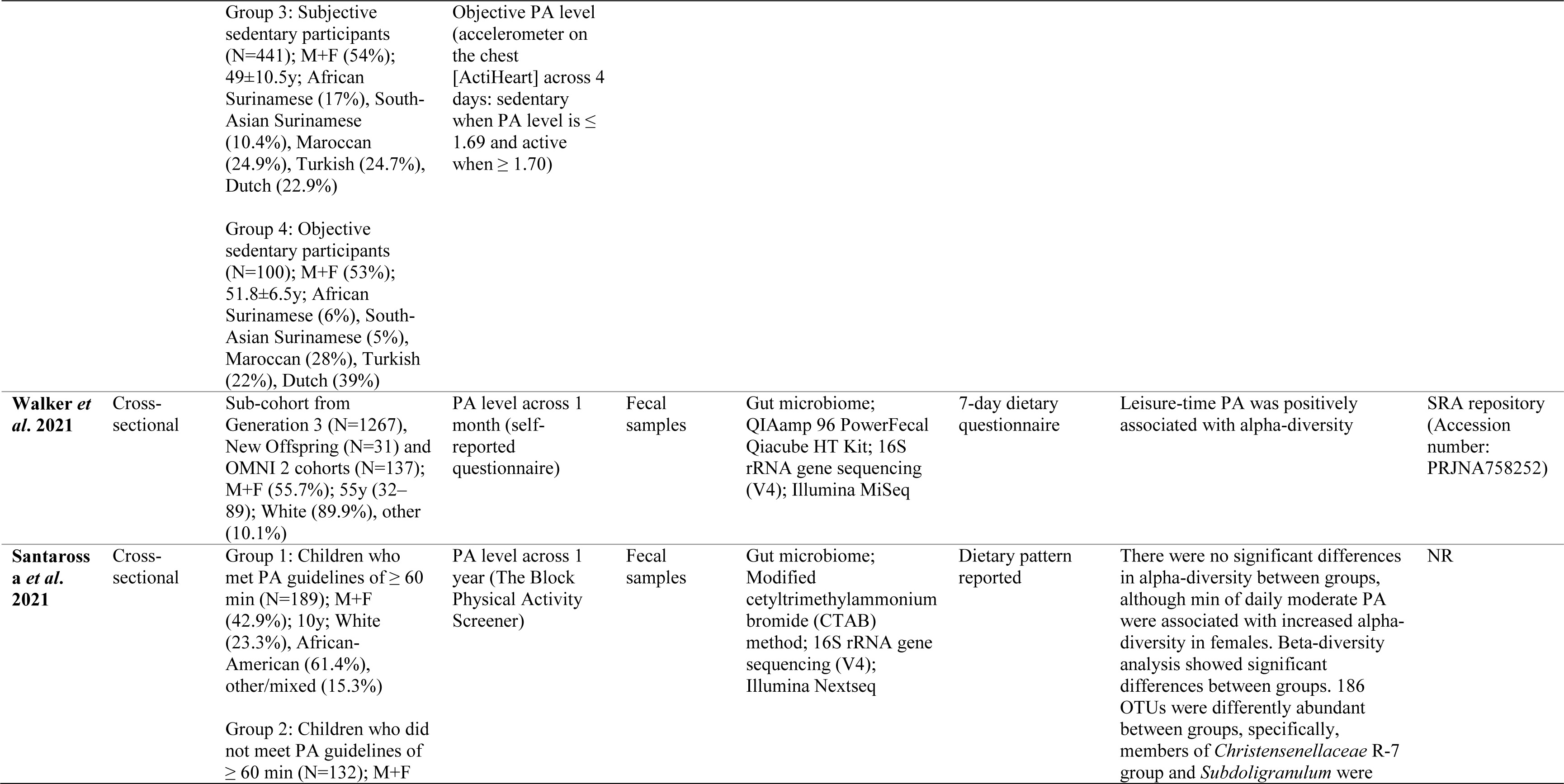
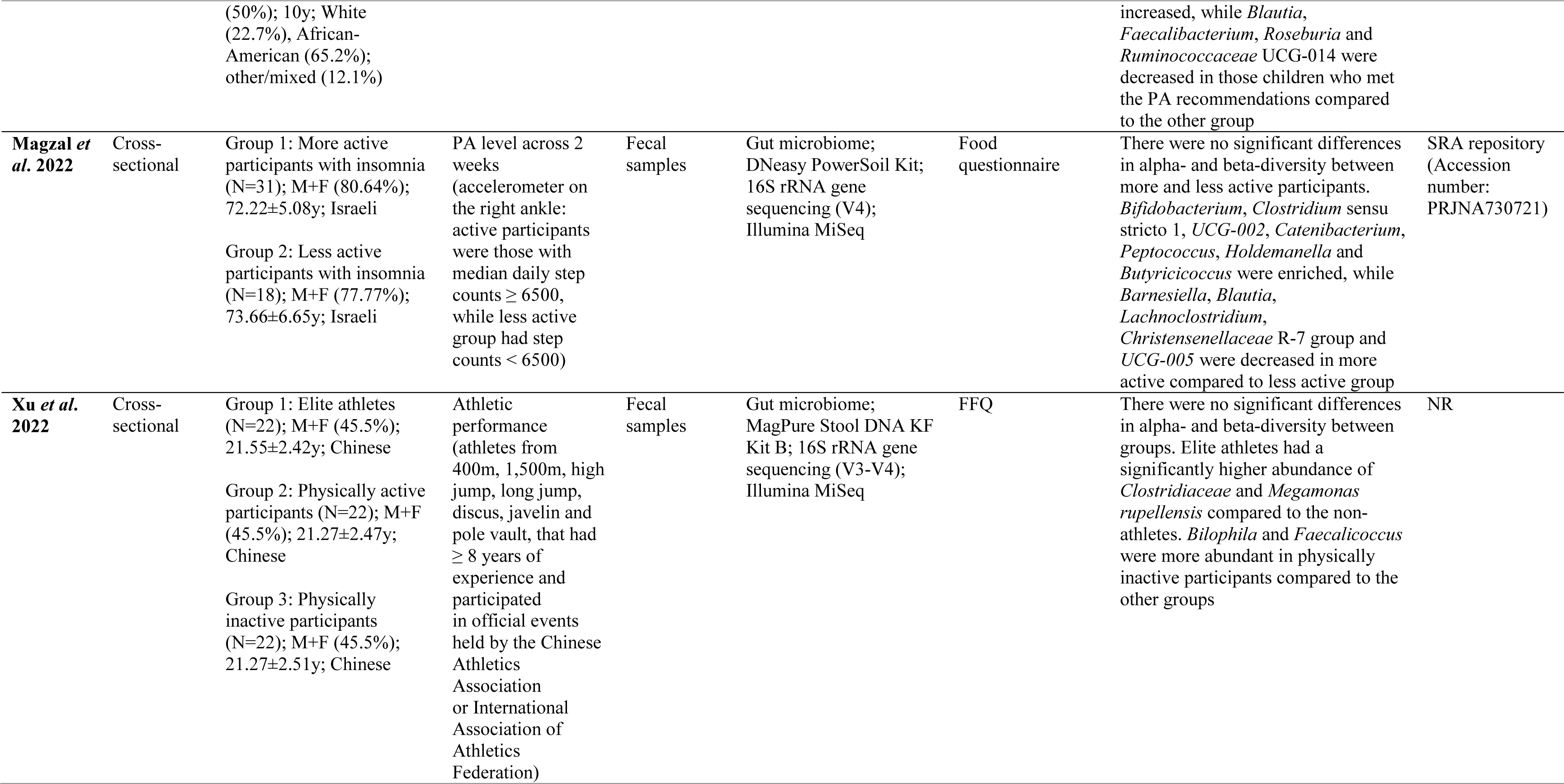
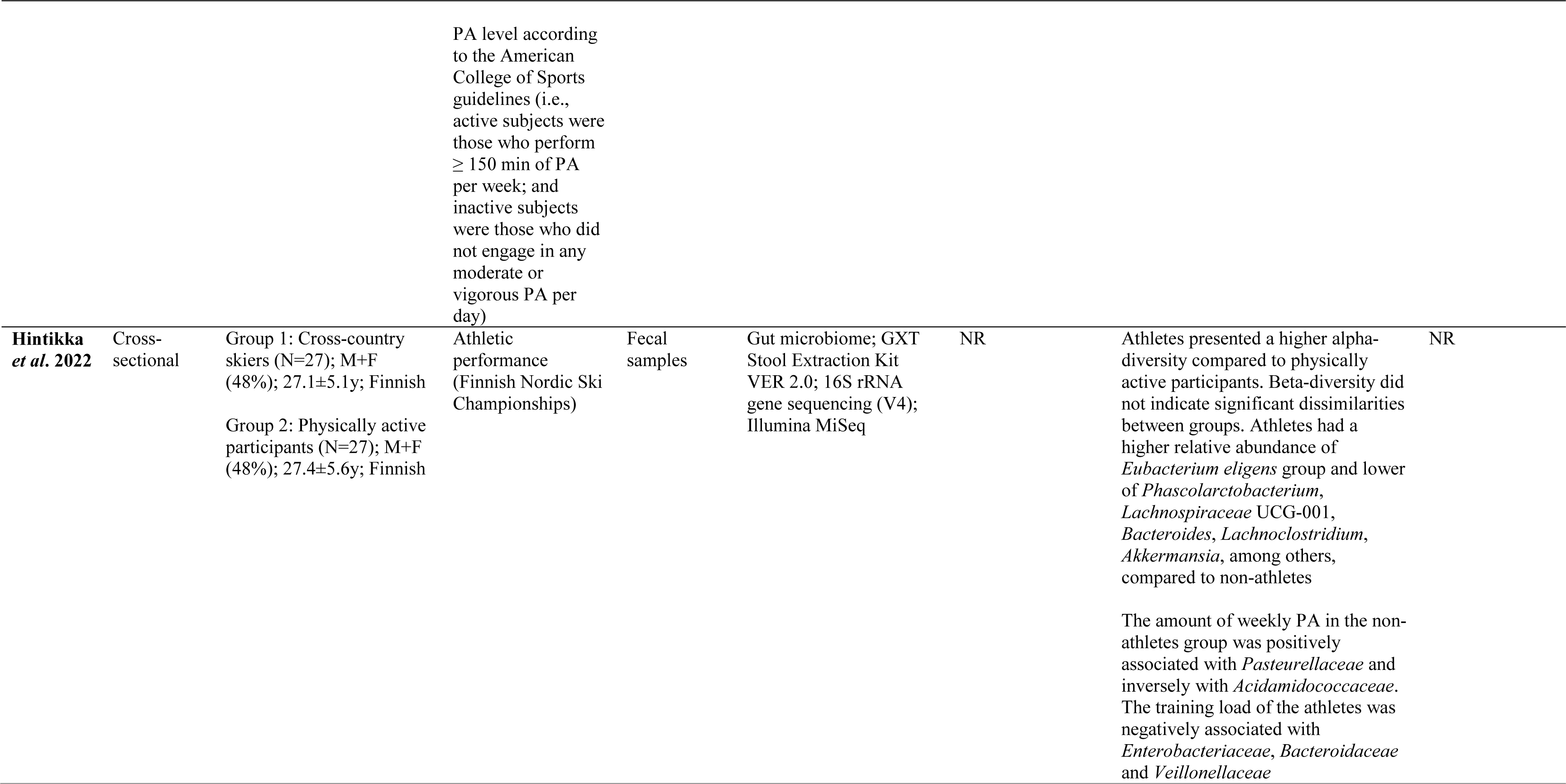
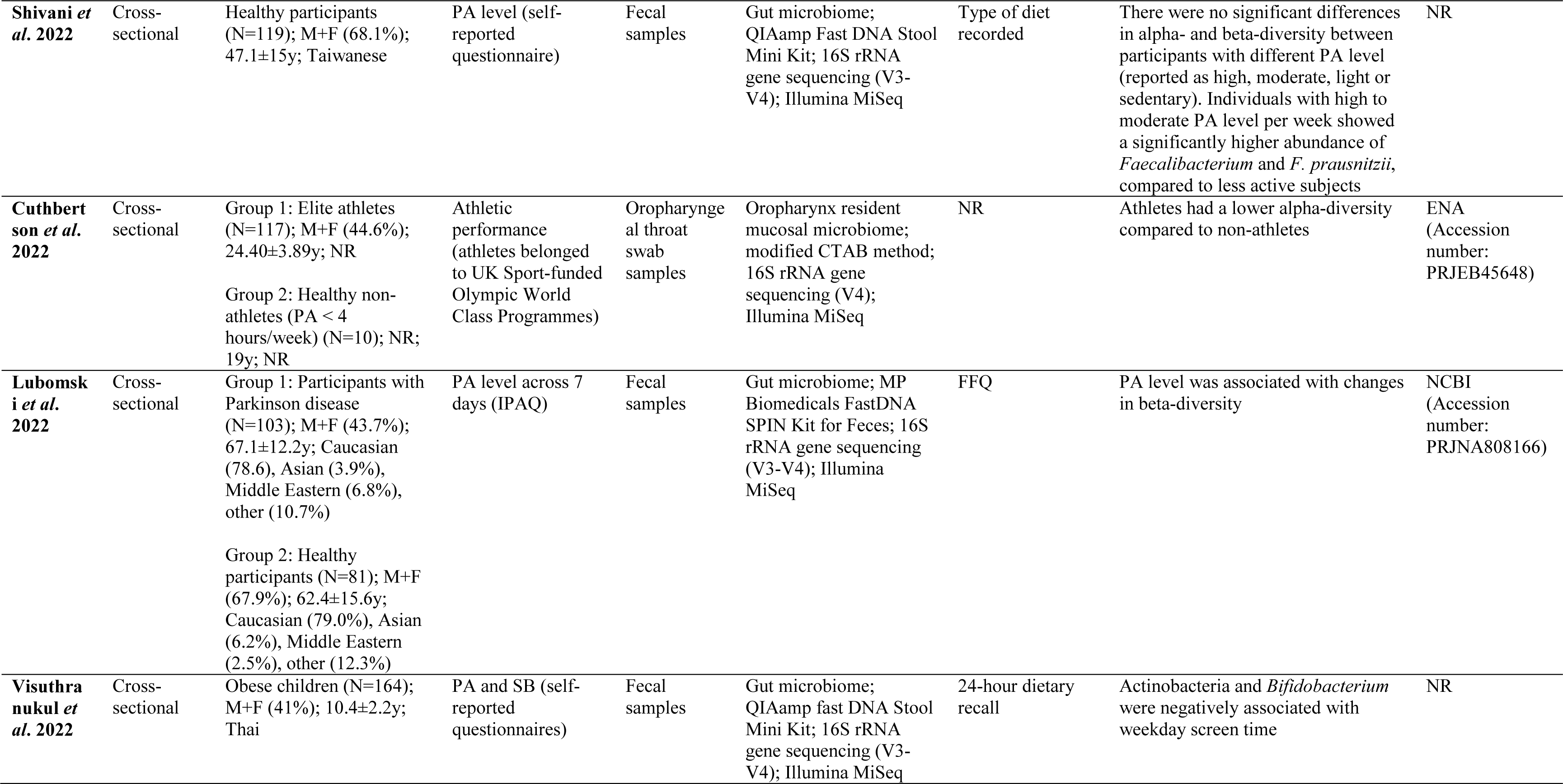
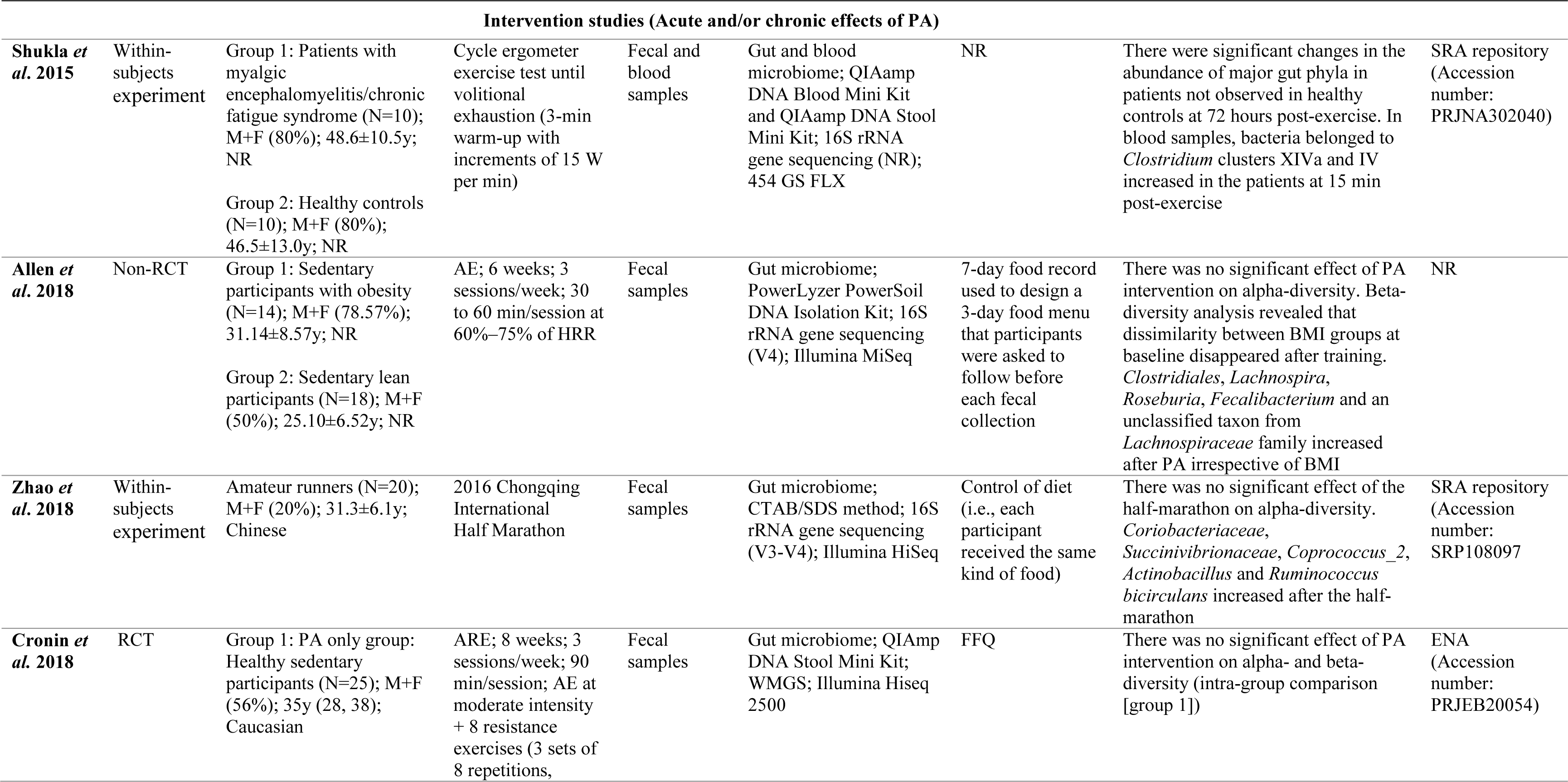
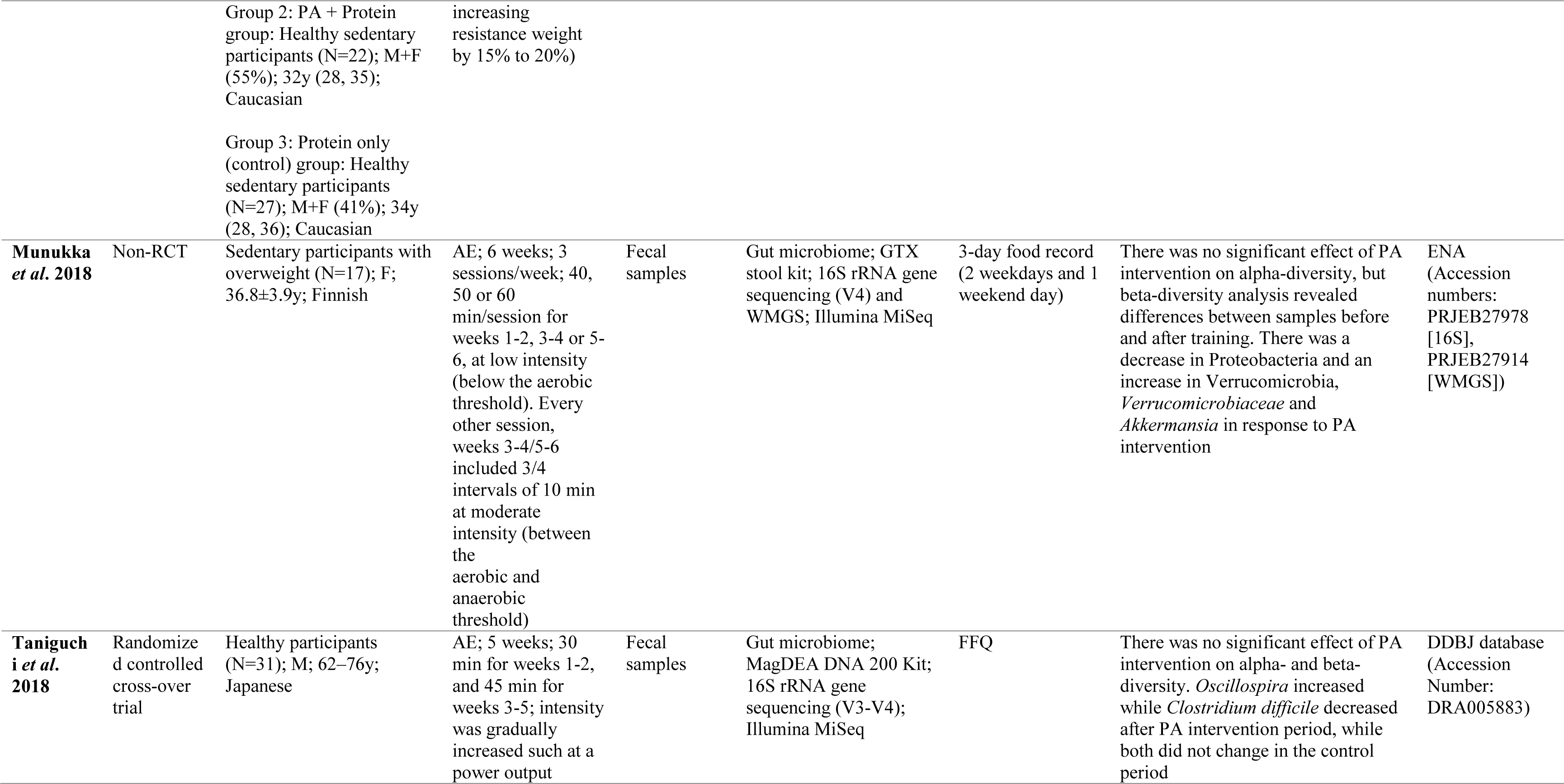
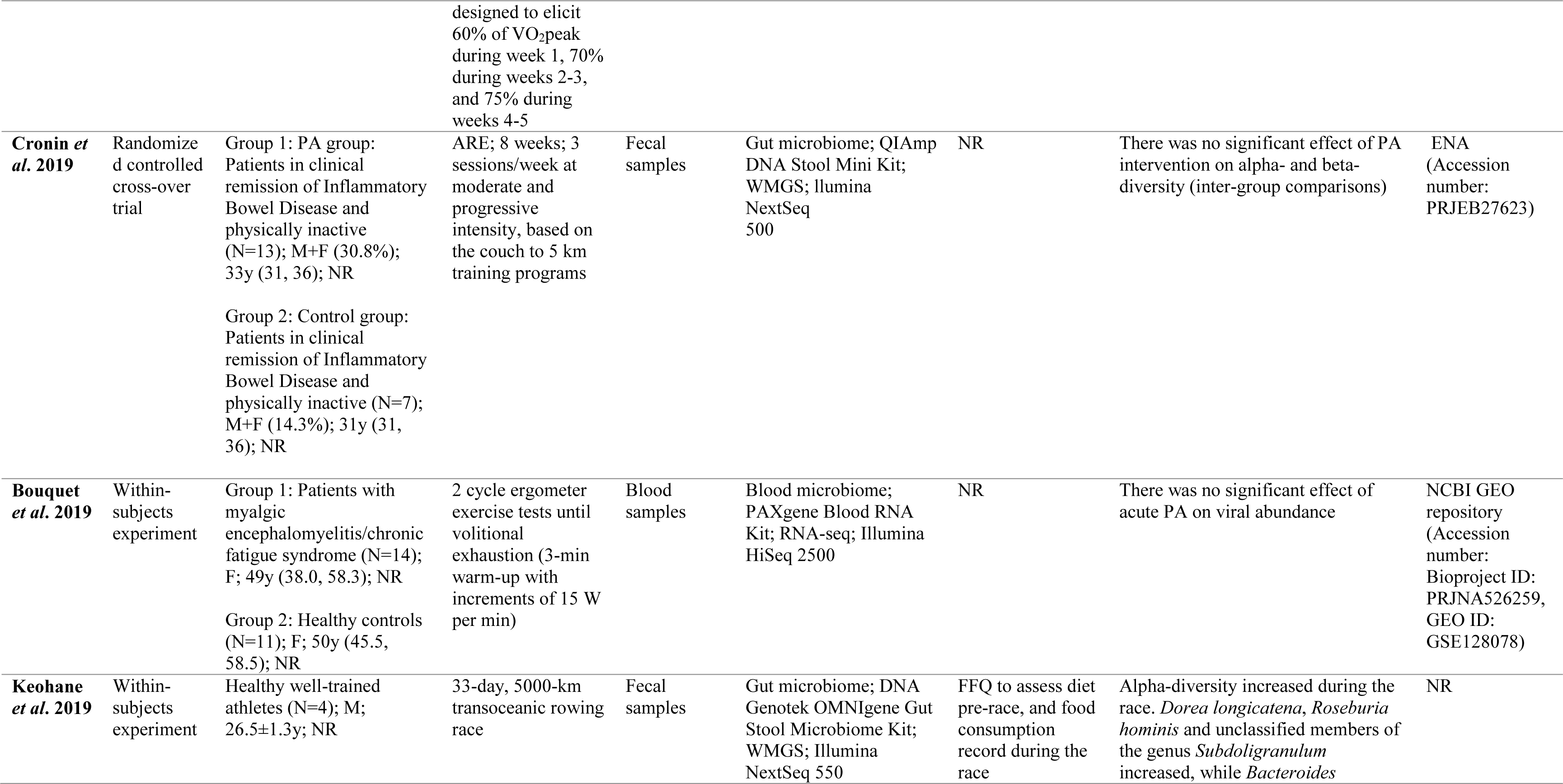
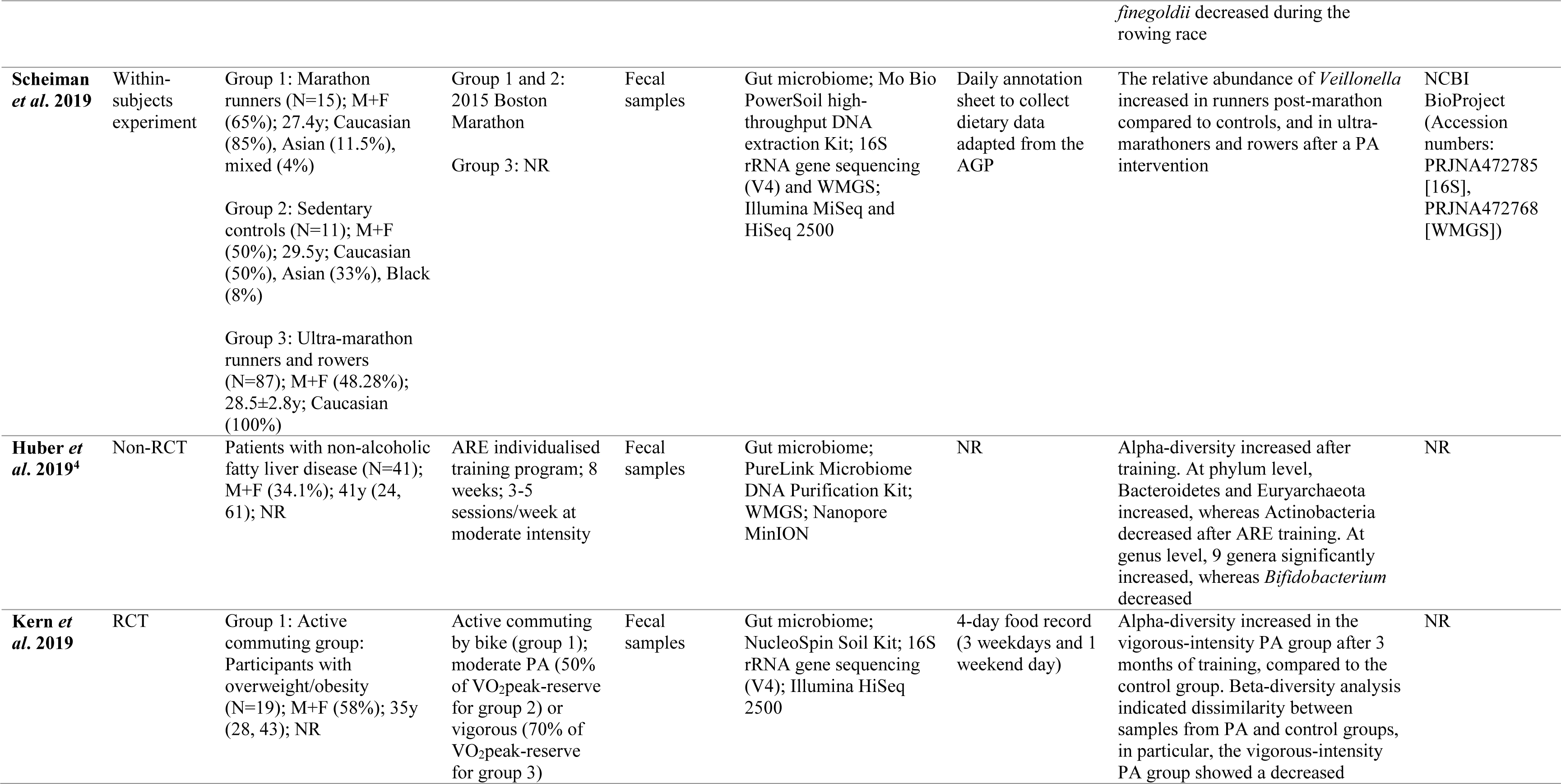
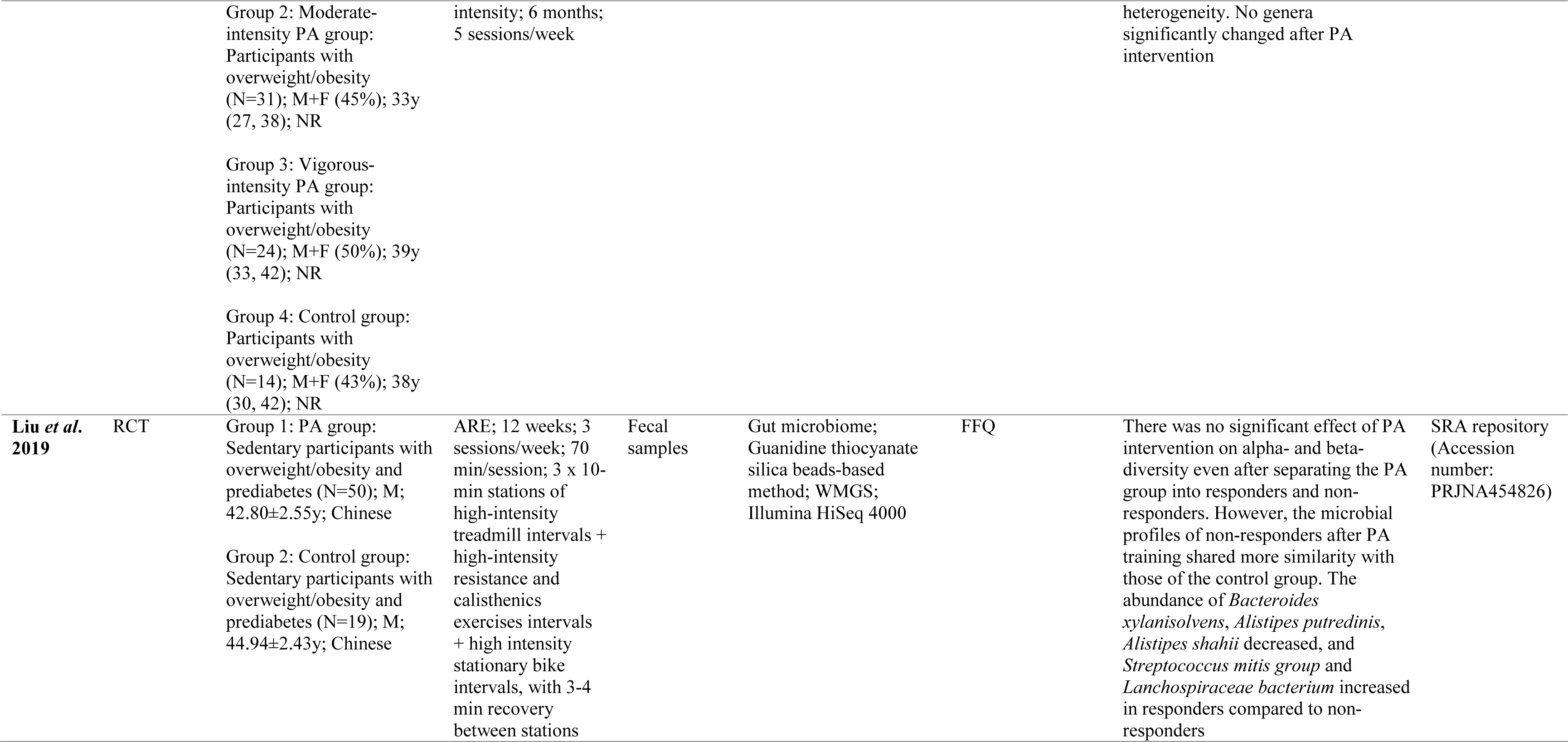
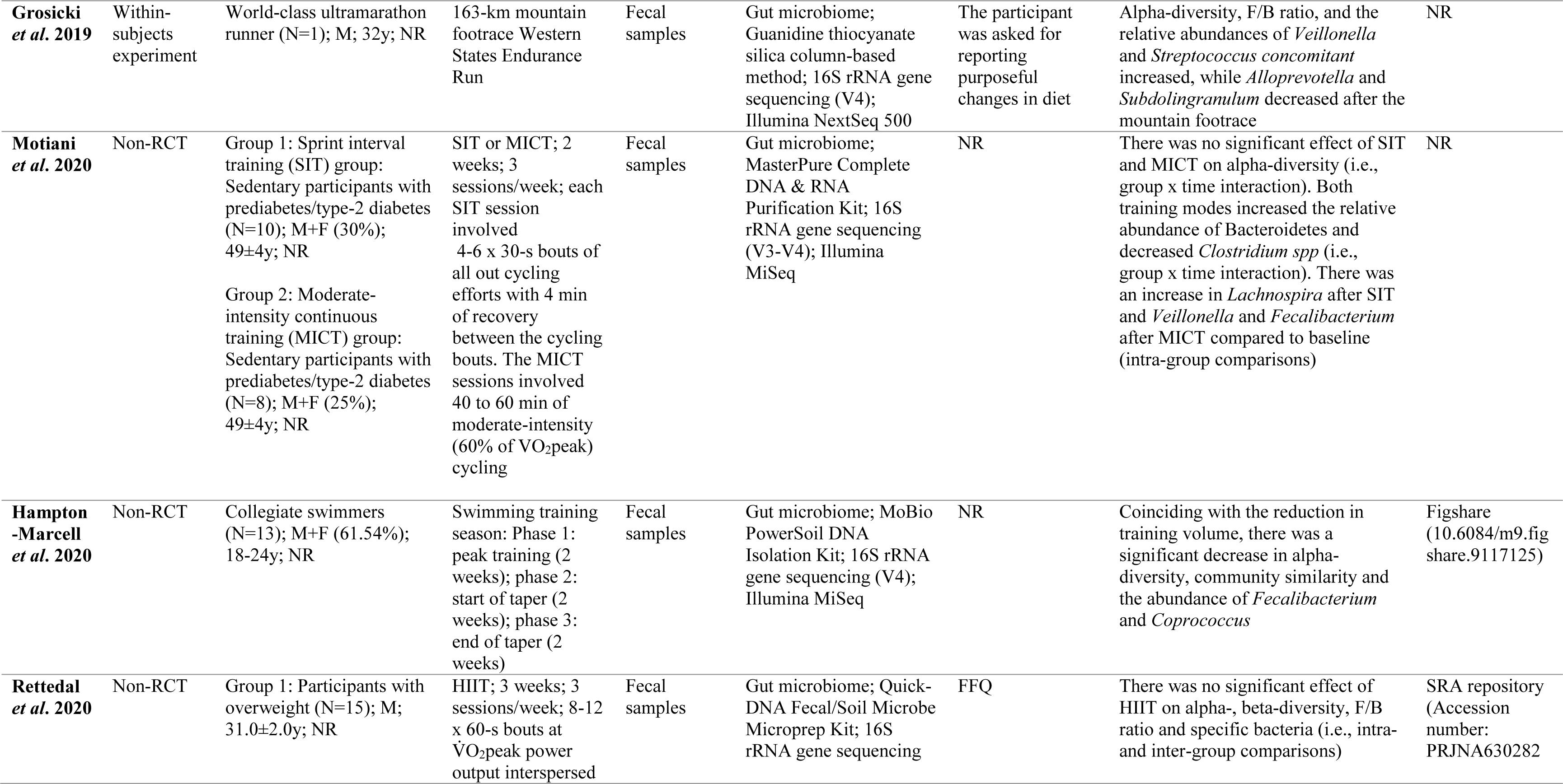
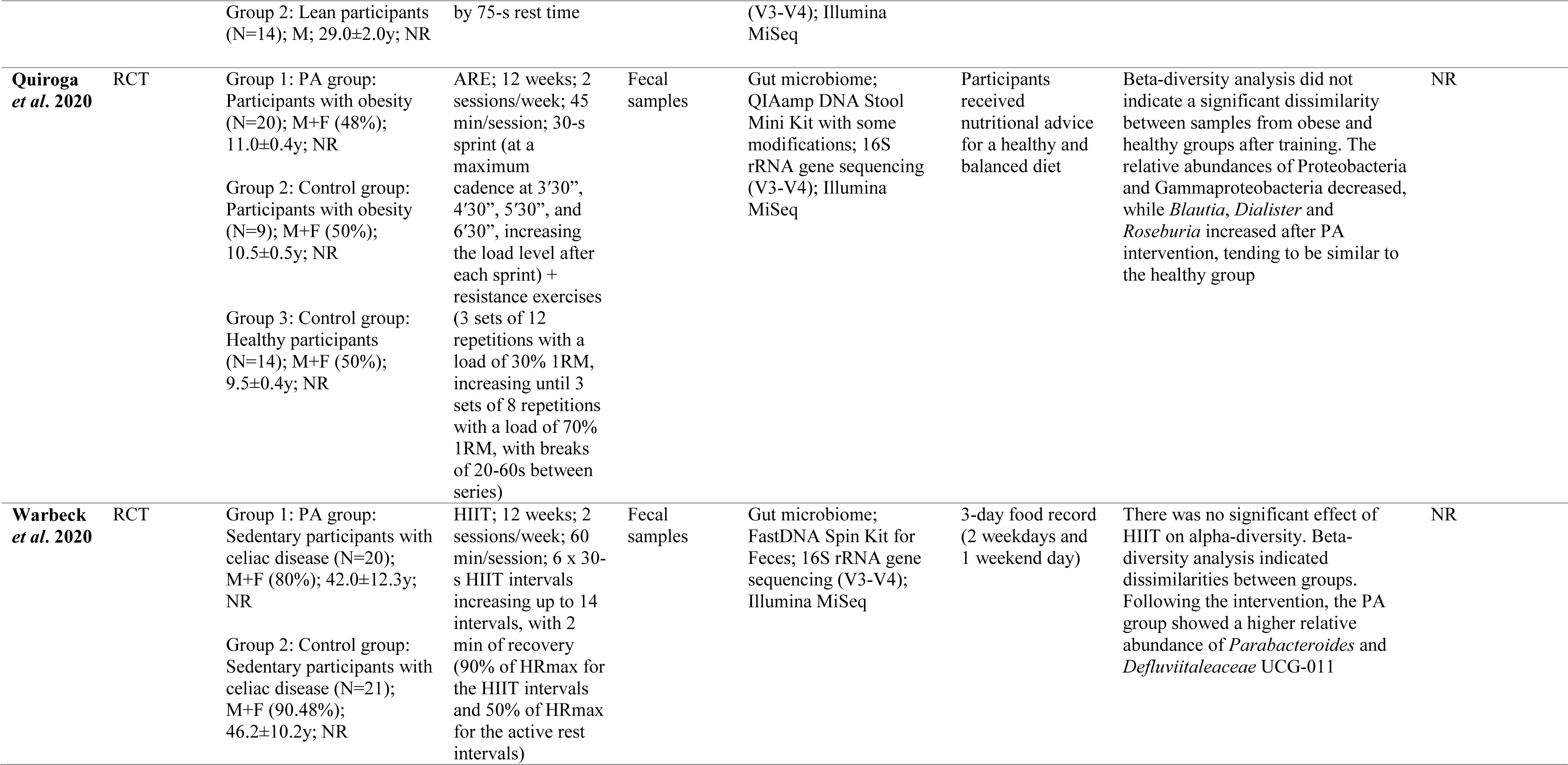
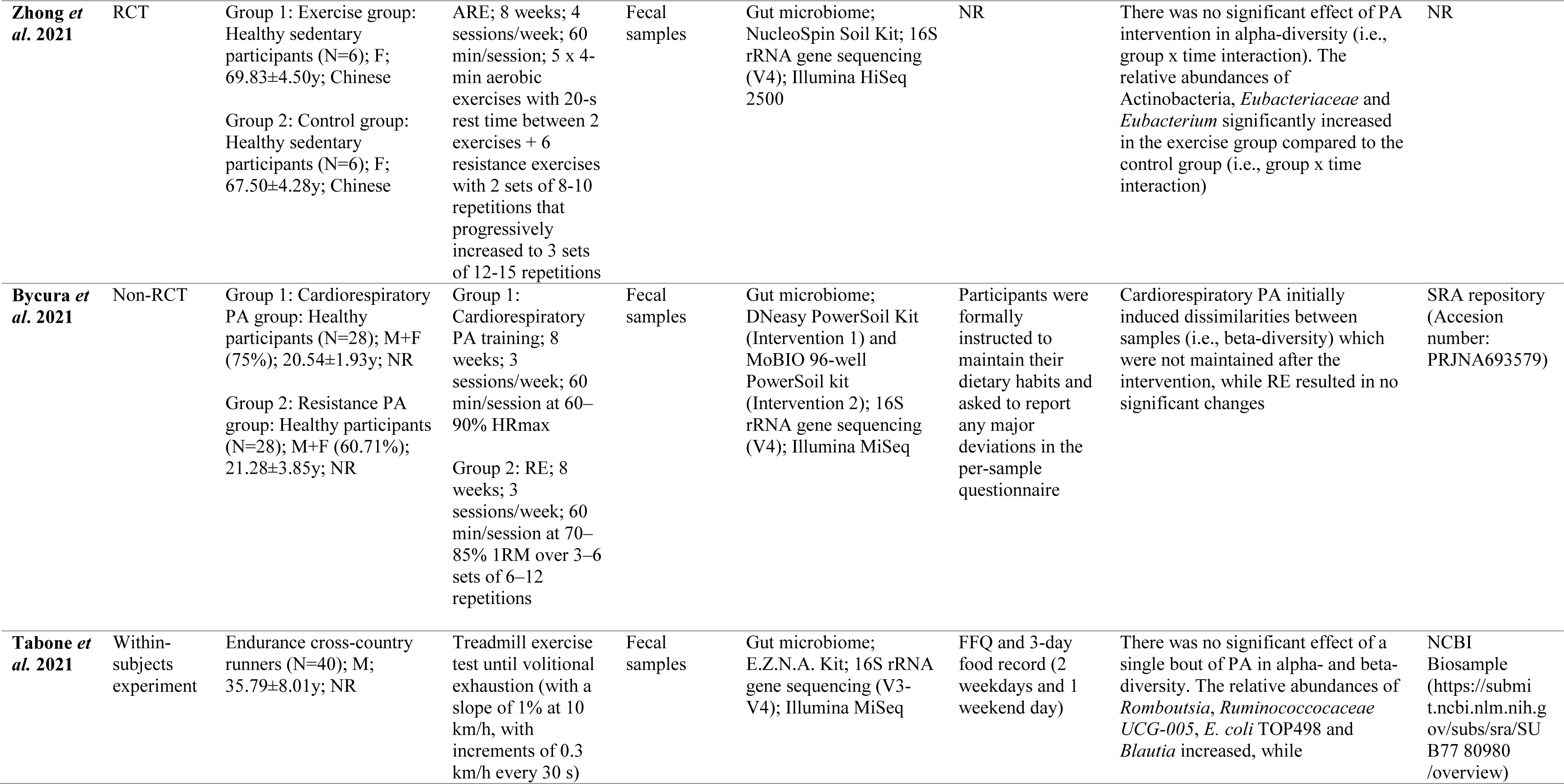
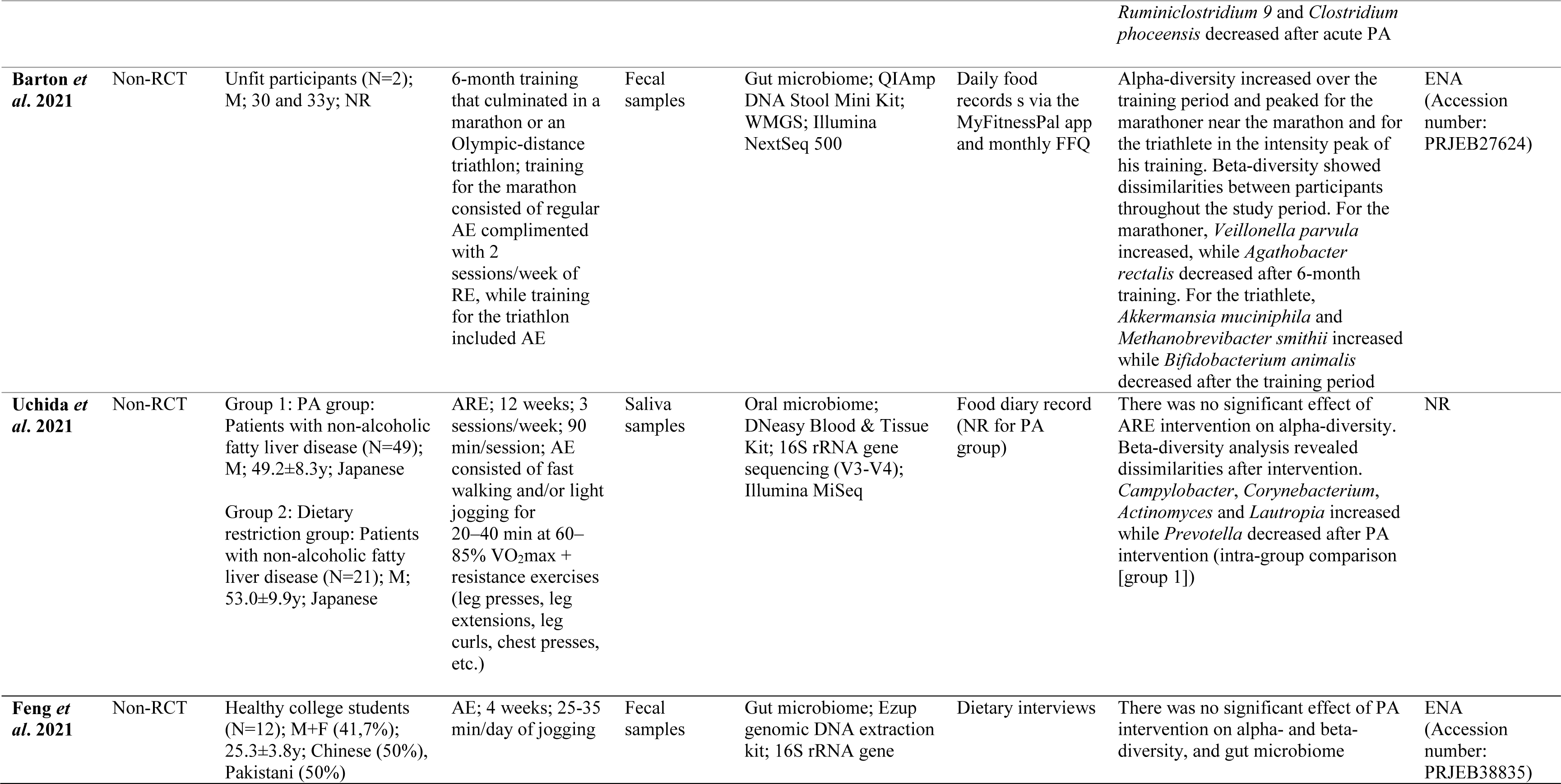
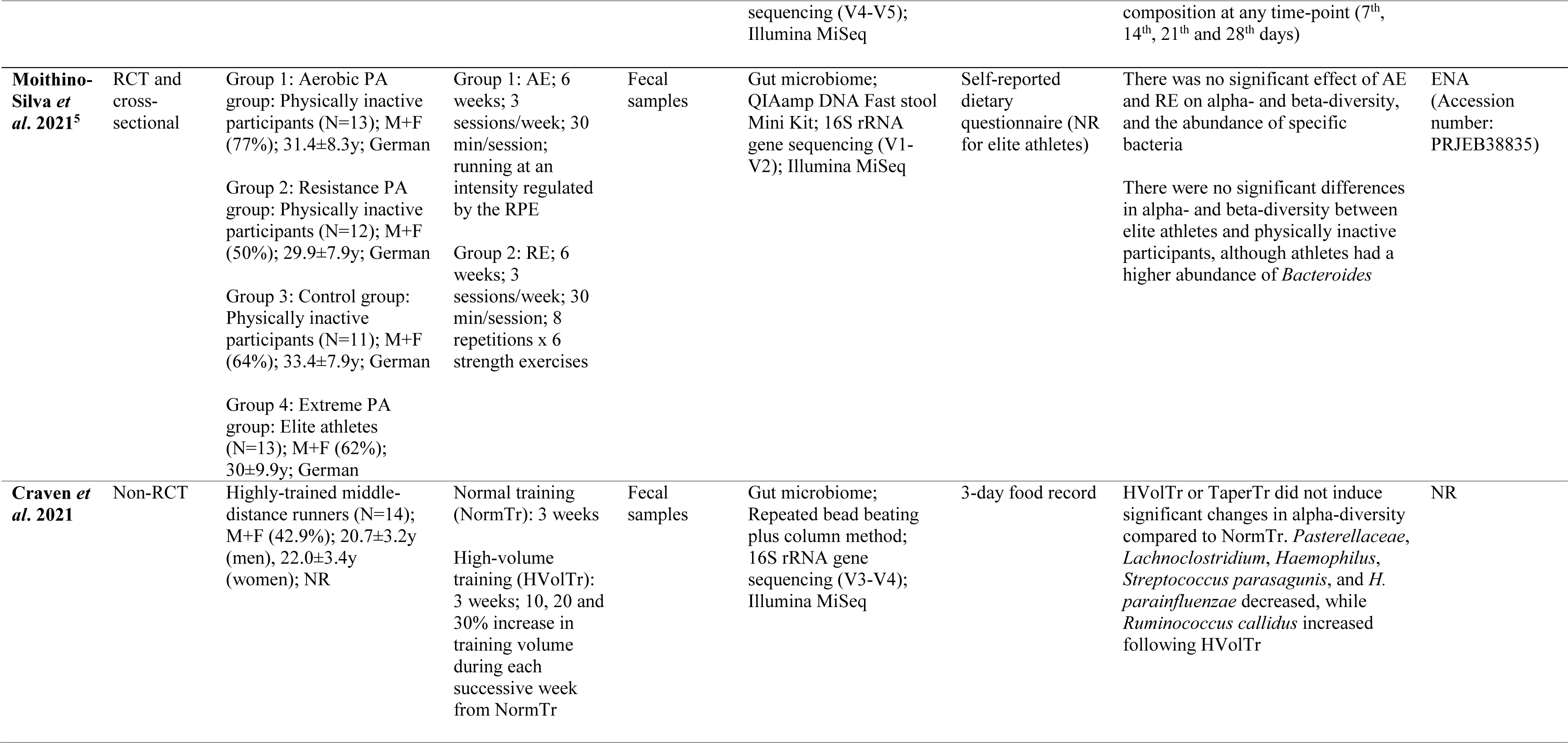
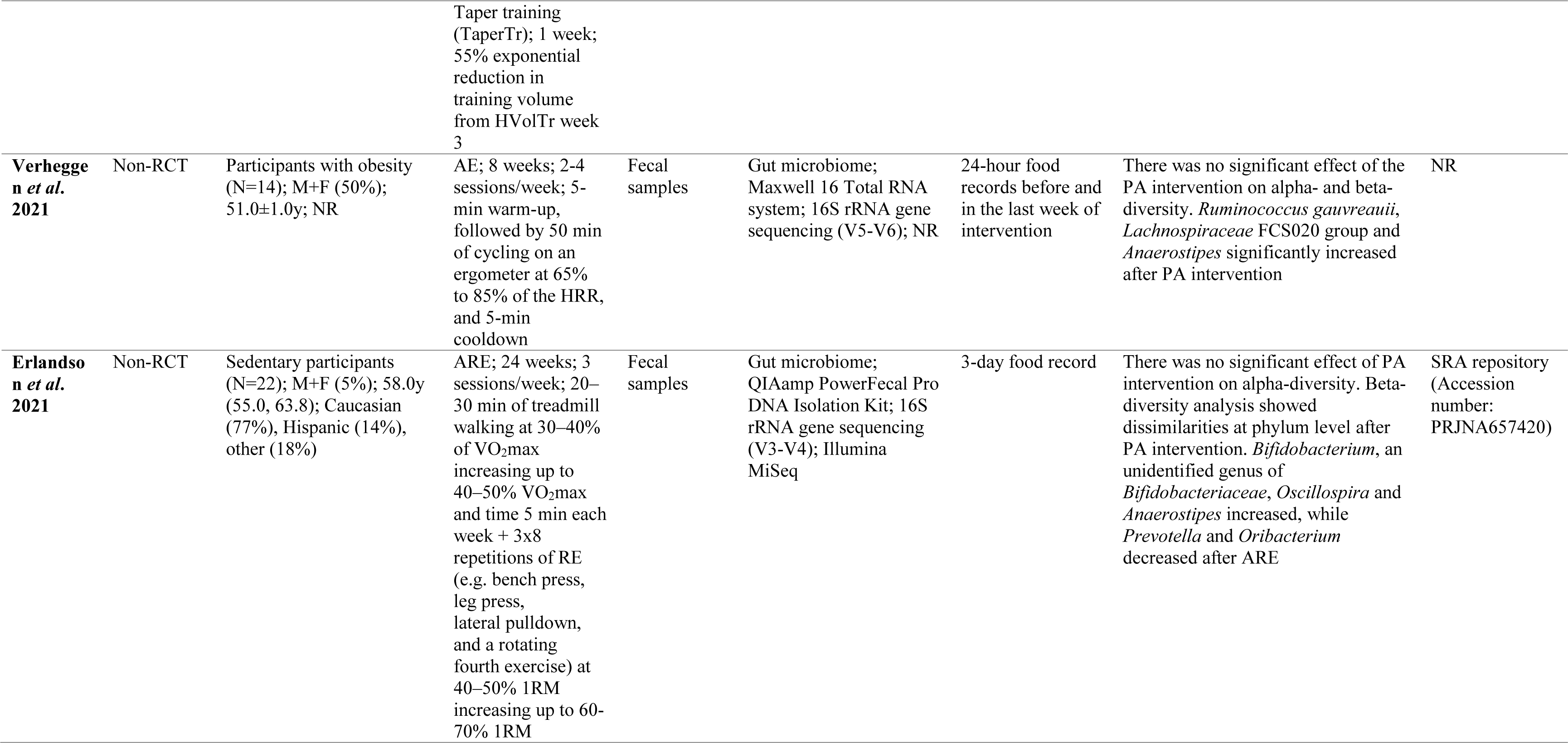
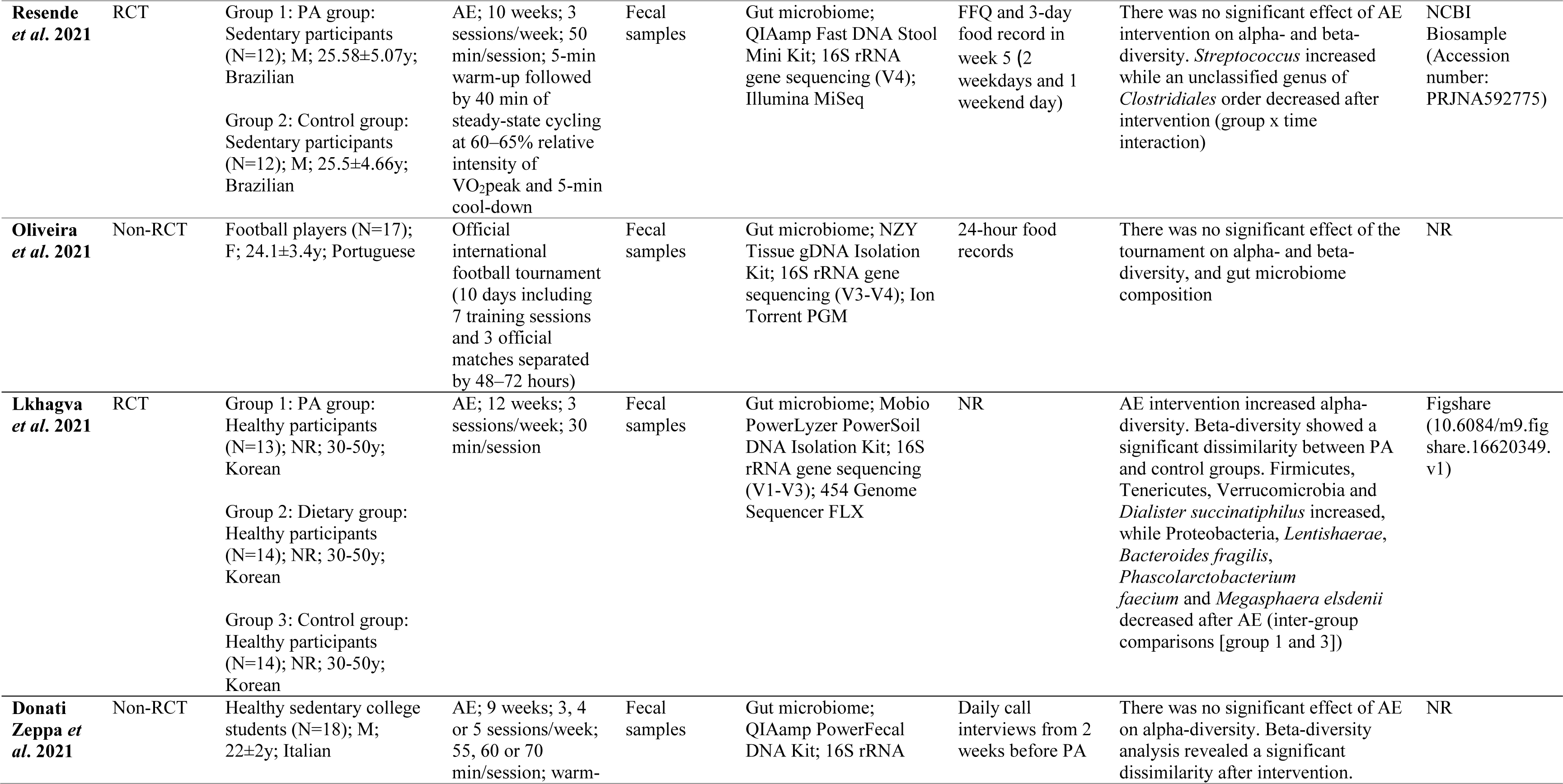
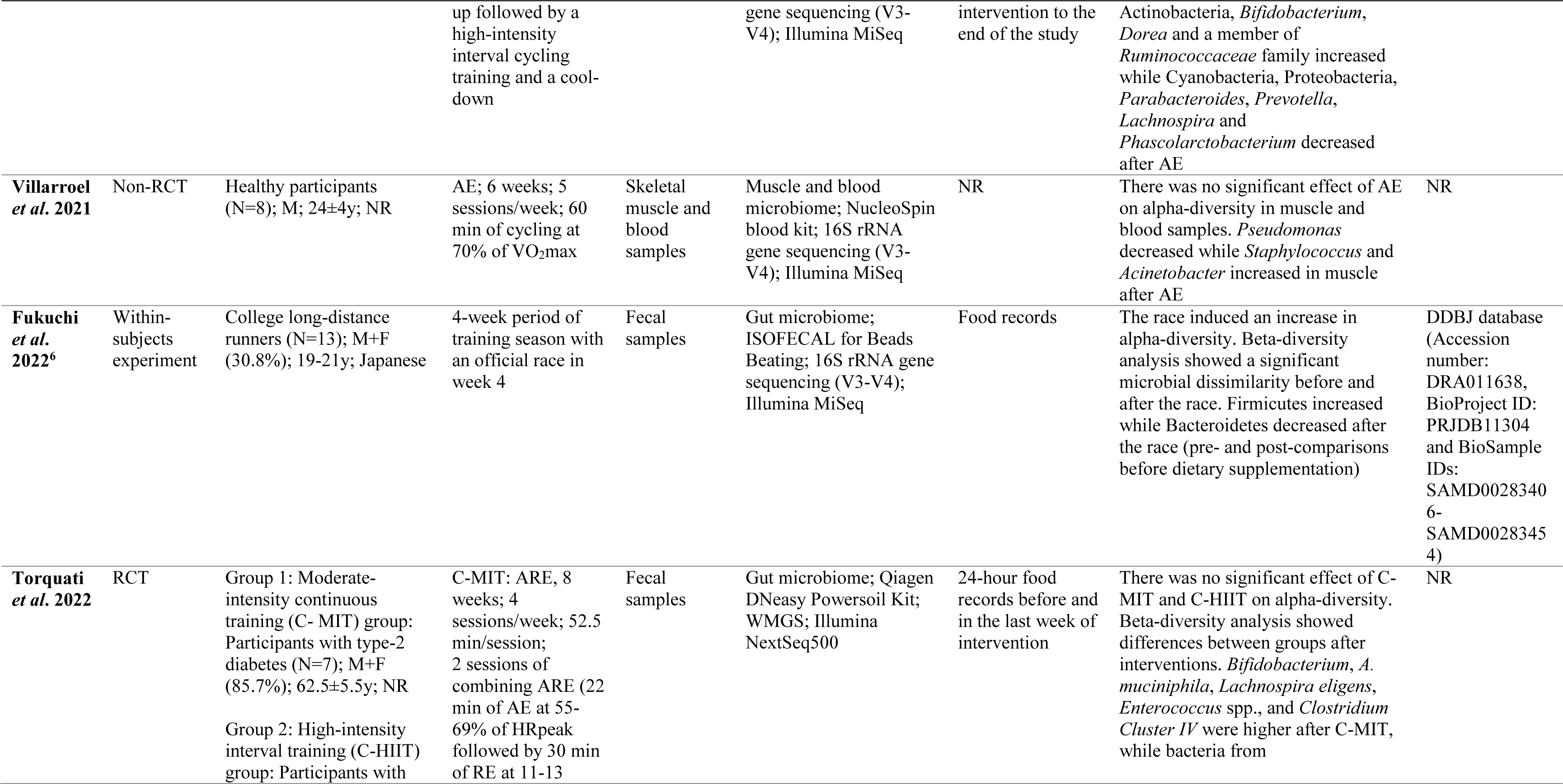
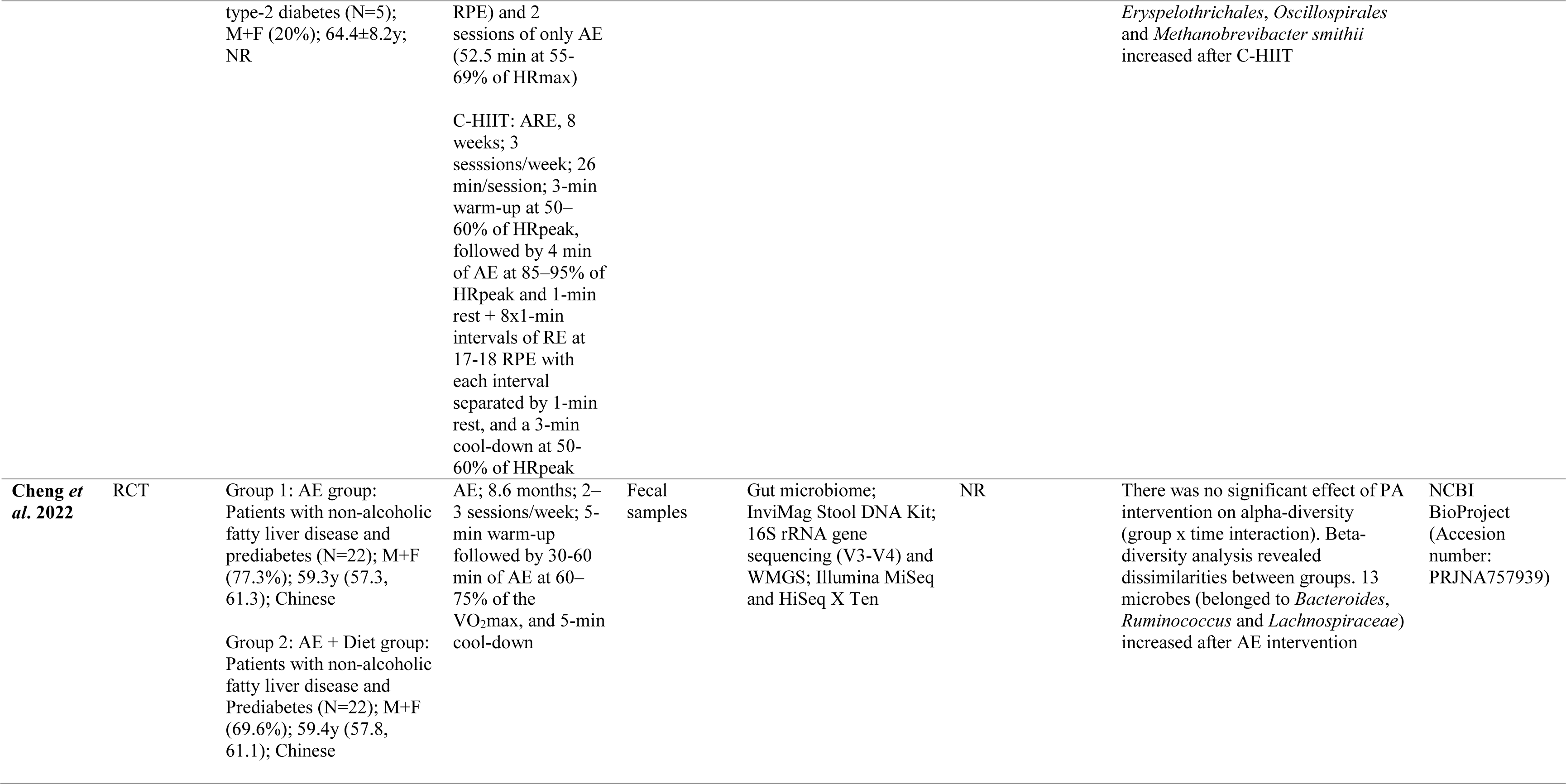
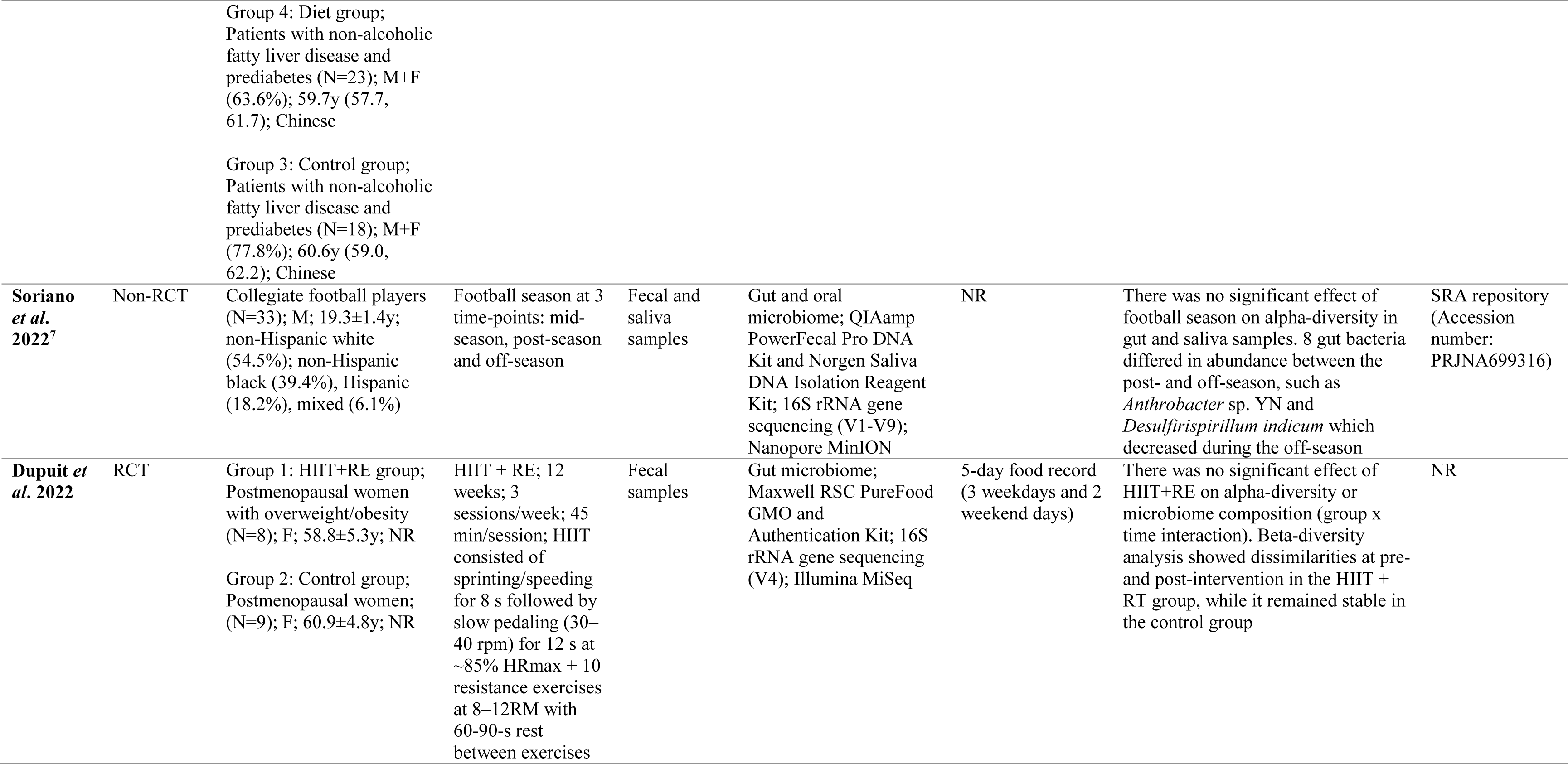
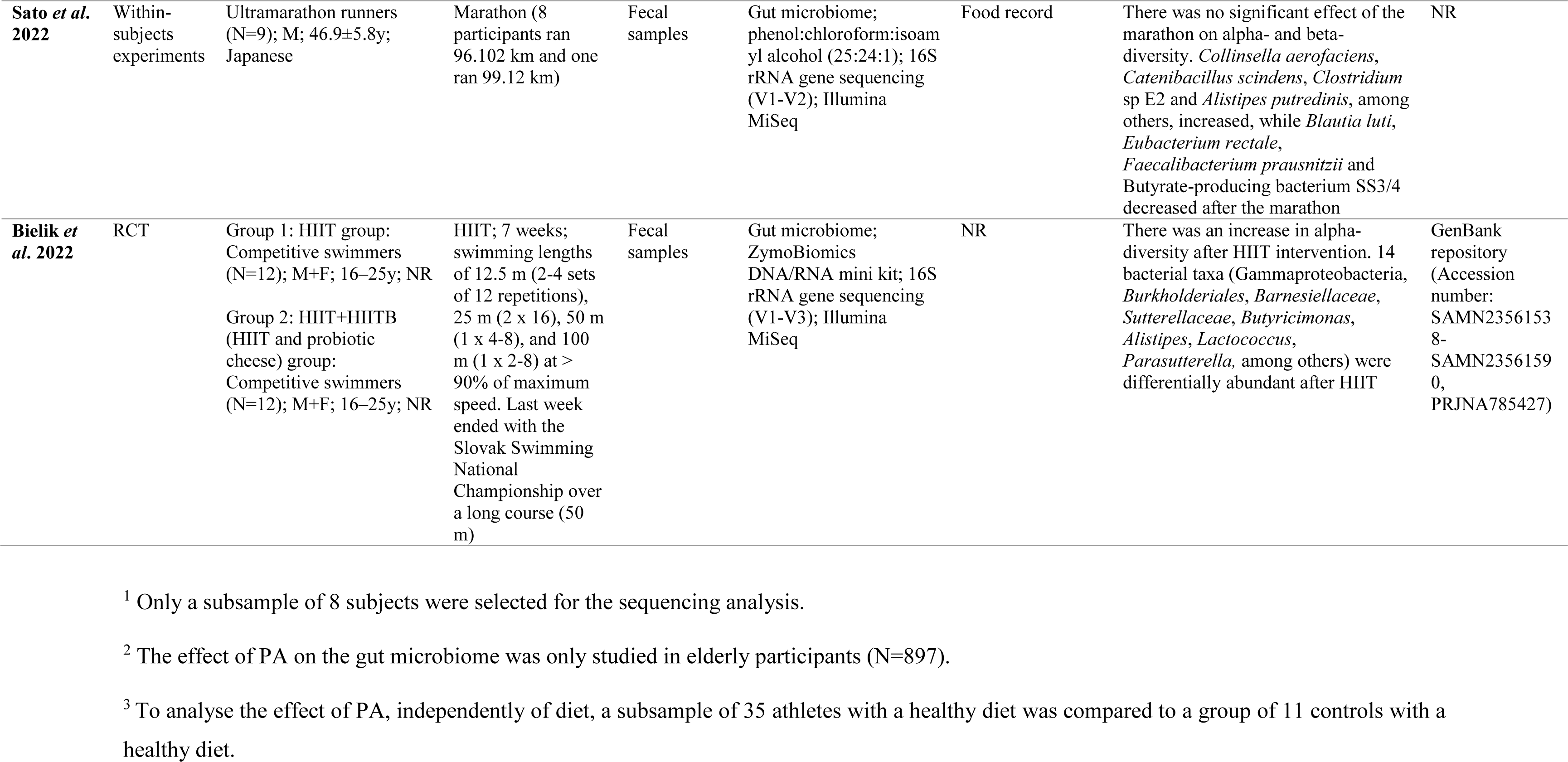
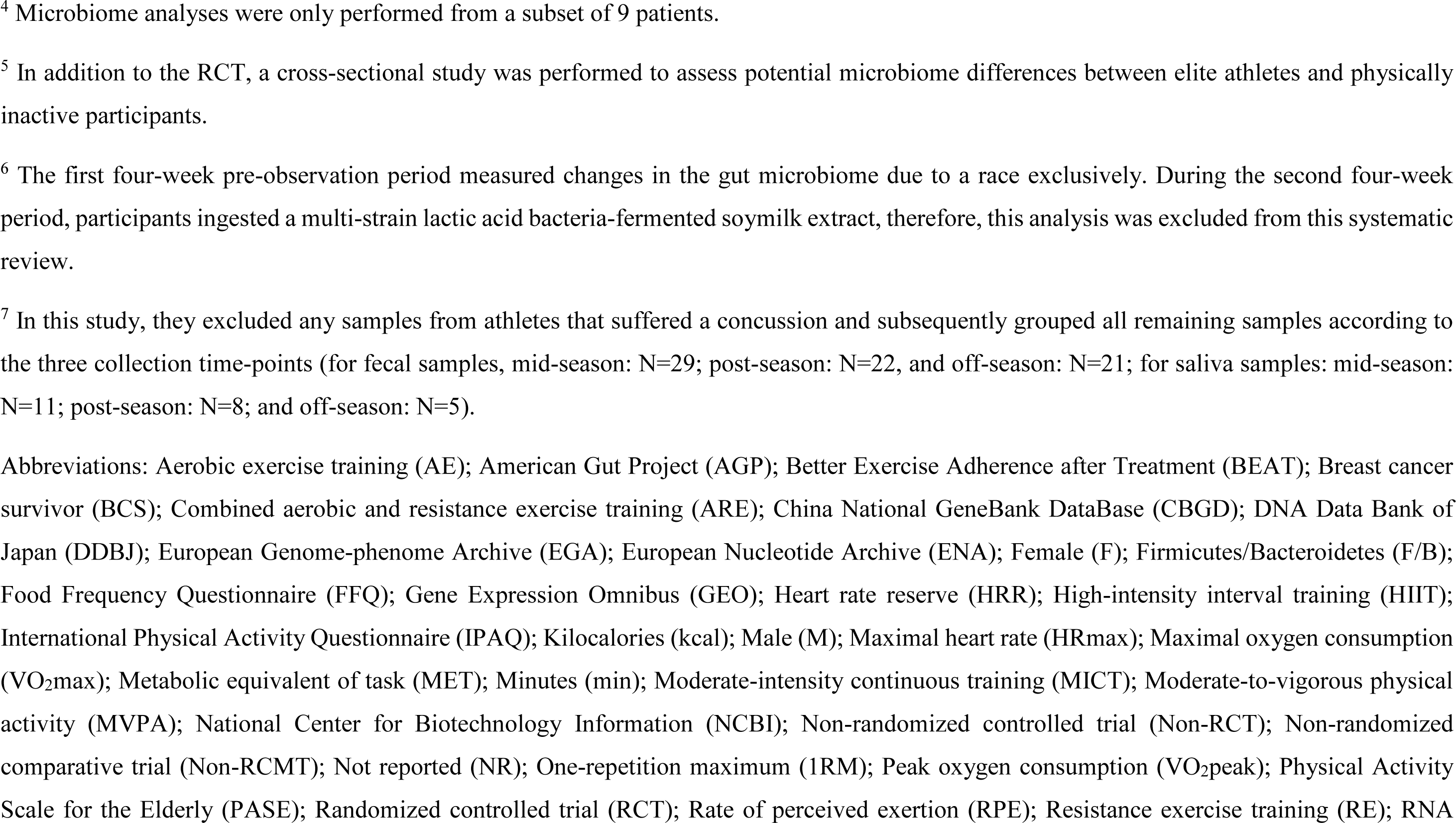
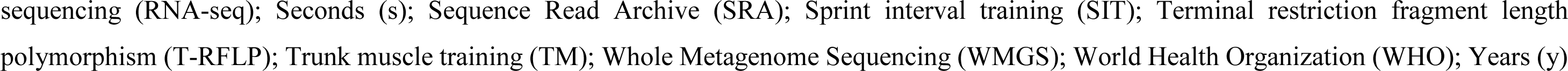
Summary of the main characteristics of the studies included in this systematic review.

The sample sizes ranged from 1 ^90^ to 2183 ^65^ (**Table 2**). Fifty-three studies involved both male and female participants ^23,36,55–57,59,63–68,38,69–73,77–81,39,83,85– 87,92,94,96,98,100,103,41,104–108,114,115,117,119,120,42,121,123,125,45,46,50,54^, while 12 were exclusively conducted on women ^37,43,110,118,48,51,58,74,82,88,95,99^ and 25 on men ^40,44,76,84,89– 91,93,97,101,102,109,47,112,113,116,122,124,49,52,53,60–62,75^. One study did not report the gender of the participants ^111^. Regarding age, 5 studies recruited children (i.e., 7-12 years) and/or adolescents (i.e., 13-17 years) ^59,64,68,73,123^, 67 included young and middle-aged adults (i.e., 18-64 years) ^23,36,49–55,60–62,40,65,66,70,71,74,76–80,42,82,85–93,43,94–96,98,100–105,44,106–113,115,116,45,117,118,120–122,124,125,46–48^, 12 older adults (i.e., ≥65 years) ^37,38,99,114,39,41,57,63,69,72,84,97^, 3 studies combined adolescents and adults ^56,58,119^ and 4 adults of different age ^67,75,81,83^. Fifty-nine studies were performed on healthy individuals ^23,36,49,50,52–59,37,60–62,64–66,68,70,71,74,40,75,76,78–81,83,85,87,89,42,90–93,96–100,103,43,104,105,109–113,116,119,44,45,47,48^, while 32 studies included participants with different diseases such as obesity or breast cancer, among others ^38,39,77,82,84,86,88,94,95,101,102,106,41,107,108,114,115,117,118,120– 123,46,124,125,51,63,67,69,72,73^.

Regarding the exposure, 26 cross-sectional studies recorded PA using self-reported questionnaires ^36,37,46,48,59,65–68,70–72,38,73,78,81,83–85,39–45^, whereas 8 studies included PA data registered by accelerometry ^42,48,63,66,69,82,84,85^ (**Table 2**). Additionally, four studies reported SB data expressed as time per sedentary breaks/bouts or screen time ^48,64,70,73^. Twenty-two cross-sectional studies recruited athletes from different sports such as rugby, athletics or football, among others ^47,49,58,60–62,74–79,50,80,104,51–57^. Six studies analyzed the effects of a marathon, footrace or rowing race on microbiome ^23,87,89,90,92,93^, three reported the effect of a single bout of PA (i.e., no sport competition) on microbiome ^86,88,91^ and 32 conducted a long-term PA intervention ranged from two weeks ^121^ to thirty-four weeks ^115^, mostly consisting of aerobic training ^94,95,113,115,119–121,97,98,103,105,107,109,111,112^ or a combination of aerobic and resistance training ^96,99,116–118,122–125,100–102,104,106,108,110,114^.

Most of the studies analyzed the gut microbiome, with the exception of ten which collected saliva, oral, oropharyngeal, muscle, blood or vaginal samples ^36,43,52,64,80,86,88,102,113,116^ (**Table 2**). Concerning the detection method, 78 studies conducted the 16S rRNA gene sequencing approach to characterize the microbiome ^23,36,45–48,51–53,55,56,58,37,59–64,66–69,38,70–79,39,80–87,90,91,40,92–95,97–100,102,103,41,104,105,107–113,115,42,116,118–121,123–125,43,44^, 16 performed metagenomics analyses ^23,49,101,106,114,115,117,122,50,54,57,60,65,89,95,96^ and two studies focused on meta-transcriptomics (i.e., microbial RNA-sequencing) ^50,88^. Twenty-one studies did not report dietary data for all the participants ^36,39,102,104,106,111,113,115–117,119,121,74,123,76,79,80,86,88,98,99^. One study performed a control of diet (each participants received the same kind of food) during the PA intervention ^87^.

Figure 2 shows a graphical summary of the main findings. Specific outcomes of microbial composition identified in the articles are discussed and interpreted in the context of the current knowledge in the Discussion section. For further details, see **Electronic Supplementary Appendix S2**.

**Fig. 2.**
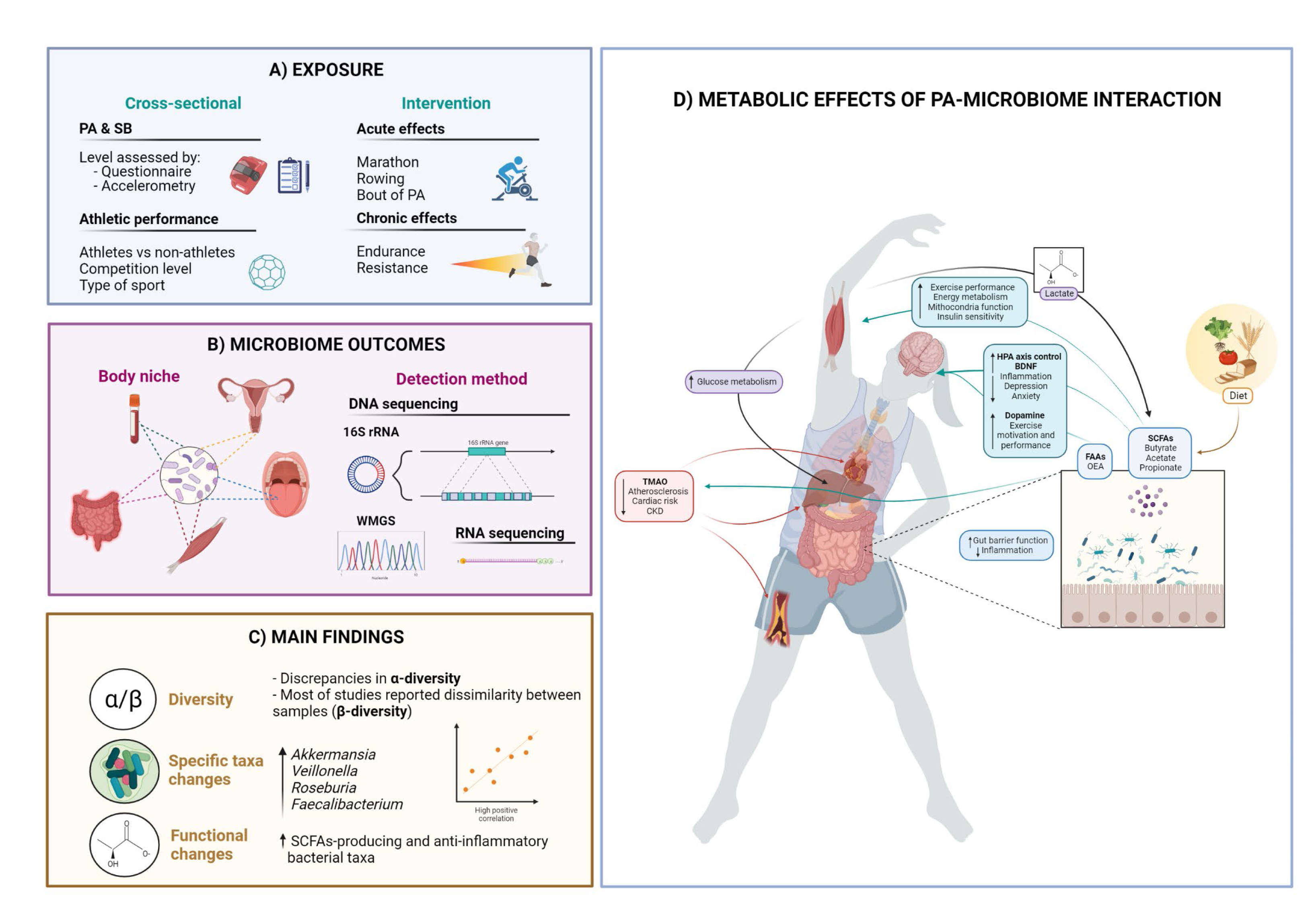
Summary of the main characteristics and findings of the studies included in this systematic review. A) Exposure: in cross-sectional studies, the effect of physical activity (PA), sedentary behavior (SB) or athletic performance on microbiome was analyzed. In intervention studies, the acute or chronic effects of PA on microbiome were evaluated. B) Microbiome outcomes: samples from different body sites (gut, saliva, blood, muscle and vagina, among others) were analyzed by distinct detection methods (16S rRNA sequencing and whole metagenome sequencing [WMGS] for DNA-based microbiome analysis; meta-transcriptomic [RNA sequencing] for RNA-based microbiome analysis). C: Main findings: relevant results concerning alpha- and beta-diversity and differential abundance analysis are shown. D) Metabolic effects of PA-microbiome interaction. Growing evidence indicates that PA increases the abundance of members of the Firmicutes phylum, bacteria able to produce short-chain fatty acids (SCFAs). SCFAs produced by the gut microbiome by processing nutrients from diet may have positive effects in the intestine, improving barrier function and inflammation state. A crosstalk between the gut microbiome and skeletal muscle through lactate (generated during PA) and its conversion to SCFAs may improve athletic performance. SCFAs have been also linked to promoting neurogenesis (through brain-derived neurotrophic factor [BDNF]), improving hypothalamic–pituitary–adrenal (HPA) axis control, reducing inflammation and the risk of psychological diseases (e.g. depression, anxiety). A microbiome-dependent gut-brain connection mediated by microbial metabolites (i.e., fatty acid amides [FAAs], such as N-oleoylethanolamide [OEA]) has been discovered in mice, which enhances exercise performing and motivation by increasing dopamine signaling during PA. Recent studies suggest that dysbiosis may lead to the growth of proteolytic microbes able to produce trimethylamine-N-oxide (TMAO), an important metabolite that in elevated concentration has been linked to adverse cardiac events and chronic kidney diseases (CKD). This figure was created with BioRender.com

### 3.2 Quality assessment

Among the 50 cross-sectional studies, 26 were categorized as high quality (quality score ≥ 75%), whereas 24 as low quality (quality score < 75) (**Electronic Supplementary Table S4**). Regarding the 9 studies about the acute effects of PA, 8 studies were considered to have a high quality and 1 showed a low quality (**Electronic Supplementary Table S5**). Concerning the 32 studies (15 RCTs and 17 non-RCTs) that reported the chronic effects of PA interventions, one RCT presented a high quality and 14 a low quality (**Electronic Supplementary Table S6**), while 12 non-RCTs were categorized as high quality and 5 as low quality studies (**Electronic Supplementary Table S7**).

### 3.3 Meta-analysis

#### 3.3.1 First meta-analysis (cross-sectional studies): high *vs*. low PA levels

This meta-analysis united 1814 participants from 8 studies, where 1157 belonged to the high PA and 657 participants to the low PA groups. No significant differences were reported between the groups of high and low PA levels on alpha-diversity represented by the Shannon diversity index (SMD= -0.101, 95% CI -0.386-0.184, p=0.488, I^2^= 33.581) and Chao1 index (SMD= -0.127, 95% CI -0.563-0.309, p=0.568, I^2^= 13.774) (Fig. 3A).

**Fig. 3.**
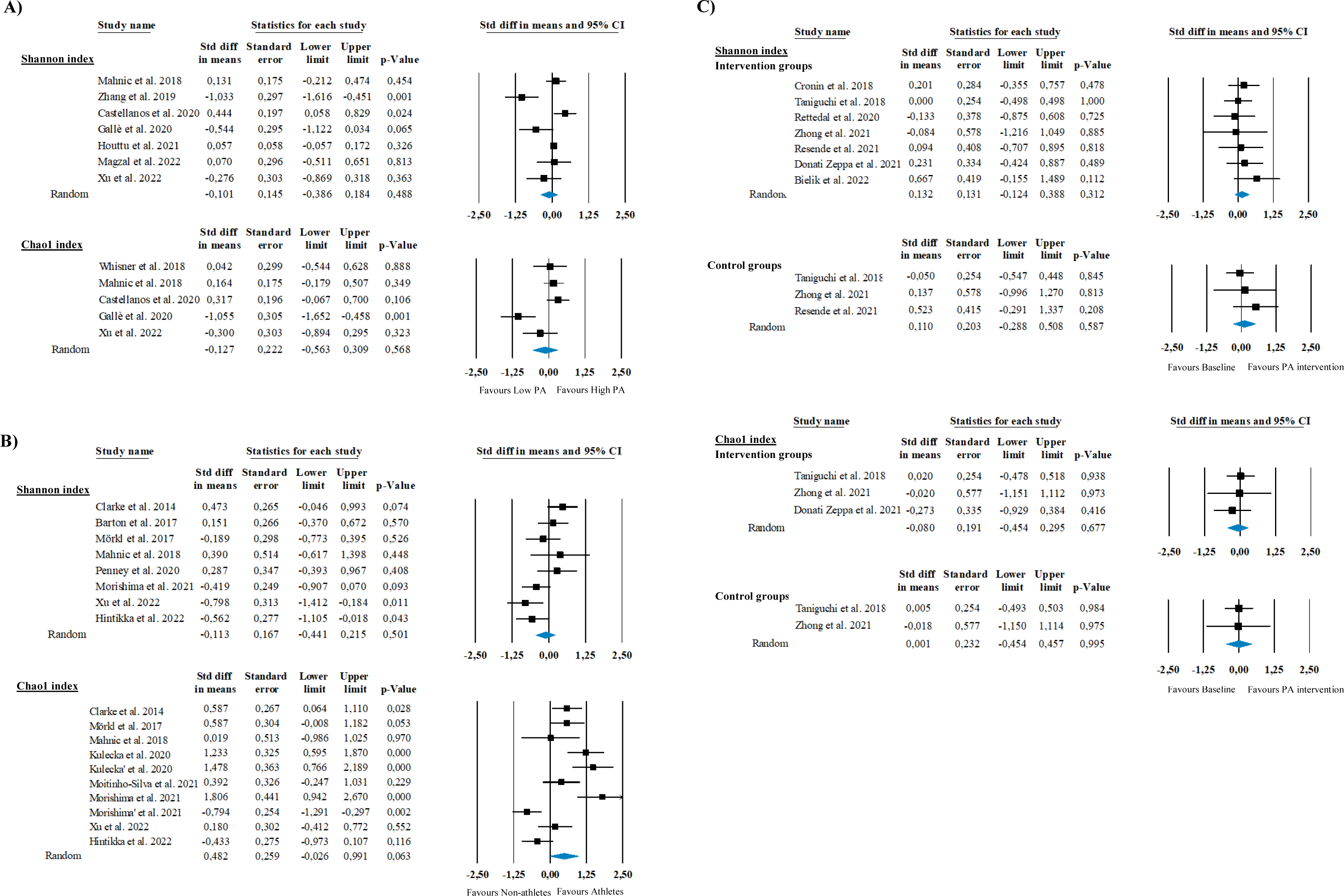
Panel A) shows the meta-analysis of the high-PA *vs*. low-PA level’s effects on Shannon diversity and Chao1 indexes (i.e., alpha-diversity metrics). Eight studies were finally included (Shannon diversity index ^44,66,69,78,81,83,85^; Chao1 index ^44,70,78,81,85^). Panel B) indicates the meta-analysis of the athletes *vs*. non-athletes’ effects on both alpha-diversity metrics, i.e., Shannon diversity (8 studies ^47,49,51,60,76,78,79,81^) and Chao1 indexes (9 studies ^47,51,56,74,76,78,79,81,104^). Panel C) shows the meta-analysis of the PA intervention (up) *vs*. control’s effects (down) on the Shannon diversity (7 studies ^96,97,99,109,112,119,124^) and Chao1 index (3 studies ^97,99,112^). Due to the lack of studies, we included both RCTs ^96,97,99,109,119^ and non-RCTs ^112,124^ in the same meta-analysis. The bottom meta-analyses reflect the effect of time in the absence of PA intervention since only includes the control groups that were available from the RCTs. We did not use the control groups of Cronin *et al*. 2018 and Bielik *et al*. 2022, as they consumed a protein or probiotic supplement^96,119^

#### 3.3.2 Second meta-analysis (cross-sectional studies): athletes *vs*. non-athletes

This meta-analysis comprised 651 participants from 11 studies, including 329 athletes and 322 non-athletes. No significant differences were reported between the groups of athletes and non-athletes on alpha-diversity using the Shannon diversity index (SMD= -0.113, 95% CI -0.441-0.215, p= 0.501, I^2^= 0.000). However, athletes tended to present a higher alpha-diversity compared to non-athletes’ when Chao1 index was used as an indicator of microbial alpha-diversity (SMD= 0.482, 95% CI -0.026-0.991, p=0.063, I^2^= 0.000) (Fig. 3B).

#### 3.3.3 Third meta-analysis (intervention studies): chronic effects of PA

The third meta-analysis united 167 participants from 7 studies, where 118 were allocated to a PA group and 49 to a control group. No significant differences were found between the PA and control groups on alpha-diversity using the Shannon diversity index (PA group: SMD=0.132, 95% CI -0.124-0.388, p= 0.312, I^2= 0.000; control group: SMD= 0.110, 95% CI -0.288-0.508, p= 0.587; I^2^= 0.000) or Chao1 index (PA group: SMD= -0.080, 95% CI -0.454-0.295, p= 0.677, I^2^= 0.000; control group: SMD= 0.001, 95% CI -0.454-0.457, p= 0.995; I^2^= 0.000) (Fig. 3C).

## 4. Discussion

The main findings of this systematic review and meta-analysis were: 1) there was no consistent effect of PA on modifying microbial alpha-diversity, although most of studies support that PA (observational and intervention studies) induces changes in microbiome composition with the increase of short-chain fatty acids (SCFAs)-producing bacteria such as *Akkermansia*, *Roseburia* or *Veillonella*, among others; 2) there is very limited evidence of the effect of SB on microbiome; 3) few studies assessed PA data by objective methods (i.e., accelerometry); 4) there are few studies about the acute effect of PA on microbiome; 5) available studies are hardly comparable due to heterogeneity of the participants (i.e., age, sex, health status), wide use of different self-reported questionnaires to record PA, lack of standardized criteria to stratify participants in active/sedentary groups in cross-sectional studies and different characteristics of PA interventions (e.g., type, intensity, duration); 6) most of studies did not include diet as a confounder in their statistical analyses; 7) well-designed multi-omics studies (i.e., metagenomics, meta-transcriptomics, meta-proteomics and meta-metabolomics) are warranted to clarify the effect of PA and SB on microbiome.

### 4.1 Cross-sectional studies: physical activity and sedentary behavior (non-athletes)

Microbiome diversity is considered a direct measure of gut health, and a loss of diversity has been linked to a higher risk of obesity, type 2 diabetes, and cancer, among others ^126^. In this systematic review, four studies found that the gut microbiome of children and adults with higher PA levels showed higher alpha-diversity, compared to those who rarely or never exercised ^59,65,66,85^. Similarly, two studies reported a positive association between PA level and gut alpha-diversity in participants with different diseases ^38,67^. A positive correlation between average PA intensity and vaginal microbiome alpha-diversity was also found in healthy college-aged women ^43^. However, other studies in individuals with different age and health conditions reported negative or no associations ^36,39,78,81–84,41,44,48,63,68–71^, as is also detected in our meta-analysis of 1814 participants (Fig. 3A). Heterogeneity in study population (i.e., health status, sex, age), methodological aspects (i.e., use of diverse self-reported questionnaires, different pipelines to analyze the microbiome, etc.), varying criteria to stratify participants based on PA level and lack of control of relevant covariates (e.g. diet) in statistical analyses may contribute to the discrepant findings across studies. In fact, Langsetmo *et al*. demonstrated different results depending on the method for measuring PA, where self-reported PA was positively associated with beta-diversity ^84^, while objectively measured PA recorded by accelerometry (expressed as step counts) showed no associations ^84^.

Regarding SB, Bressa *et al*. reported that less time in sedentary bouts was positively associated with alpha-diversity (Shannon and Chao1 indexes) in premenopausal women ^48^. In contrast, there were no significant differences in alpha-diversity when compared the gut microbiome of physically active women (those who perform at least 3 hours of PA per week) and sedentary women (i.e., those who perform <3 hours) ^48^. Whisner *et al*. did not find any significant differences in alpha-diversity parameters across quartiles of SB in a cohort of college students ^70^. However, a later study detected an increased alpha-diversity in the saliva of children who reported less screen time ^64^. Interestingly, recent evidence indicates a positive association between SB and *Streptococcus,* detected in feces and saliva ^63,64^. *Streptococcus* has been described as a key bacteria in disease such as old-onset colorectal cancer ^127^. The existence of an oral-gut microbiome crosstalk has been proposed, highlighting a possible association between oral dysbiosis, oral–gut microbiome axis and the pathogenesis of different diseases such as gastrointestinal disease or colorectal cancer ^128^. Thus, more future research is needed to unravel the role of SB as a potential modulator of microbial communities.

There are more consistent findings about the associations between PA and the gut microbiome, mostly at lower taxonomic categories. At phylum level, Firmicutes seems to be more abundant in the gut of those individuals with higher PA levels ^41,59^, although several studies found the inverse association ^44–46^. Since Firmicutes has been associated with fiber ^129^, different dietary habits may be partially explaining variability between the studies. Interestingly, growing evidence supports that PA increases the abundance of a Firmicutes-belonging group of commensal bacteria able to produce SCFAs from non-digestible carbohydrates ingested through diet, such as butyrate, propionate and acetate^130^. Most of the included studies reported higher abundances of SCFAs-producing bacteria from *Lachnospiraceae* and *Erysipelotrichaceae* families ^42,59,63,66,70^, and *Roseburia*, *Coprococcus*, *Lachnospira*, *Blautia* and *Faecalibacterium* genera, among others, in more active individuals compared to those with lower PA levels ^42,59,70,71,84,85^. Particularly, Bressa *et al*. quantified the relative abundance of *Akkermansia muciniphila*, *Faecalibacterium prausnitzii* and *Roseburia hominis* by real-time PCR (qPCR) and detected higher abundances in physically active compared to inactive women ^48^. SCFAs have been linked to good health due to their role on metabolic function, being substrates for energy metabolism as well as important signaling molecules implicated in the gut-microbiota axis and in the regulation of the immune response ^131,132^. Since the availability of SCFAs are influenced by both, the ingestion of nutritional components and their digestion directed by the gut microbes ^130^, the previous results could indicate SCFAs as the key molecular link between PA, diet and microbiome.

### 4.2 Cross-sectional studies: athletes *vs*. non-athletes

Available evidence generally agrees on a trend towards the increase of the gut microbial diversity in athletes of different sports disciplines compared to non-athletes (see our meta-analysis of 651 participants; Fig. 3B). Further, a recent meta-analysis evaluated microbial alpha-diversity of shotgun metagenomics data of the gut microbiomes of 207 athletes of different sports and 107 non-athletes and found a significantly higher species richness in athletes compared to non-athletes ^133^. However, it is also well known that specific dietary requirements are usually implemented based on the duration and intensity of PA training ^134^, which makes difficult to determine the isolated effect of athletic performance on the microbial communities. In 2014, Clarke *et al*. reported, for the first time, a positive association between athletic performance and alpha-diversity parameters, when compared the gut microbiome by 16S rRNA sequencing of a group of professional rugby players and sedentary participants with low and high BMI (i.e., BMI≤25 or >28, respectively) ^47^. However, the athletes’ enhanced diversity was also associated with high protein consumption in this group. Barton *et al*. ^49^ re-analyzed the participants from Clarke *et al*. to evaluate the microbiome diversity with the whole metagenome shotgun sequencing, confirming the previous results ^47^. More recently, Penney *et al*. analyzed the combined effects of diet and athletic performance in the gut microbiome of those participants, and found a significant association with alpha-diversity when combined the effect of both athletic performance and dietary habits ^60^. Later studies described an enriched microbial diversity in athletes with special diets, compared to sedentary participants ^61,62^, and others did not find any significant differences between athletes and non-athletes with similar dietary patterns ^52,53,57^. In contrast, 2 studies reported a higher alpha-diversity in athletes compared to sedentary participants with similar dietary habits ^51,56^. Large variety of sports disciplines included in the abovementioned studies (marathon runners, bodybuilders, cross-country skiers, rugby players, etc.) can be also contributing to inconsistency of the results. So far, the isolated effect of athletic performance, independently of diet, is still unclear.

Since diet is one of the most important modulators of the microbiome, differences in nutritional habits may also affect the relative abundance of specific microorganisms^135^. In fact, high-digestible carbohydrate diets have been related to the growth of SCFAs-producing bacteria. Clarke *et al*. reported a higher abundance of Firmicutes phylum and a decreased abundance of Bacteroidetes in rugby players compared to sedentary individuals with high BMI ^47^. Both groups presented a distinct nutritional profile, with an increased consumption of protein, fiber, carbohydrate and monounsaturated and polyunsaturated fat in the athletes group. A later study also described a higher abundance of Firmicutes and lower levels of Bacteroidetes in rugby players compared to non-athletes^76^. Accordingly to these findings, animal and human studies have positively associated Firmicutes to fiber intake but negatively to fat consumption, while Bacteroidetes showed the opposite association ^129^. Additionally, later metabolic pathway analyses revealed that rugby players had an enriched profile of SCFAs ^49^. Other SCFAs-producer, *F. prausnatzii,* was also found to be more abundant in senior athletes compared to older sedentary participants after adjusting for different covariates, including diet ^57^. Morishima *et al*. found an increase of *Faecalibacterium* in female runners compared to non-athletes, and a higher abundance of succinate, a SCFA that can be produced by *Faecalibacterium*^74^.

Liang *et al*. reported that professional martial arts athletes had an enriched microbiome compared to amateurs, and identified changes in the abundance of several bacteria after adjusting for different confounders including diet ^55^. Furthermore, one study found higher diversity and Firmicutes/Bacteroidetes ratio in female elite compared to non-elite athletes ^58^. However, metagenomics and meta-transcriptomics analyses conducted by Petersen *et al*. only detected differences at transcriptomic (RNA) level, highlighting the need for more microbiome studies at functional level ^50^.

Few studies have identified significant microbial shifts in relation with the type of sport ^53,54,56^. Interestingly, O’Donovan *et al*. compared athletes from 16 different sports and found specific bacterial taxa such as *Anaerostipes hadrus*, *F. prausnitzii* and *Bacteroides caccae*, differently abundant between sports with a moderate-dynamic component (e.g. fencing), high-dynamic and low-static components (e.g. field hockey), and high-dynamic and static components (e.g. rowing) ^54^.

### 4.3 Acute effects of PA

Most of the studies aimed to analyze potential changes in the gut microbial composition following a marathon ^23,87,90^. In this sense, two studies detected an increase in Firmicutes/Bacteroidetes ratio of the gut microbiome in long-distance runners post-race ^90,92^. Significantly, Grosicki *et al*. also detected a higher abundance of *Veillonella*, accordingly to the results obtained by Scheiman *et al*. ^23,90^. The last study proposed a microbiome-encoded enzymatic mechanism that could partially explain how microbiome and its metabolites (i.e., SCFAs) may contribute to enhance athletic performance ^23^. After detecting a higher abundance of *Veillonella* in runners after the race, they observed that administration of *Veillonella atypica* in a mouse model improved run time and demonstrated its capability of metabolically converting the exercise-induced lactate into propionate in the colon to subsequently re-enter the systemic circulation. In search of confirming these findings, Moitinho-Silva *et al*. quantified the relative abundance of *V. atypica* by qPCR and sequencing in a subset of elite athletes (mainly cyclists and triathletes) and sedentary participants, but failed to find any significant differences between the groups ^104^. These contrasting results could be partially explained by several limitations of the last study such as the lack of dietary data for the athletes group. Other studies have detected an increase in several SCFA-producing bacteria, including *Coprococcus*_2, *Dorea* or *Roseburia* after a marathon or a transoceanic rowing race ^87,89^. Although the number of human studies is still limited, these findings support emerging evidence of the existence of a crosstalk between the gut microbiota and skeletal muscle through lactate (generated during exercise) and its conversion to SCFAs by the gut microbes which, consequently, could improve athletic performance ^25^. In fact, SCFAs have been recently defined as “biotics” (substances able to modulate the microbiome by increasing the abundance of beneficial microbes) that could be used as an exogenous microbiome modulation approach for improving health and athletic performance ^136^. Interestingly, a recent study discovered a gut–brain connection in mice that enhances athletic performance by increasing dopamine signaling during PA ^137^. These results indicate that motivation for PA is influenced by the gut microbes derived-metabolites, suggesting a microbiome-dependent mechanism for explaining inter-individual variability in PA motivation and performance.

On the other hand, the acute effect of a bout of PA on the microbiome continues to be a scarcely investigated topic. Tabone *et al*. followed this approach analyzing fecal samples from athletes who underwent a moderate-intensity treadmill session until volitional exhaustion and detected changes in six bacteria (*Romboutsia*, *Escherichia coli* TOP498, *Ruminococcaceae* UCG-005, *Blautia*, *Ruminiclostridium* 9 and *Clostridium phoceensis*) ^91^. Overall, acute interventions collect serum samples where potential changes can be detected earlier compared to fecal ones. In this context, one study collected blood and fecal samples of myalgic encephalomyelitis / chronic fatigue syndrome participants and detected changes at major bacterial phyla such as Actinobacteria, Bacteroidetes, Firmicutes and Proteobacteria, in both samples after a cycle ergometer maximal exercise test ^86^. A later study exclusively analyzed viral reads (i.e., virome) from blood samples by RNA-seq and did not detect any differences after acute PA ^88^. Thus, more research directed to analyze blood microbiome is needed to accurately assess the short-term effect of PA, specially, meta-transcriptomics and meta-metabolomics could be a novel and useful approach to study the active microbiome in the context of acute effects of PA.

### 4.4 Chronic effects of PA

To further deepen the overall knowledge of the chronic effects of PA on human microbiome and, generally, the host health, several clinical trials have been published in the last years ^94,95,104–113,96,114–123,97,124,125,98–103^. Our meta-analysis of 167 participants is the first analysis that quantifies those trials in healthy participants, indicating that controversial results for alpha-diversity are consistently found (Fig. 3C). Two studies performed in healthy adults that underwent a 12-week aerobic PA intervention (3 sessions of 30 min per week) or 7-week high-intensity interval training (consisting of swimming lengths) reported an increase in microbial alpha-diversity ^111,119^. Conversely, Moitinho-Silva *et al*. detected a slight decrease in alpha-diversity after an aerobic PA intervention (6 weeks; 3 sessions of 30 min per week) in healthy adults [63], although no differences were observed in another group subjected to a strength training [63]. Another study recruited healthy adults to undergo a PA intervention (aerobic and resistance training; 8 weeks; 3 sessions of 90 min per week), but no significant changes in alpha-diversity were detected after the intervention ^96^. Most of the studies in unhealthy individuals did not report any significant changes in alpha-diversity after a PA intervention ^94,95,122,124,125,102,106–108,114,115,118,121^. However, the chronic effect of PA on microbiome composition becomes clearer in the beta-diversity analysis, where more studies agree on a significant dissimilarity in the microbial communities of the individuals after long-term PA ^94,95,118–120,123,125,98,101,102,108,111,112,114,115^. Interestingly, Allen *et al*. observed how differences in the beta-diversity detected at baseline between the participants with normal-weight and obesity disappeared after an aerobic PA intervention (6 weeks; 3 of 30-60 min sessions per week) ^94^. Different study designs (17 non-RCT, 13 RCT and 2 randomized controlled cross-over trials), health status of participants (15 studies with healthy and 17 with unhealthy populations), characteristics of PA interventions (type, duration, and intensity), and methodological differences in microbiome analysis, diet, among other factors, might partially influence the varying results obtained.

In accordance with observational studies ^38,48,59,63,66,69,71,74,85^, an increase in SCFAs-producing bacteria such as *Lachnospiraceae*, *Verrucomicrobiaceae, Lachnospira*, *Akkermansia*, *Veillonella*, *Faecalibacterium, Bifidobacterium* and *Roseburia* was also reported in participants with different age and health conditions (including obesity, prediabetes and insulin resistance, among others) after PA interventions ranging from 2 to 34 weeks ^94,95,122–124,101,102,107,108,112,114,115,121^. More specifically, Liu *et al*. described an increase in *A. muciniphila* and an improvement in insulin sensitivity after a 12 weeks-concurrent PA intervention in men with prediabetes that were classified as responders compared to non-responders ^122^. Later studies have also reported an increase in *A. muciniphila* in participants with overweight/obesity or type 2 diabetes after long-term PA ^101,114^. *A. muciniphila* has been related to prevention of multiple metabolic diseases such as obesity, metabolic syndrome and type 2 diabetes ^138^. In a recent publication, a multi-omics approach (transcriptomics, proteomics, metabolomics and lipidomics) investigated the underlying molecular mechanism of *A. muciniphila* in obesity. It concluded that *A. muciniphila* reduced lipid accumulation and downregulated the expression of genes related to adipogenesis and lipogenesis in adypocites ^139^. These studies point to *A. muciniphila* as a promising microbial target (potentially modulated by PA) with therapeutics effects in obesity and other metabolic diseases. At genus level, *Akkermansia* has also been widely found to be positively associated with PA in cross-sectional studies ^47,48^ and increased after PA interventions^95,122^.

### 4.5 Future Directions

Our understanding about the effect of PA (little research is conducted on SB) on microbial communities is still in its infancy, vastly limited to the amplicon sequencing approach (i.e., 16S rRNA sequencing) and is highly variable between the studies. This heterogeneity highlights the need to perform well-designed studies focusing on specific detailed populations and establishing reference pipelines to ensure the accuracy and comparability of the results. To ensure the reproducibility and comparability of the future studies in the field, we recommend the researchers to follow the recent good practice guidelines ^140,141^ when microbiome analyses are performed.

Most of cross-sectional studies in this systematic review recorded PA measures by self-reported questionnaires. Accelerometry has been widely demonstrated to be a more valid and comparable method for objectively collecting participants’ PA and SB levels ^142^. Therefore, more accelerometry-based studies will allow researchers to apply standardized criteria to classify participants based on the use of cut-points for PA and SB which will reduce the inconsistency between study findings and reveal the accurate association of PA and SB with microbiome. In intervention studies that assess the chronic effects of PA on microbiome, we detected a low quality in the RCTs. These results could be partially explained by the use of a checklist ^143^ with a stricter scale for the quality assessment.

Since most of the studies analyzed DNA sequences regardless of microbial variability or functionality (only two studies performed a meta-transcriptomic analysis), we are not close to determine the functional microbes susceptible to PA. Moreover, future multi-omics analyses (i.e., combining metagenomics, meta-transcriptomics, meta-proteomics and meta-metabolomics) would further unravel the complex host-microbial molecular pathways implicated in the molecular response to PA. In this regard, the Molecular Transducers of Physical Activity Consortium (MoTrPAC) ^6^ will provide a powerful source of information to advance our understanding of PA’s effects on the microbiome in humans and animal models performing multi-omics analyses.

### 4.6 Limitations and strengths

Due to the lack of available information, an important limitation of our meta-analysis was the use and transformation of directly reported data from the articles instead of re-analyzing raw data to reduce potential bias introduced by applying different methodologies and pipelines across studies. Besides, limited information prevented us from additionally analyzing other microbiome outcomes of interest, such as the differential abundance of key bacteria. Future studies should make publicly available raw sequences generated from sequencing platforms to allow future meta-analyses to cover these gaps in the literature. Nevertheless, our meta-analysis has been performed including specifically those studies in healthy population and conducting sub-groups analysis according to study design to gain homogeneity. Moreover, we followed a rigorous and reliable methodology previously validated ^144,145^ to obtain numerical data when they were unavailable. Additional strengths of our systematic review are the elaboration according to PRISMA guidelines, use of four different search databases (PubMed, Web of Science, SCOPUS and Cochrane), and performance of quality assessment with validated tools specific for each study design, which ensure the scientific rigor.

## 5. Perspective

Our systematic review summarizes the available knowledge about the relationship between PA and SB and the microbiome from multiple body sites and across different human populations. So far, growing evidence points to higher abundances of SCFAs-producing bacteria in more active individuals or after a PA intervention. Our meta-analysis uniting 2632 participants indicated no consistent effect of PA on microbial alpha-diversity, although there seems to be a trend toward a higher richness in athletes compared to non-athletes. Thus, accelerometry-based observational studies and RCTs are needed to face this inconsistency. Additionally, there are scarce information about the effect of SB on microbiome. In conclusion, precisely-designed, well-controlled and multi-omics studies are needed to reduce heterogeneity, obtain comparable results and, therefore, gain reliable knowledge about the effect of PA and SB on the human microbiome.

## Supporting information

Electronic Supplementary Material

## Data Availability

Original data used in the present study are sourced from publicly available, peer-reviewed published articles. All data produced are contained in the manuscript.

## Acknowledgments

Projects SAF2017-87526-R, PID2021-127280OB-I00 and PID2020-120249RB-100 funded by MCIN/AEI/10.13039/501100011033/FEDER “Una manera de hacer Europa”; Project A-CTS-614-UGR20 funded by FEDER/Junta de Andalucía-Consejería de Economía y Conocimiento; Projects P20_00158 and P20_00124 funded by Junta de Andalucía; Unidad de excelencia SOMM17/6107/UGR funded by Plan Propio de Investigación/Universidad de Granada; S.A, I.P.P are supported by grants RYC-2016-21199, FPU19/05561, funded by MCIN/AEI/10.13039/501100011033 and FSE “El FSE invierte en tu futuro”. A.P.F contribution was funded in part by NIH grant #: U01 TR002004 (REACH project). E.U.G is supported by the María Zambrano fellowship by the Ministerio de Universidades and the Unión Europea–NextGenerationEU. This work is part of a Ph.D. thesis conducted in the Biomedicine Doctoral Studies of the University of Granada, Spain.

## Author contributions

I.P.P conceptualized, designed, and wrote the manuscript. A.P.F participated in all systematic review phases (except for writing) and reviewed the manuscript draft for important intellectual content. E.U.G participated in elaborating the meta-analysis and reviewed the initial manuscript draft for important intellectual content. F.B.O and S.A conceptualized and designed the review, supervised all the article processes and reviewed the manuscript for important intellectual content.

