## Supplementary material for "Physical Activity, Sedentary Behavior and Microbiome: A Systematic Review and Meta-analysis": Electronic Supplementary Material

**Electronic Supplementary Material** (ESM) of this article includes the following files:

- **Appendix S1: Supplementary Methods** (ESM_AppendixS1.docx): A detailed explanation of the quality assessment and the meta-analysis.
- **Appendix S2: Supplementary Results** (ESM_AppendixS2.docx): A detailed description of the results divided by type of study design.
- **Table S1** (ESM_TableS1.docx). Search terms used in PubMed, Web of Science, Scopus and Cochrane databases.
- **Table S2** (ESM_TableS2.docx). PRISMA main checklist 2020.
- **Table S3** (ESM_TableS3.docx). PRISMA abstract checklist 2020.
- **Table S4** (ESM_TableS4.docx). Quality assessment of cross-sectional studies.
- **Table S5** (ESM_TableS5.docx). Quality assessment of acute physical activity studies (i.e., acute effects).
- **Table S6** (ESM_TableS6.docx). Quality assessment of chronic physical activity (i.e., chronic effects) randomized controlled trials.
- **Table S7** (ESM_TableS7.docx). Quality assessment of chronic physical activity (i.e., chronic effects) non-randomized controlled trials.

### Appendix S1: Supplementary Methods

#### Quality assessment

The quality of the included studies was independently assessed by two researchers (I.P.P and A.P.F). The Joanna Briggs Institute Critical Appraisal Tool for Systematic Reviews was used for cross-sectional, non-randomized (non-RCT), and randomized controlled trials (RCT) [1]. The abovementioned tool for quality assessment includes different and specific checklists for each type of study design. Each item included in these checklists had 4 answers: "criterion met" (yes), "criterion not met" (no), "not applicable", or "unclear". We used a modified version of the Downs and Black checklist for studies that reported the acute effects of PA on microbiome [2]. This checklist, composed of 17 questions, has been adapted for the quality assessment of studies reporting the acute effects of PA on circulating blood biomarkers [3]. Each researcher carefully assessed each item and reported an overall appraisal. To provide a general indication of quality, a total quality score (%) per study was calculated by dividing the number of positively scored criteria in each study (i.e., points obtained after answering as "yes") by the total number of applicable criteria. When the quality score was ≥ 75%, studies were considered as "high quality", while those studies with a quality score < 75% were considered as "low quality" [4]. Possible discrepancies were solved through common consensus.

#### Data synthesis and meta-analysis

We conducted a meta-analysis including cross-sectional studies or trials reporting the effect of PA interventions (chronic effects) on gut microbiome diversity (specifically alpha-diversity, expressed by the Shannon diversity and Chao1 indexes) in healthy individuals. This decision was done based on the limited microbiome data available and the heterogeneity of the studies identified. Likewise, alpha-diversity is considered a key indicator of the gut microbiome and overall host health, that is generally thought to increase after PA interventions and is positively associated with PA levels in cross-sectional studies [5–8]. Importantly, we selected only healthy population because pathological phenotypes have been associated with a dysbiotic microbiome [9] and could not be appropriate for meta-analysis combining both healthy and unhealthy participants. Furthermore, an independent meta-analysis with studies including only unhealthy populations was not performed due to the heterogeneity of different diseases among studies and the low number of studies (≤ 3) with available microbiome data.

It is important to clarify the following aspects in our meta-analysis: (1) when microbiome diversity data (i.e., mean and standard deviation [SD]) to calculate effect sizes and perform the meta-analysis was not reported in the study, we requested this information from the authors. For those studies where we could not obtain the mean and SD in the article or by request, the Web-PlotDigitizer 4.4 software (Ankit Rohatgi [https://automeris.io/WebPlotDigitizer/]) was used to estimate the mean and SD from graphs reported in the study. The Web-PlotDigitizer is a web-based plot digitizing tool for extracting data from different plots and has proven valid and reliable [10–12]. In cases it was not possible to obtain the mean and SD for one study, we extracted the median, the interquartile range (IQR), and the maximum and minimum values from graphs of the study. To calculate the effect size, those values obtained as median and IQR from graphs were transformed into mean and SD by the Wan's method [13]. This method allows to estimate mean and SD by incorporating the sample size, median, IQR and maximum and minimum values. The Wan's method has shown a good performance when analyzing normally and non-normally distributed data, presenting more accurate mean and SD estimations than other methods (e.g. Hozo et al.'s method and Bland's method) [14]; (2) the first meta-analysis we performed for cross-sectional studies analyzes differences on microbiome alpha-diversity between groups of high and low PA levels. The effect sizes were computed for the groups with the highest and lowest PA levels in each study; (3) the second meta-analysis we performed for cross-sectional studies evaluated differences in terms of microbiome alpha-diversity in athletes *vs*. non-athletes; (4) the third meta-analysis included the studies that reported the chronic effects of PA (i.e., RCTs and non-RCTs) on microbiome alpha-diversity; (5) at least three studies were considered to conduct each meta-analysis; (6) meta-analysis for SB was not performed due to the scarce studies on the topic.

### Appendix S2: Supplementary Results

#### 1. Cross-sectional studies: physical activity and sedentary behavior (non-athletes)

*1.1 Healthy populations*

Five studies found a positive association between PA and gut and vaginal microbiome alpha-diversity [1–5], while one study reported a negative association in the gut [6]. In contrast, 8 studies did not report significant associations between PA level and alpha-diversity of the gut and lower respiratory tract microbiome [7–14]. Two studies positively correlated SB time with alpha-diversity of the gut and saliva microbiome [7,15]. Concerning beta-diversity analysis, 5 studies observed dissimilarities between the gut microbial communities of active compared to inactive individuals [1,2,5,10,12], while 6 did not detect any differences between study groups [7–9,13,14,16]. Two studies revealed dissimilarities at community-level in the gut and saliva microbial composition when compared groups of SB, expressed as screen time [8,15].

Seven studies reported positive associations between PA and the relative abundance of several gut bacteria such as *Lachnospiraceae*, *Erysipelotrichaceae*, *Roseburia*, *Coprococcus*, *Veillonella*, *Akkermansia municiphila* and *Faecalibacterium prausnitzii*, among others [1,2,5,7,8,14,17], whereas shifts at phylum level were variably reported [2,10,18]. Contrastingly, one study reported a lower abundance of *Roseburia*, *Faecalibacterium* and *Blautia*, among others, in the gut of more active individuals [12]. Four studies reported Firmicutes/Bacteroidetes ratio of the gut microbiome but did not find any associations with PA [7,8,10,18]. Five studies analyzed fecal or saliva samples and found positive associations between SB time and the relative abundance of *Desulfovibrionaceae, Bacteroides*, *Paraprevotella, Prevotella* and *Streptococcus*, among others [1,7,10,15,17].

*1.2 Participants with disease*

Two studies found a positive association between PA and alpha-diversity of the gut microbiome [19,20], whereas one study described a negative association [21]. However, 5 studies reported no associations [22–26]. Four studies correlated self-reported or objetive PA to microbial dissimilarities in the gut according to beta-diversity analysis [21,23,25,27], while one did not find significant differences [26].

Six studies identified positive associations between PA and several gut bacteria such as Actinobacteria, *Coprococcus*, *Flavobacterium*, *Cetobacterium* or *Bifidobacterium* [19,23,24,26], and negative associations with Firmicutes, *Bacteroides*, *Megamonas* or *Barnesiella* [21,26,28]. Two studies negatively associated SB (expressed as standing or screen time) with Actinobacteria, *Bifidobacterium*, *Lachnospiraceae* and *Ruminococcus*, among others, and positively with *Streptococcus* in the gut [25,29].

#### 2. Cross-sectional studies: athletes vs non-athletes

Eighteen cross-sectional studies compared athletes *vs*. non-athletes [13,30–46], 5 compared athletes from different competition levels (i.e., professional/elite *vs*. amateur/non-elite) [31,46–49], and 3 studies analyzed athletes from different sports [42,43,50]. Seven studies reported that alpha-diversity was higher in the gut microbiome of athletes compared to non-athletes [30,31,38–40,43,46], four studies showed a higher alpha-diversity in athletes from high compared to low competition levels [31,46,48,49], three studies reported a negative association between sport performance and alpha-diversity in the gut and oral microbiome [35,37,41], and 9 studies did not find any significant association [13,32–34,36,42,44,45,50]. In beta-diversity analyses, 8 studies found dissimilarities between gut and oral microbial communities from athletes *vs*. non-athletes' comparisons [30,33–35,39–41,44], while 5 studies did not find any differences [13,32,36,38,42]. Additionally, two studies described differences in beta-diversity in the gut microbiome of athletes from different levels or sports [49,50].

Nine studies reported that athletes presented a higher abundance of certain gut bacteria such as Firmicutes, *Prevotellaceae*, *Akkermansiaceae, Prevotella, Akkermansia, Faecalibacterium, A. municiphila* and *F. prausnatzii* and lower of Bacteroidetes and *Bacteroides*, compared to non-athletes [30,32–35,38,42–44]. Two studies reported microbial shifts in the gut microbiome of professional/elite *vs*. amateur/non-elite athletes, with higher abundances of Firmicutes and *Parabacteroides*, and lower of Bacteroidetes and *Megasphaera*, among others, in high-level athletes [48,49].

### 3. Acute effects of PA

Three studies detected an increased alpha-diversity in the gut microbiome following a rowing or run race [51–53], while another three studies did not find any changes in alpha- or beta-diversity analyses after a marathon or a treadmill test [54–56].

Six studies described significant changes in the relative abundance of several gut bacteria, such as increases in Firmicutes/Bacteroidetes ratio, *Coprococcus*, *Veillonella*, *Collinsella aerofaciens* or *F. prausnitzii*, among others, after a marathon, rowing or run race [51–54,56,57]. Two studies identified differentially abundant gut microbes after a treadmill or cycle ergometer test [55,58]. One study did not report any effect of acute PA on blood virome after a cycle ergometer test [59].

### 4. Chronic effects of PA

#### *4.1 Healthy populations*

Four studies detected a higher alpha-diversity in the gut microbial communities after aerobic PA interventions (ranging from 2 to 12 weeks, respectively) [32,60–62], whereas 10 studies did not report any effect of PA interventions (from 1.4 to 14 weeks) in fecal or saliva samples [63–72]. Regarding beta-diversity, three studies detected changes in beta-diversity analysis after aerobic PA (12, 9 and 7 weeks, respectively) [61,62,70]. One study described significant microbial changes in the gut during a phase of a swimming training coinciding with a decrease in training volume [60]. Seven studies did not find significant differences in beta-diversity after PA intervention (from 1.4 to 10 weeks) [32,63–66,68,69].

Five studies reported a higher abundance of specific gut bacteria such as Actinobacteria, *Faecalibacterium*, *Coprococcus*, *Eubacterium*, *Oscillospira*, *Bifidobacterium*, *Butyricimonas* or *Alistipes* after aerobic or combined aerobic and resistance PA interventions ranging from 2 to 9 weeks [60,62,64,65,70]. One study described a higher Firmicutes/Bacteroidetes ratio in the peak of a swimming training [60], while another study detected 8 differentially abundant gut bacteria after a football season [72]. However, four studies did not detect any microbial change after PA intervention [32,66,69,73].

#### *4.2 Participants with disease*

Three studies found an increased alpha-diversity in the gut microbiome after aerobic or combined aerobic and resistance PA interventions (8 and 24 weeks) in participants with non-alcoholic fatty liver disease or obesity [74–76], while 13 studies did not report any effect of PA interventions (from 2 to 34.4 weeks) on gut and oral alpha-diversity in patients with obesity, non-alcoholic fatty liver disease, insulin resistance or celiac disease, among others [77–89]. Furthermore, 11 studies observed dissimilarities in gut or oral microbial communities in beta-diversity analyses after aerobic or combined aerobic and resistance PA interventions ranging from 6 to 34.4 weeks [75,76,90,77–81,86,87,89], while four studies did not report any differences (PA interventions from 3 to 12 weeks) [82,84,85,88].

Fifteen studies found increases in Verrucomicrobia, Bacteroidetes, *Lachnospiraceae*, *Akkermansia*, *Veillonella*, *Faecalibacterium*, *Roseburia* and *Bifidobacterium*, and decreases in Proteobacteria, *Clostridium*, *Bacteroides* or *Prevotella,* among others, in fecal and oral samples after different PA interventions of variable duration (2 to 34.4 weeks) [74,75,86–90,76–80,83–85]. One study found a decreased Firmicutes/Bacteroidetes ratio after a 2 weeks-aerobic PA program [83].

8. Whisner CM, Maldonado J, Dente B, Krajmalnik-Brown R, Bruening M. Diet, physical activity and screen time but not body mass index are associated with the gut microbiome of a diverse cohort of college students living in university

housing: a cross-sectional study. *BMC Microbiol*. 2018;18(1):210. doi:10.1186/s12866-018-1362-x

35. Morishima S, Oda N, Ikeda H, et al. Altered Fecal Microbiotas and Organic Acid Concentrations Indicate Possible Gut Dysbiosis in University Rugby Players: An

Observational Study. *Microorganisms*. 2021;9(8). doi:10.3390/microorganisms9081687

48. Liang R, Zhang S, Peng X, et al. Characteristics of the gut microbiota in professional martial arts athletes: A comparison between different competition

levels. *PLoS One*. 2019;14(12):e0226240. doi:10.1371/journal.pone.0226240

doi:10.1055/a-1079-5450

**Table S1.** Search terms used in PubMed, Web of Science, Scopus and Cochrane databases.

### PubMed

**Exposure**

("sedentary behavior"[Title/Abstract] OR "sedentary behavior"[Title/Abstract] OR "sedentary time"[Title/Abstract] OR "Sedentary Lifestyle"[Title/Abstract] OR "Physical Inactivity"[Title/Abstract] OR exercise*[Title/Abstract] OR "Physical Activity"[Title/Abstract] OR "Physical Activities"[Title/Abstract] OR "sitting"[Title/Abstract] OR "Physical Fitness"[Title/Abstract] OR "Cardiorespiratory Fitness" OR "Physical Training" [Title/Abstract] OR Sport*[Title/Abstract] OR Athletic Performance*[Title/Abstract] OR Sport Performance*[Title/Abstract] OR "muscular strength")

**Outcomes**

("microbiome"[Title/Abstract] OR "microbiota"[Title/Abstract] OR "flora"[Title/Abstract] OR "microflora"[Title/Abstract] OR microorganism*[Title/Abstract] OR bacteria*[Title/Abstract] OR microbe*[Title/Abstract] OR "microbial community"[Title/Abstract] OR "virome"[Title/Abstract] OR metatranscriptom*[Title/Abstract] OR metagenom*[Title/Abstract] OR "DNA-Seq"[Title/Abstract] OR "16S rRNA"[Title/Abstract] OR "16S RNA"[Title/Abstract] OR "16S gene"[Title/Abstract] OR "16S DNA"[Title/Abstract])

### Web of Science

**Exposure**

TS=("sedentary behaviour" OR "sedentary behavior" OR "sedentary time" OR "Sedentary Lifestyle" OR "Physical Inactivity" OR exercise* OR "Physical Activity" OR "Physical Ac tivities" OR "sitting" OR "Physical Fitness" OR "Cardiorespiratory Fitness" OR "Physical Training" OR Sport* OR Athletic Performance* OR Sport Performance* OR "muscular strength")

**Outcomes**

TS=("microbiome" OR "microbiota" OR "flora" OR "microflora" OR microorganism* OR bacteria* OR microbe* OR "microbial community" OR "virome" OR metatranscriptom* OR metagenom* OR "DNA-Seq" OR "16S rRNA" OR "16S RNA" OR "16S gene" OR "16S DNA")

### Scopus

**Exposure**

TITLE-ABS ("sedentary behaviour" OR "sedentary behavior" OR "sedentary time" OR "Sedentary Lifestyle" OR "Physical Inactivity" OR "exercise*" OR "Physical Activity" OR "Physical Activities" OR "sitting" OR "Physical Fitness" OR "Cardiorespiratory Fitness" OR "Physical Training" OR "Sport*" OR "Athletic Performance*" OR "Sport Performance*" OR "muscular strength")

**Outcomes**

TITLE-ABS ("microbiome" OR "microbiota" OR "flora" OR "microflora" OR "microorganism*" OR "bacteria*" OR "microbe*" OR "microbial community" OR "virome" OR "metatranscriptom*" OR "metagenom*" OR "DNA-Seq" OR "16S rRNA" OR "16S RNA" OR "16S gene" OR "16S DNA")

### The Cochrane Library

**Exposure**

("sedentary behaviour" OR "sedentary behavior" OR "sedentary time" OR "Sedentary Lifestyle" OR "Physical Inactivity" OR exercise* OR "Physical Activity" OR "Physical Ac tivities" OR "sitting" OR "Physical Fitness" OR "Cardiorespiratory Fitness" OR "Physical Training" OR Sport* OR Athletic Performance* OR Sport Performance* OR "muscular strength")

**Outcomes**

("microbiome" OR "microbiota" OR "flora" OR "microflora" OR microorganism* OR bacteria* OR microbe* OR "microbial community" OR "virome" OR metatranscriptom* OR metagenom* OR "DNA-Seq" OR "16S rRNA" OR "16S RNA" OR "16S gene" OR "16S DNA")

**Table S2**. PRISMA main checklist 2020.

| Section and Topic | Item # | Checklist item | Location where item is reported |
|---|---|---|---|
| **TITLE** | | | |
| Title | 1 | Identify the report as a systematic review. | Page 1 |
| **ABSTRACT** | | | |
| Abstract | 2 | See the PRISMA 2020 for Abstracts checklist. | See Electronic Supplementary Table S3 |
| **INTRODUCTION** | | | |
| Rationale | 3 | Describe the rationale for the review in the context of existing knowledge. | Pages 5, 6 |
| Objectives | 4 | Provide an explicit statement of the objective(s) or question(s) the review addresses. | Page 6 |
| **METHODS** | | | |
| Eligibility criteria | 5 | Specify the inclusion and exclusion criteria for the review and how studies were grouped for the syntheses. | Page 7 |
| Information sources | 6 | Specify all databases, registers, websites, organisations, reference lists and other sources searched or consulted to identify studies. Specify the date when each source was last searched or consulted. | Page 6 |
| Search strategy | 7 | Present the full search strategies for all databases, registers and websites, including any filters and limits used. | Presented in Electronic Supplementary Table S1 and reflected on page 7 |
| Selection process | 8 | Specify the methods used to decide whether a study met the inclusion criteria of the review, including how many reviewers screened each record and each report retrieved, whether they worked independently, and if applicable, details of automation tools used in the process. | Page 7 |
| Data collection process | 9 | Specify the methods used to collect data from reports, including how many reviewers collected data from each report, whether they worked independently, any processes for obtaining or confirming data from study investigators, and if applicable, details of automation tools used in the process. | Page 7 |
| Data items | 10a | List and define all outcomes for which data were sought. Specify whether all results that were compatible with each outcome domain in each study were sought (e.g. for all measures, time points, analyses), and if not, the methods used to decide which results to collect. | Page 7 |
| | 10b | List and define all other variables for which data were sought (e.g. participant and intervention characteristics, funding sources). Describe any assumptions made about any missing or unclear information. | Page 7 |
| Study risk of bias assessment | 11 | Specify the methods used to assess risk of bias in the included studies, including details of the tool(s) used, how many reviewers assessed each study and whether they worked independently, and if applicable, details of automation tools used in the process. | Presented in Electronic Supplementary Appendix S1 and reflected on page 7 |
| Effect measures | 12 | Specify for each outcome the effect measure(s) (e.g. risk ratio, mean difference) used in the synthesis or presentation of results. | Page 8 |
| Synthesis | 13a | Describe the processes used to decide which studies were eligible for each synthesis (e.g. tabulating the study intervention characteristics | Pages 7, 8, |

| Section and Topic | Item # | Checklist item | Location where item is reported |
|---|---|---|---|
| methods | | and comparing against the planned groups for each synthesis (item #5)). | and Electronic Supplementary Appendix S1 |
| | 13b | Describe any methods required to prepare the data for presentation or synthesis, such as handling of missing summary statistics, or data conversions. | Electronic Supplementary Appendix S1 |
| | 13c | Describe any methods used to tabulate or visually display results of individual studies and syntheses. | Table 2 and Figure 2 |
| | 13d | Describe any methods used to synthesize results and provide a rationale for the choice(s). If meta-analysis was performed, describe the model(s), method(s) to identify the presence and extent of statistical heterogeneity, and software package(s) used. | Page 8 and Electronic Supplementary Appendix S1 |
| | 13e | Describe any methods used to explore possible causes of heterogeneity among study results (e.g. subgroup analysis, meta-regression). | Electronic Supplementary Appendix S1 |
| | 13f | Describe any sensitivity analyses conducted to assess robustness of the synthesized results. | NA |
| Reporting bias assessment | 14 | Describe any methods used to assess risk of bias due to missing results in a synthesis (arising from reporting biases). | Electronic Supplementary Appendix S1 |
| Certainty assessment | 15 | Describe any methods used to assess certainty (or confidence) in the body of evidence for an outcome. | Page 8 |
| **RESULTS** | | | |
| Study selection | 16a | Describe the results of the search and selection process, from the number of records identified in the search to the number of studies included in the review, ideally using a flow diagram. | Page 8 and Figure 1 |
| | 16b | Cite studies that might appear to meet the inclusion criteria, but which were excluded, and explain why they were excluded. | Figure 1 |
| Study characteristics | 17 | Cite each included study and present its characteristics. | Table 2 |
| Risk of bias in studies | 18 | Present assessments of risk of bias for each included study. | Electronic Supplementary Tables S4, S5, S6 and S7 |
| Results of individual studies | 19 | For all outcomes, present, for each study: (a) summary statistics for each group (where appropriate) and (b) an effect estimate and its precision (e.g. confidence/credible interval), ideally using structured tables or plots. | Figure 3 |
| Results of syntheses | 20a | For each synthesis, briefly summarise the characteristics and risk of bias among contributing studies. | Page 10 |
| | 20b | Present results of all statistical syntheses conducted. If meta-analysis was done, present for each the summary estimate and its precision (e.g. confidence/credible interval) and measures of statistical heterogeneity. If comparing groups, describe the direction of the effect. | Page 11 |
| | 20c | Present results of all investigations of possible causes of heterogeneity among study results. | Page 11 |
| | 20d | Present results of all sensitivity analyses conducted to assess the robustness of the synthesized results. | NA |
| Reporting biases | 21 | Present assessments of risk of bias due to missing results (arising from reporting biases) for each synthesis assessed. | NA |

| Section and Topic | Item # | Checklist item | Location where item is reported |
|---|---|---|---|
| Certainty of evidence | 22 | Present assessments of certainty (or confidence) in the body of evidence for each outcome assessed. | Page 11 |
| **DISCUSSION** | | | |
| Discussion | 23a | Provide a general interpretation of the results in the context of other evidence. | Pages 11, 12 |
| | 23b | Discuss any limitations of the evidence included in the review. | Pages 11-20 |
| | 23c | Discuss any limitations of the review processes used. | Page 19 |
| | 23d | Discuss implications of the results for practice, policy, and future research. | Pages 18-20 |
| **OTHER INFORMATION** | | | |
| Registration and protocol | 24a | Provide registration information for the review, including register name and registration number, or state that the review was not registered. | Page 6 |
| | 24b | Indicate where the review protocol can be accessed, or state that a protocol was not prepared. | Page 6 |
| | 24c | Describe and explain any amendments to information provided at registration or in the protocol. | NA. The review protocol can be updated in PROSPERO database after publishing the manuscript |
| Support | 25 | Describe sources of financial or non-financial support for the review, and the role of the funders or sponsors in the review. | Page 20 |
| Competing interests | 26 | Declare any competing interests of review authors. | Page 20 |
| Availability of data, code and other materials | 27 | Report which of the following are publicly available and where they can be found: template data collection forms; data extracted from included studies; data used for all analyses; analytic code; any other materials used in the review. | Data extracted from the articles presented in Table 2. Supplementary files are publicly available |

**Table S3**. PRISMA abstract checklist 2020.

| Topic | No. | Item | Reported? |
|---|---|---|---|
| **TITLE** | | | |
| **Title** | 1 | Identify the report as a systematic review. | Yes |
| **BACKGROUND** | | | |
| **Objectives** | 2 | Provide an explicit statement of the main objective(s) or question(s) the review addresses. | Yes |
| **METHODS** | | | |
| **Eligibility criteria** | 3 | Specify the inclusion and exclusion criteria for the review. | No |
| **Information sources** | 4 | Specify the information sources (e.g. databases, registers) used to identify studies and the date when each was last searched. | Yes |
| **Risk of bias** | 5 | Specify the methods used to assess risk of bias in the included studies. | No |
| **Synthesis of results** | 6 | Specify the methods used to present and synthesize results. | Yes |
| **RESULTS** | | | |
| **Included studies** | 7 | Give the total number of included studies and participants and summarise relevant characteristics of studies. | Yes |
| **Synthesis of results** | 8 | Present results for main outcomes, preferably indicating the number of included studies and participants for each. If meta-analysis was done, report the summary estimate and confidence/credible interval. If comparing groups, indicate the direction of the effect (i.e. which group is favoured). | Main outcomes and direction of the effect reported |
| **DISCUSSION** | | | |
| **Limitations of evidence** | 9 | Provide a brief summary of the limitations of the evidence included in the review (e.g. study risk of bias, inconsistency and imprecision). | Yes |
| **Interpretation** | 10 | Provide a general interpretation of the results and important implications. | Yes |
| **OTHER** | | | |
| **Funding** | 11 | Specify the primary source of funding for the review. | No |

| Topic | No. | Item | Reported? |
|---|---|---|---|
| **Registration** | 12 | Provide the register name and registration number. | No |

**Table S4**. Quality assessment of cross-sectional studies.

| Study | Item 1 | Item 2 | Item 3 | Item 4 | Item 5 | Item 6 | Item 7 | Item 8 | Quality score for each study | Quality category for each study |
|---|---|---|---|---|---|---|---|---|---|---|
| Clarke *et al*. 2014 | YES | YES | NA | NA | YES | YES | YES | NO | 71.4 | Low quality |
| Lim *et al*. 2016 | YES | YES | YES | NA | YES | YES | YES | NO | 85.7 | High quality |
| Shin *et al*. 2016 | YES | YES | YES | NA | NO | NO | YES | NO | 57.1 | Low quality |
| Bressa *et al*. 2017 | YES | NO | YES | YES | YES | YES | YES | YES | 87.5 | High quality |
| Barton *et al*. 2017 | NO | NO | NA | NA | YES | YES | YES | NO | 50.0 | Low quality |
| Petersen *et al*. 2017 | YES | NO | NA | NA | NO | NO | YES | NO | 33.3 | Low quality |
| Mörkl *et al*. 2017 | YES | YES | NA | YES | YES | YES | YES | YES | 100.0 | High quality |
| Bai *et al*. 2018 | YES | YES | NO | YES | YES | YES | YES | NO | 75.0 | High quality |
| Minty *et al*. 2018 | YES | YES | NA | NA | YES | NO | YES | NO | 66.7 | Low quality |
| Whisner *et al*. 2018 | YES | YES | YES | NA | NO | NO | YES | YES | 71.4 | Low quality |
| Mahnic *et al*. 2018 | YES | YES | NO | NA | YES | NO | YES | NO | 57.1 | Low quality |
| Carter *et al*. 2019 | YES | YES | YES | YES | YES | YES | YES | YES | 100.0 | High quality |
| Zhang *et al*. 2019 | YES | YES | NO | NA | NO | NO | YES | NO | 42.9 | Low quality |
| Langsetmo *et al*. 2019 | YES | YES | YES | NA | YES | YES | YES | NO | 85.7 | High quality |
| Jang *et al*. 2019 | YES | NO | NA | NA | NO | NO | YES | NO | 33.3 | Low quality |
| O'Donovan *et al*. 2019 | NO | NO | NA | NA | YES | YES | YES | NO | 50.0 | Low quality |

| | | | | | | | | | |
|---|---|---|---|---|---|---|---|---|---|
| Liang *et al*. 2019 | YES | YES | NA | NA | YES | YES | YES | YES | 100.0 | High quality |
| Castellanos *et al*. 2020 | YES | YES | YES | YES | YES | YES | YES | YES | 100.0 | High quality |
| Lin *et al*. 2020 | YES | YES | YES | YES | YES | YES | YES | YES | 100.0 | High quality |
| Zhu *et al*. 2020[b] | YES | NO | NO | NA | NO | NA | YES | NO | 33.3 | Low quality |
| Yoon *et al*. 2020 | YES | YES | YES | NA | NO | NO | YES | NO | 57.1 | Low quality |
| Kulecka *et al*. 2020 | YES | YES | NA | NA | YES | YES | YES | YES | 100.0 | High quality |
| Heinzel *et al*. 2020 | YES | NO | NO | YES | YES | YES | YES | YES | 75.0 | High quality |
| Castellanos *et al*. 2020 | NO | NO | YES | YES | NO | NO | YES | YES | 50.0 | Low quality |
| Song *et al*. 2020 | YES | YES | NO | NA | YES | YES | YES | YES | 85.7 | High quality |
| Gallè *et al*. 2020 | YES | YES | YES | NA | YES | YES | YES | NO | 85.7 | High quality |
| Fart *et al*. 2020 | YES | NO | YES | NA | YES | YES | YES | NO | 71.4 | Low quality |
| Han *et al*. 2020 | YES | YES | NA | NA | NO | NO | YES | YES | 66.7 | Low quality |
| Valeriani *et al*. 2020 | YES | YES | YES | NA | YES | YES | YES | NO | 85.7 | High quality |
| Penney *et al*. 2020 | YES | NO | NA | NA | YES | YES | YES | YES | 83.3 | High quality |
| Genç *et al*. 2020 | YES | NO | NA | NA | NO | NO | YES | NO | 33.3 | Low quality |
| Özkan *et al*. 2020 | YES | NO | NA | NA | NO | NO | YES | NO | 33.3 | Low quality |
| Palmas *et al*. 2021 | YES | YES | YES | YES | YES | YES | YES | YES | 100.0 | High quality |
| Zhong et *al*. 2021 | YES | YES | YES | NA | YES | YES | YES | YES | 100.0 | High quality |
| Engberg *et al*. 2021 | NO | NO | YES | NA | YES | YES | YES | YES | 71.4 | Low quality |

| | | | | | | | | | |
|---|---|---|---|---|---|---|---|---|---|
| Morishima *et al*. 2021 | NO | NO | NA | NA | NO | NO | YES | NO | 16.7 | Low quality |
| Šoltys *et al*. 2021 | YES | YES | NA | NA | YES | NO | YES | YES | 83.3 | High quality |
| Jie *et al*. 2021 | NO | NO | NO | NA | YES | YES | YES | YES | 57.1 | Low quality |
| Morishima *et al*. 2021 | YES | YES | NA | NA | NO | NO | YES | NO | 50.0 | Low quality |
| Babszky *et al*. 2021 | YES | NO | NA | YES | NO | NO | YES | NO | 42.9 | Low quality |
| Houttu *et al*. 2021 | NO | YES | YES | NA | YES | YES | YES | YES | 85.7 | High quality |
| Walker *et al*. 2021 | YES | YES | NO | YES | YES | YES | YES | YES | 87.5 | High quality |
| Santarossa *et al*. 2021 | YES | YES | YES | NA | YES | YES | YES | NO | 85.7 | High quality |
| Magzal *et al*. 2022 | YES | NO | YES | YES | YES | YES | YES | YES | 87.5 | High quality |
| Xu *et al*. 2022 | YES | YES | YES | NA | YES | YES | YES | YES | 100.0 | High quality |
| Hintikka *et al*. 2022 | YES | YES | NA | NA | YES | YES | YES | NO | 83.3 | High quality |
| Shivani *et al*. 2022 | YES | YES | NO | NO | NO | NO | YES | YES | 50.0 | Low quality |
| Cuthbertson *et al*. 2022 | YES | YES | NA | YES | NO | NO | YES | NO | 57.1 | Low quality |
| Lubomski *et al*. 2022 | YES | YES | YES | YES | YES | YES | YES | YES | 100.0 | High quality |
| Visuthranukul *et al*. 2022 | YES | YES | NO | YES | YES | YES | YES | YES | 87.5 | High quality |

The quality score for each study (%) was calculated by dividing the number of positively scored criteria (i.e., points obtained after answering as "yes") by the total number of criteria (i.e., 8). All the items sum 1 point if the answer is "yes". A study was considered as "high quality" when the quality score was at least 75%, whereas studies were considered as "low quality" when the quality score was lower than 75%. Yes: meet the criterion; No: not meet the criterion; NA: not applicable criterion.

Quality assessment was performed using the Joanna Briggs Institute Critical Appraisal Tool for Systematic Reviews and, in particular, the Checklist for Analytical Cross-Sectional Studies which consists of 8 items. Item 1: Were the criteria for inclusion in the sample clearly defined?; Item 2: Were the study subjects and the setting described in detail?; Item 3: Was the exposure measured in a valid and reliable way?; Item 4: Were objective, standard criteria used for measurement of the condition?; Item 5: Were confounding factors identified?; Item 6: Were strategies to deal with confounding factors stated?; Item 7: Were the outcomes measured in a valid and reliable way?; Item 8: Was appropriate statistical analysis used?

**Table S5**. Quality assessment of acute physical activity studies (i.e., acute effects).

| Study | Item 1 | Item 2 | Item 3 | Item 4 | Item 5 | Item 6 | Item 7 | Item 8 | Item 9 | Item 10 | Item 11 | Item 12 | Item 13 | Item 14 | Item 15 | Item 16 | Item 17 | Quality score for each study | Quality category for each study |
|---|---|---|---|---|---|---|---|---|---|---|---|---|---|---|---|---|---|---|---|
| Shukla *et al*. 2015 | YES | YES | YES | YES | YES | NO | YES | YES | YES | YES | YES | YES | YES | YES | NO | NO | NO | 76.5 | High quality |
| Bouquet *et al*. 2019 | YES | YES | YES | YES | YES | YES | YES | YES | YES | YES | YES | NO | YES | YES | NO | NO | NO | 76.5 | High quality |
| Zhao *et al*. 2018 | YES | YES | YES | YES | YES | YES | YES | YES | YES | YES | YES | YES | YES | YES | YES | YES | YES | 100 | High quality |
| Keohane *et al*. 2019 | YES | YES | YES | YES | YES | NO | YES | YES | YES | YES | NO | YES | YES | YES | YES | YES | YES | 88.2 | High quality |
| Scheiman *et al*. 2019 | YES | YES | YES | YES | YES | YES | YES | YES | YES | YES | NO | YES | YES | YES | YES | YES | YES | 94.1 | High quality |
| Grosicki *et al*. 2019 | YES | YES | YES | YES | NO | YES | YES | YES | YES | YES | NO | NO | YES | YES | YES | NO | NO | 70.6 | Low quality |
| Tabone *et al*. 2021 | YES | YES | YES | YES | NO | NO | YES | YES | YES | YES | NO | YES | YES | YES | YES | YES | NO | 76.5 | High quality |
| Fukuchi *et al*. 2022 | YES | YES | YES | NO | YES | YES | YES | YES | YES | YES | YES | YES | YES | YES | YES | YES | YES | 94.1 | High quality |
| Sato *et al*. 2022 | YES | YES | YES | YES | YES | YES | YES | YES | YES | YES | YES | NO | YES | YES | YES | YES | YES | 94.1 | High quality |

The quality score for each study (%) was calculated by dividing the number of positively scored criteria (i.e., points obtained after answering as "yes") by the total number of criteria (i.e., 17). All the items sum 1 point if the answer is "yes". A study was considered as "high quality" when the quality score was at least 75%, whereas studies were considered as "low quality" when the quality score was lower than 75%. Yes: meet the criterion; No: not meet the criterion; NA: not applicable criterion; Pts: Points.

Quality assessment was performed using a modified version of the Downs and Black checklist which consists of 17 items scored with points. Each item that sums 1 or more points was considered to meet the criterion (i.e., "yes"). Item 1: Is the hypothesis/aim/objective of the study clearly described?; Item 2: Are the main outcomes to be measured clearly described in the introduction or methods section?; Item 3: Are the characteristics (e.g., age, height, weight, training and health status) of the participants included in the study clearly described?; Item 4: Are the interventions of interest clearly described?; Item 5: Are the main findings of the study clearly described?; Item 6: Does the study provide estimates of the random variability in the data for the main outcomes?; Item 7: Have all important adverse events that may be a consequence of the intervention been reported?; Item 8: Was an attempt made to blind study subjects to the intervention they have received? For exercise interventions where it is not possible to blind, answer yes; Item 9: Was an attempt made to blind those measuring the main outcomes of the intervention? For exercise interventions where it is not possible to blind, answer yes; Item 10: If any of the results of the study were based on 'data dredging' was this made clear?; Item 11: Was the timing of sampling clearly described?; Item 12: Were the statistical tests used to assess the main outcomes appropriate?; Item 13: Were the main outcome measures used accurate (valid and reliable)?; Item 14: Were study subjects randomised to intervention groups? Answer yes if the order of treatment, or allocation to groups, was randomly assigned. If it was not possible for the study to be randomised (e.g., single-trial studies) answer yes; Item 15: Was at least one familiarization session conducted prior to exercise testing? Answer yes if they conducted a familiarization trial, or if familiarization was not necessary (e.g., if the study uses a single, non-performance-based, exercise bout); Item 16: Were the exercise test conditions adequately standardised and described?; Item 17: Was nutritional status for sampling adequately described?

**Table S6**. Quality assessment of chronic physical activity (i.e., chronic effects) randomized controlled trials.

| Study | Item 1 | Item 2 | Item 3 | Item 4 | Item 5 | Item 6 | Item 7 | Item 8 | Item 9 | Item 10 | Item 11 | Item 12 | Item 13 | Quality score of each study | Quality category of each study |
|---|---|---|---|---|---|---|---|---|---|---|---|---|---|---|---|
| Cronin *et al*. 2018 | NO | NO | YES | NA | NA | NO | YES | NO | NO | YES | YES | NO | YES | 45.5 | Low quality |
| Taniguchi *et al*. 2018 | NO | NO | YES | NA | NA | NO | YES | NO | NO | YES | YES | YES | NO | 45.5 | Low quality |
| Cronin *et al*. 2019 | YES | NO | YES | NA | NA | NO | YES | NO | YES | YES | YES | YES | NO | 63.6 | Low quality |
| Kern *et al*. 2019 | YES | NO | YES | NA | NA | NO | YES | NO | YES | YES | YES | YES | YES | 72.7 | Low quality |
| Liu *et al*. 2020 | YES | YES | YES | NA | NA | NO | YES | NO | NO | YES | YES | YES | YES | 72.7 | Low quality |
| Quiroga *et al*. 2020 | NO | NO | YES | NA | NA | NO | YES | NO | NO | YES | YES | NO | YES | 45.5 | Low quality |
| Warbeck *et al*. 2020 | YES | NO | YES | NA | NA | NO | YES | NO | NO | YES | YES | NO | YES | 54.5 | Low quality |
| Zhong *et al*. 2021 | YES | NO | YES | NA | NA | NO | YES | NO | NO | YES | YES | NO | YES | 54.5 | Low quality |
| Moitinho-Silva *et al.* 2021 | NO | NO | YES | NA | NA | NO | YES | NO | NO | YES | YES | NO | YES | 45.5 | Low quality |
| Resende *et al*. 2021 | NO | NO | YES | NA | NA | NO | YES | YES | NO | YES | YES | YES | YES | 63.6 | Low quality |
| Lkhagva *et al*. 2021 | YES | NO | YES | NA | NA | NO | YES | NO | NO | YES | YES | YES | YES | 63.6 | Low quality |
| Torquati *et al*. 2021 | YES | YES | NO | NA | NA | NO | YES | YES | NO | YES | YES | YES | YES | 72.7 | Low quality |
| Cheng *et al*. 2022 | YES | YES | YES | NA | NA | YES | YES | YES | NO | YES | YES | YES | YES | 90.9 | High quality |
| Dupuit *et al*. 2022 | NO | NO | YES | NA | NA | NO | YES | YES | NO | YES | YES | YES | YES | 63.6 | Low quality |
| Bielik *et al*. 2022 | NO | NO | YES | NA | NA | NO | YES | YES | NO | YES | YES | YES | YES | 63.6 | Low quality |

The quality score for each study (%) was calculated by dividing the number of positively scored criteria (i.e., points obtained after answering as "yes") by the total number of criteria (i.e., 13). All the questions sum 1 point if the answer is "yes". A study was considered as "high quality" when the quality score was at least 75%, whereas studies were considered as "low quality" when the quality score was lower than 75%. Yes: meet the criterion; No: not meet the criterion; NA: not applicable criterion.

Quality assessment was performed using the Joanna Briggs Institute Critical Appraisal Tool for Systematic Reviews and, in particular, the Checklist for Randomized Controlled Trials which consists of 13 items. Item 1: Was true randomization used for assignment of participants to treatment groups?; Item 2: Was allocation to treatment groups concealed?; Item 3: Were treatment groups similar at the baseline?; Item 4: Were participants blind to treatment assignment?; Item 5: Were those delivering treatment blind to treatment assignment?; Item 6: Were outcomes assessors blind to treatment assignment?; Item 7: Were treatment groups treated identically other than the intervention of interest?; Item 8: Was follow up complete and if not, were differences between groups in terms of their follow up adequately described and analyzed?; Item 9: Were participants analyzed in the groups to which they were randomized?; Item 10: Were outcomes measured in the same way for treatment groups?; Item 11: Were outcomes measured in a reliable way?; Item 12: Was appropriate statistical analysis used?; Item 13: Was the trial design appropriate, and any deviations from the standard RCT design (individual randomization, parallel groups) accounted for in the conduct and analysis of the trial?

**Table S7**. Quality assessment of chronic physical activity (i.e., chronic effects) non-randomized controlled trials.

| Study | Item 1 | Item 2 | Item 3 | Item 4 | Item 5 | Item 6 | Item 7 | Item 8 | Item 9 | Quality score of each study | Quality category of each study |
|---|---|---|---|---|---|---|---|---|---|---|---|
| Allen *et al*. 2018 | YES | YES | NA | NO | NA | YES | YES | YES | YES | 85.7 | High quality |
| Munukka *et al*. 2018 | YES | YES | NA | NO | NA | YES | YES | YES | YES | 85.7 | High quality |
| Huber *et al*. 2019 | YES | YES | NA | NO | NA | NO | YES | YES | YES | 71.4 | Low quality |
| Motiani *et al*. 2019 | YES | YES | NA | NO | NA | YES | YES | YES | YES | 85.7 | High quality |
| Hampton-Marcell *et al*. 2019 | YES | YES | NA | NO | NA | YES | YES | YES | YES | 85.7 | High quality |
| Rettedal *et al*. 2020 | YES | YES | NA | NO | NA | YES | YES | YES | YES | 85.7 | High quality |
| Bycura *et al*. 2021 | YES | YES | NA | NO | NA | NO | YES | YES | YES | 71.4 | Low quality |
| Barton *et al*. 2021 | YES | YES | NA | NO | NA | YES | YES | YES | NO | 71.4 | Low quality |
| Uchida et al. 2021 | YES | YES | NA | NO | NA | NO | YES | YES | NO | 57.1 | Low quality |
| Feng *et al*. 2021 | YES | NO | NA | NO | NA | YES | YES | YES | YES | 71.4 | Low quality |
| Craven *et al*. 2021 | YES | YES | NA | NO | NA | YES | YES | YES | YES | 85.7 | High quality |
| Verheggen *et al*. 2021 | YES | YES | NA | NO | NA | YES | YES | YES | YES | 85.7 | High quality |
| Erlandson *et al*. 2021 | YES | YES | NA | NO | NA | YES | YES | YES | YES | 85.7 | High quality |
| Oliveira *et al*. 2021 | YES | YES | NA | NO | NA | YES | YES | YES | YES | 85.7 | High quality |
| Donati Zeppa *et al*. 2021 | YES | YES | NA | NO | NA | YES | YES | YES | YES | 85.7 | High quality |

| | | | | | | | | | | |
|---|---|---|---|---|---|---|---|---|---|---|
| Villarroel *et al*. 2021 | YES | YES | NA | NO | NA | YES | YES | YES | YES | 85.7 | High quality |
| Soriano *et al*. 2022 | YES | YES | NA | NO | NA | YES | YES | YES | YES | 85.7 | High quality |

The quality score for each study (%) was calculated by dividing the number of positively scored criteria (i.e., points obtained after answering as "yes") by the total number of criteria (i.e., 9). All the questions sum 1 point if the answer is "yes". A study was considered as "high quality" when the quality score was at least 75%, whereas studies were considered as "low quality" when the quality score was lower than 75%. Yes: meet the criterion; No: not meet the criterion; NA: not applicable criterion.

Quality assessment was performed using the Joanna Briggs Institute Critical Appraisal Tool for Systematic Reviews and, in particular, the Checklist for Quasi-experimental Studies which consists of 9 items. Item 1: Is it clear in the study what is the "cause" and what is the "effect"?; Item 2: Were the participants included in any comparisons similar?; Item 3: Were the participants included in any comparisons receiving similar treatment/care, other than the exposure or intervention of interest?; Item 4: Was there a control group?; Item 5: Were there multiple measurements of the outcome both pre and post the intervention/exposure?; Item 6: Was follow up complete and if not, were differences between groups in terms of their follow up adequately described and analyzed?; Item 7: Were the outcomes of participants included in any comparisons measured in the same way?; Item 8: Were outcomes measured in a reliable way?; Item 9: Was appropriate statistical analysis used?
